# Age and the Diurnal Oscillatory Features of the Human Chronobiome

**DOI:** 10.64898/2026.01.21.26344528

**Authors:** Carsten Skarke, Nicholas F. Lahens, Antonijo Mrčela, Niclas Olsson, Joseph Glessner, Amruta Naik, Katherine N. Theken, Caroline Chivily, Declan Douglas, Kyle N. Hess, Aleksandr Gaun, Ahmed M. Moustafa, Scott G. Daniel, Kyle Bittinger, Sarah Teegarden, Ronan Lordan, Nadim El Jamal, Hu Meng, Taylor Hollingsworth, Ashley Woolfork, Arjun Sengupta, Thomas G. Brooks, Aalim Weljie, Gregory Grant, Jonathon O’Brien, Fiona McAllister, Eugene Melamud, Garret A. FitzGerald

**Author notes:** These authors contributed equally (Nicholas F. Lahens, Antonijo Mrčela). Corresponding Authors: Carsten Skarke, MD & Garret A. FitzGerald, MD, Institute for Translational Medicine and Therapeutics (ITMAT), University of Pennsylvania Perelman School of Medicine, Smilow Center for Translational Research 10-101, 3400 Civic Center Blvd, Philadelphia, Pennsylvania 19104, USA.

## Abstract

The molecular clock regulates diverse aspects of human biology. As people age, diurnal rhythms deteriorate, most evidently in the daytime napping and nighttime waking of older individuals. To understand how temporal deconsolidation of oscillatory networks could contribute to age-related disease expression, we studied the chronobiome at unprecedented depth in young and old apparently healthy individuals. Transomic integration segregated age groups and identified candidate mechanisms by which oscillatory function might contribute to age dependent distinctions. In an orthogonal approach, we validated as true cyclers many proteins identified in the UK Biobank as predictors of health and disease outcomes. Here, age-specific alterations in the cycling proteome across disease phenotypes is consistent with our hypothesis that deconsolidated circadian programs associate with increased susceptibility to age-related disease.

## Introduction

Studies in model systems imply the importance of the molecular clock in the regulation of diverse aspects of human biology. Indeed, the importance of the clock has been supported by the association of human genetic variants with sleep syndromes and the recapitulation of these phenotypes in model systems ^1^. A second approach involves postmortem samples, using algorithms that adjust for differences between the time of death and tissue harvesting, yielding distinct circadian features across multiple tissues that reflect sex and age ^2^. Finally, the use of forced desynchrony protocols in highly controlled environments has allowed for segregation of endogenous from environmentally driven time-dependent rhythms and has revealed the impact of jet lag and feeding regimens on cardio-metabolic and immune function in volunteers ^3^. However, each of these approaches has intrinsic limitations. While multiple variants in core clock genes have been associated with phenotypic traits, few have been investigated with respect to their functional relevance. Postmortem analyses are critically dependent on the validity of the algorithms that adjust for differences between the time of death and tissue harvesting, and forced desynchrony protocols are performed in highly controlled laboratory settings. Some years ago, we reported a complementary approach – the integration of multiomics analyses with remote sensing to characterize diurnally oscillating features - the chronobiome - in free ranging humans “in the wild” ^4^. In a pilot study of 6 volunteers, we could discern elements of the multiome where variance was predominantly attributable to time of sampling rather than inter-individual differences.

As people age, the consolidation of their diurnal rhythms deteriorates, most evidently in the daytime napping and nighttime waking of older individuals ^5^. We hypothesize that temporal deconsolidation of oscillatory networks that integrate organ function to maintain metabolic and immunological homeostasis contributes to age-related disease expression. As a first step to address this hypothesis, we sought age related differences in the chronobiomes of apparently healthy individuals. This produced the largest dataset to date with unprecedented time-specific deep coverage of phenomic and multiomic outputs over two circadian cycles. In an orthogonal approach, we utilized the frequency of sampling over 48 hours in this study to validate as true cyclers many proteins identified in the UK Biobank as predictors of health and disease outcomes ^6,7^. Here, age-specific changes in the cycling proteome across disease phenotypes is consistent with our hypothesis that deconsolidated circadian programs associate with increased susceptibility to age related disease.

## Results

We collected 143 million data points across 911,000 features from apparently healthy younger (21-28 years) and older (56-74 years) volunteers (n=10 per cohort, *Supplementary Figure 1*) to seek an age-dependent distinction in the chronobiome (*Supplementary Table 1, Supplementary Table 2*). These datapoints come from a mix of wearable devices, surveys, and molecular analyses of biosamples. We excluded night shift workers, people with recent transmeridian travel and pre-existing conditions.

### Age-Specific Features in the Human Chronobiome

We observed expected signals in the aged: a shift to earlier chronotypes ^8^, earlier onset of physical activity, lower grip strength, lower cognitive response time, lower auditory function, higher 24-hour blood pressure profile and dipping ratio, a 24-hour sympathovagal balance with a higher sympathetic and lower parasympathetic function, higher glucose point-of-care measurements, and phase-advanced cortisol rhythms (cosinor *q*<0.0001 young/old, phase ɸ9.9H young, ɸ8.3H old, *Figure 1 A-C*). The two cohorts were clearly separated by chronological age (*Figure 1 D*). Biological age determined from Horvath epigenetic age estimates ^9^ similarly supported separation between young and old (*Figure 1 A*), and showed intra-individual fluctuations across the repeated assessments over 48hrs (mean±SD minimum/maximum differences 6.9±2.7 years in young, 13.8±4.6 years in old). An epigenetic age estimator developed in-house suggested diurnal variability in estimates of biological age, but this did not attain statistical significance (*Figure 1 E*).

**Figure 1.**
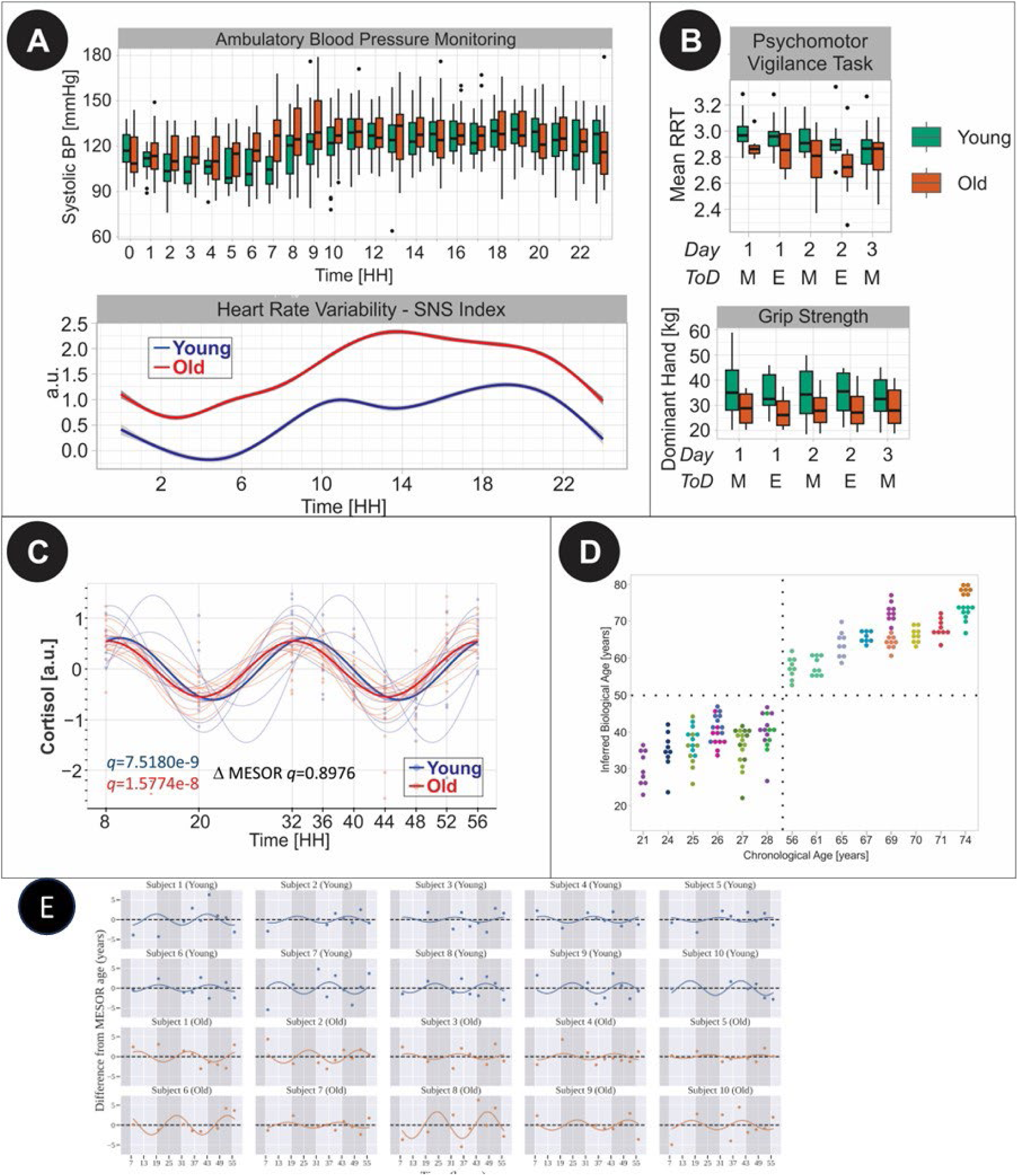
(A) Separation of young and old apparent healthy volunteer groups by chronological age (at time of study enrollment, abscissa) and biological age (ordinate), latter derived from Horvath epigenetic estimates on methylation signatures. Note the variability in the biological age. Subjects are color-coded to distinguish participants with the same chronological age. Single data points indicate the nine repeat measurements per subject over 48 hours starting with a 12hr sampling schedule on day 1 at 08:00 and 20:00, followed by 4hr sampling schedule starting at 08:00 on the next morning. (B) Lower cognitive function and grip strength in old compared to young assessed in the mornings (M) and evenings (E) on day 1, 2 and 3. ToD indicates time-of-day. Boxplots with median and 1st, 3rd quartile. (C) Significant oscillatory patterns in plasma cortisol abundances fitted over 48 hours in both young and old. Differences in mesor cortisol abundance were not detected between young and old. Cosine fits across young and old individuals (light color) and groups (dark color) over 48 hours. (D) Top: Higher systolic blood pressure in old compared to young assessed from 24-hour ambulatory blood pressure monitoring. Both age groups showed the dipping phenotype during sleep time and the morning surge. Boxplots with median and 1st, 3rd quartile. Bottom: Higher sympathetic nervous system (SNS) activation in old compared to young over the course of 24 hours. The SNS index was calculated from heart rate variability readouts based on continuous EKG monitoring. (E) Differences from subject-level MESORs of age predictions using novel elastic net model, across subjects and time points, together with subject-level cosinor fits.

### Diurnal Omics Landscape

We simultaneously analyzed the omics datasets from both age cohorts for oscillatory patterns using cosinor regression and non-linear fits (see materials for full details). This approach allows us to identify molecule features with age-dependent differences (delta-MESOR test), time-specific variance (cosinor parameter tests), and the interaction between age- and time-dependent variability (cosinor interaction tests, Bayesian Information Criterion [BIC], and two-factor tests).

We distinguished age-specific differences in abundance for 15.2% of the transcriptome (2844/18728), 21.8% of the methylome (140623/644928), 2.7% of the O-link measured (AntiBody)-proteome (79/2943) and 5.1% of the Mass Spectrometric measured (MS)-proteome (20/394) (Δ MESOR *q*<0.05, *Supplementary Table 3*).

Using a stringent threshold value of *q*<0.05, we detected oscillatory signals in large parts of the transcriptome. A total of 21.1% and 29.1% of features were rhythmic in young (3945/18728) and old (5444/18728), respectively (*Figure 2a*); this difference did not attain statistical significance (*p*=0.386 permuting the subject-level age labels 500 times). Notably, enrichment of rhythmic features was skewed towards the evening (*Supplementary Figure 2a top*). Oscillations in the methylome were limited to the old (0.12%, 758/644928) compared to the young (0/644928). Few tests for oscillatory signals in the methylome dataset survived multiple testing correction, due to the high number of features. The AB-proteome with panels selected for their association with health and disease counted more cyclers in the young (43.2%, 1262/2925) compared to the old (39.1%, 1144/2925). Notably, aging-associated features predictive of mortality ^10^ were elevated in old but differed in the absence (GDF15) or presence of oscillatory patterns (WFDC2, FGF21) (*Figure 2b*). The unbiased MS-proteome showed a similar fraction of cyclers in young (25.9%, 102/394) and old (25.6%, 101/394). We found that cycling AB-proteins were enriched in the evening (*Supplementary Figure 2a bottom*). More than double the metabolites were rhythmic in the old (38.6%, 59/153) in contrast to the young (15.7%, 24/153). Heatmaps are shown in *Supplementary Figure 2b*. Few cyclers emerged for the urine lipidome and the saliva microbiome (*Supplementary Table 3*).

**Figure 2.**
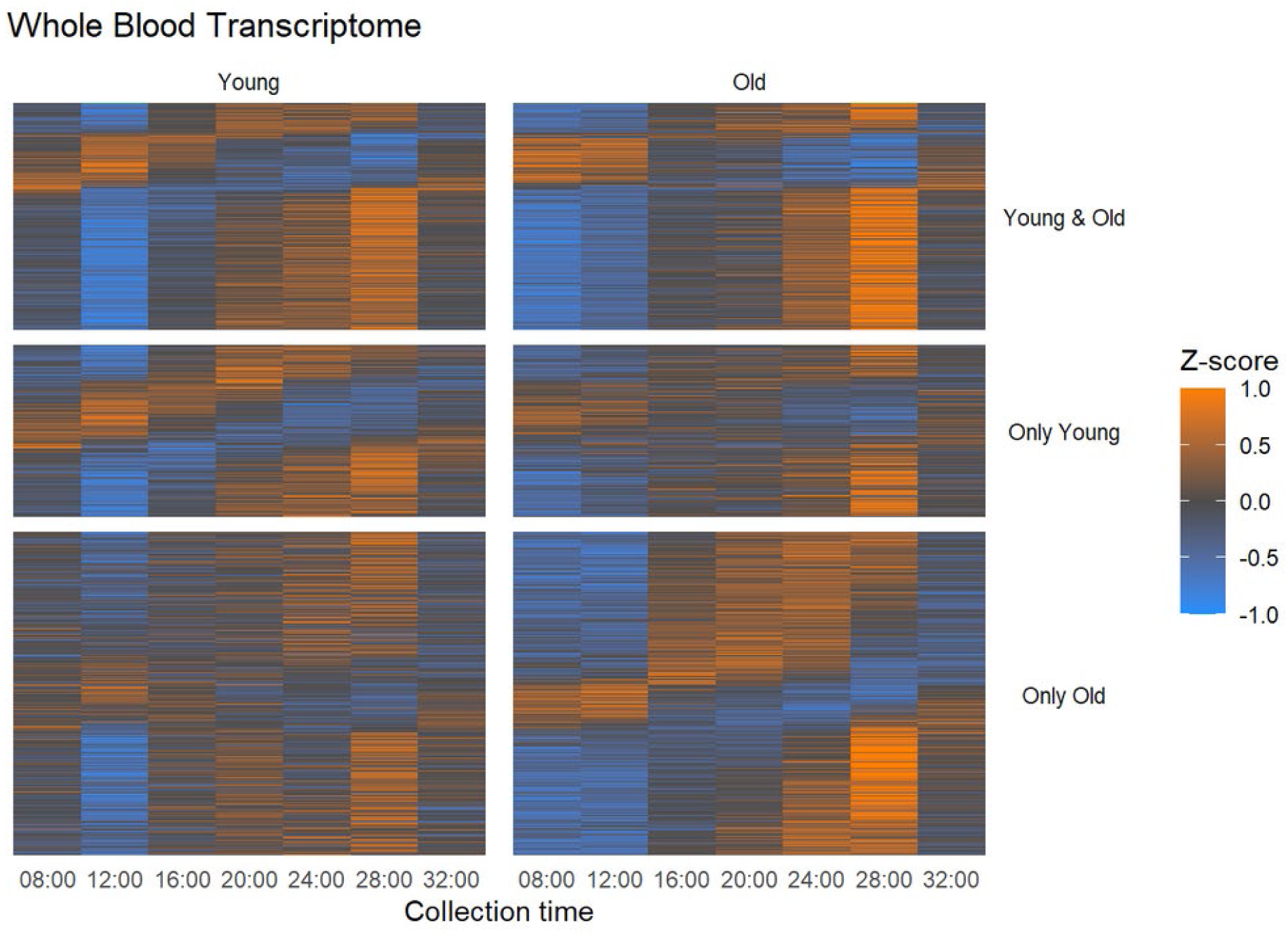
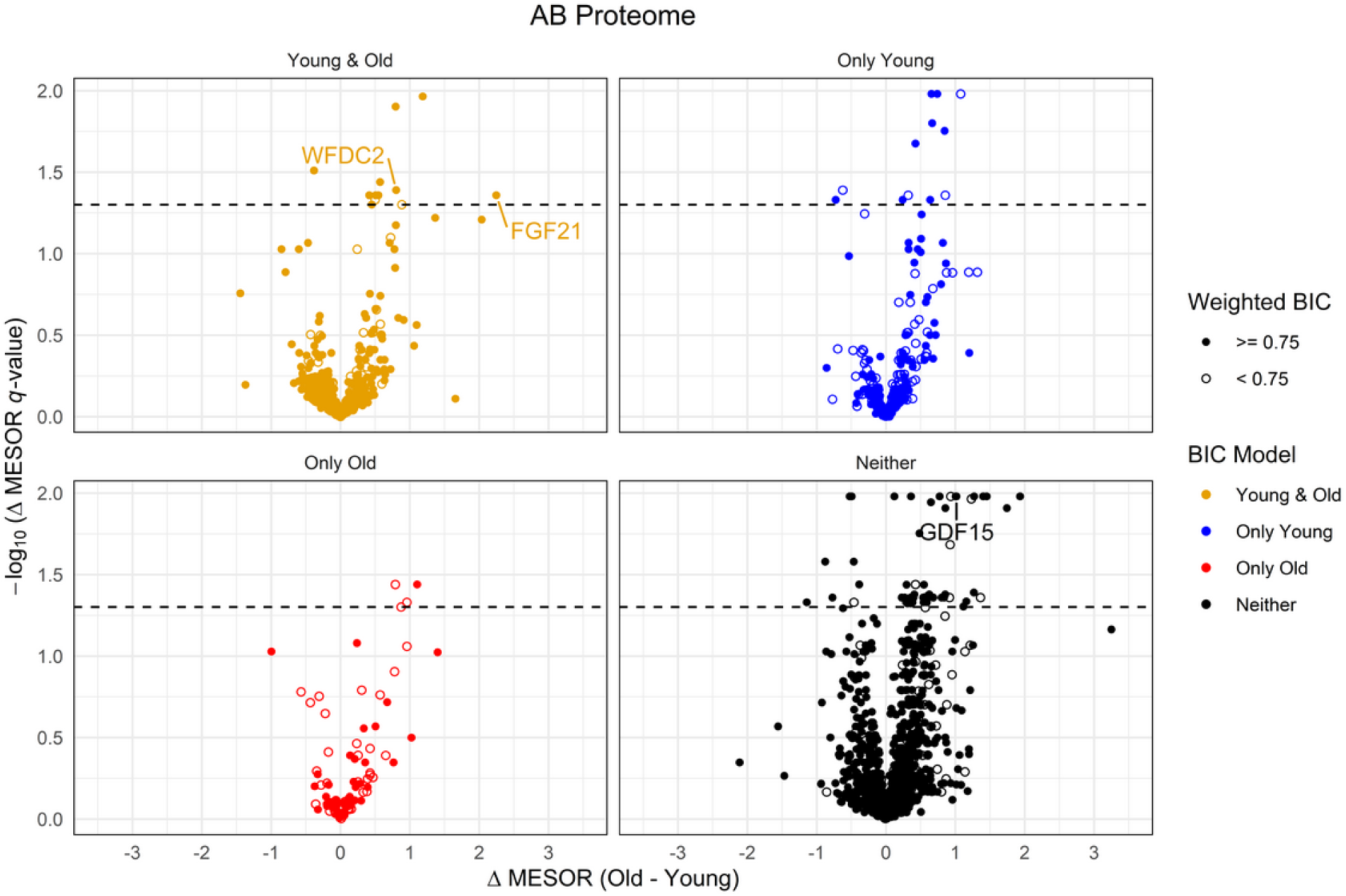
(A) Heatmaps for transcriptome in young (left column) and old (right column) visualize the variability in feature abundance quantified in 4hr intervals over 24 hours. The top rectangles feature oscillatory transcripts in young and old while subsets of transcripts oscillate either in young only (center) or old only (bottom). (B) Volcano plots displaying age-specific differences in the abundance and its level of significance of the features measured in the AB-Proteome.

While we could identify features that varied with time or by age group, we were underpowered to detect oscillatory features modified by age, that is, the interaction of age x time.

### Core Clock Transcripts Oscillate Robustly

Examination of the central core clock machinery offers insight into the temporal alignment of its components and how this is modified by age. Interrogating n=929 protein-coding transcripts annotated in GeneCards ^11^ with core circadian clock and clock-related gene function, we found that many transcripts share rhythmicity in young and old (n=333), including most of the core circadian clock genes (*Supplementary Figure 3A*). This suggests that aging per se does not deconsolidate central clock machinery at the gene expression level. Oscillations were evident in the positive feedback loop, *CLOCK* (cosinor *q*=0.07 young, cosinor *q*=0.02 old), the negative feedback loop, *PER1*, *PER2* and *PER3* (cosinor *q*<0.0001 young/old), and the interlocked feedback loop, *NR1D1* (cosinor *q*<0.001 young/old), *RORA* (cosinor *q*=0.02 young, cosinor *q*=0.04 old) (*Figure 3A*). The temporal alignment between positive and negative feedback loops underscored the internal consistency of this dataset. *CLOCK* peaked around midnight (ɸ23-1H) while *PER1* was phase-shifted to the early morning hours (ɸ6-7H). The differences between young and old were subtle. Only DBP trended toward higher abundance in young compared to old (Δ MESOR *q*=0.0988) (*Supplementary Figure 3B*). As expected, transcriptional activity in the old was phase-advanced by ≥1 hour, for example in *PER2* (ɸ5.6H young, ɸ4.3H old) or *NR1D1* (ɸ2H young, ɸ0.5H old), based on the cosinor fits. However, our sampling scheme limits assessments of acrophase differences at this resolution. Notably, the phase-agreement of the *PER* homologs contrasted with the subject-level divergence of phases in *CRY1* expression. Accounting for variability in phase across individuals, we detected oscillations for *CRY1* in young (*q_non-linear-fit_*=0.046) and old (*q_non-linear-fit_*=0.002) which had been missed in the cosinor regression model (young *cosinor-q=*0.45, old *cosinor-q=*0.29). This demonstrates a strategy to reveal time-specific expression patterns missed at the population level despite the noise introduced by sampling conditions from subjects in the wild.

**Figure 3.**
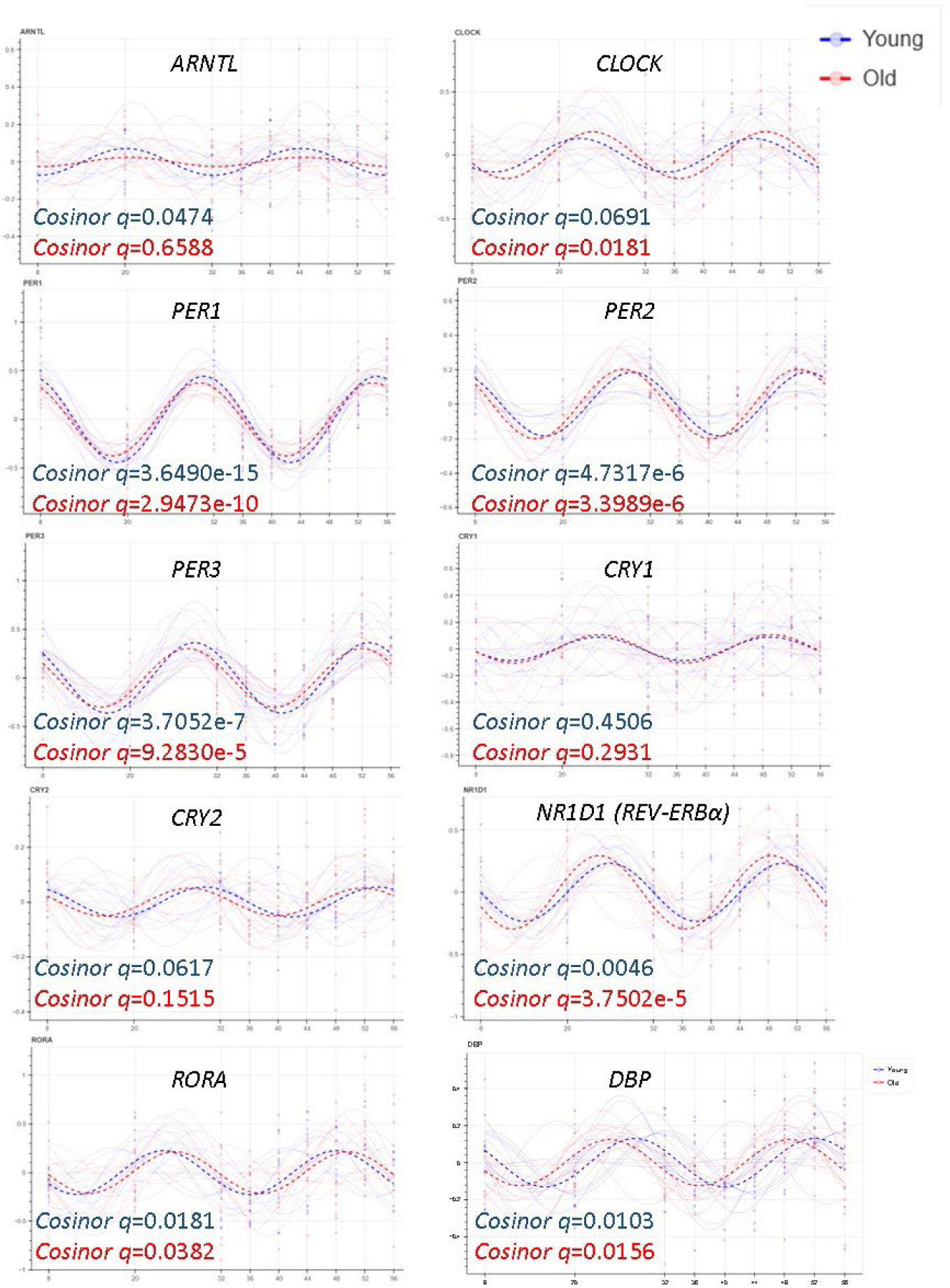
Diurnal gene expression patterns of molecular clock gene transcripts over 48 hours. Dashed bold lines represent population fits for young and old subjects while fine lines are the subject-level fits. Two cosinor q-values are provided for each transcript as significance measures for rhythmicity in the young and old cohorts. Please note that gene transcript abundances are MESOR-centered to enhance the comparison of oscillatory behavior between young and old.

The paucity of core clock proteins quantified in the AB- and MS-plasma proteome limited this assessment (*Supplementary Figure 3C, D*). Noticeably, we observed oscillatory trends for many methylated gene loci associated by proximity with core clock genes when relaxing FDR-corrected significance cutoffs (cosinor-*q*≤0.2, *Supplementary Figure 4*). Further functional mapping may elucidate the interplay between clock machinery and environmental factors.

### Diurnal Inflammaging Molecular Phenotypes in Young and Old

Inflammaging-associated proteins (4) circulate in plasma and their abundance is modulated by gene transcriptional activity and epigenetic regulation. Several of these AB-protein biomarkers oscillated in both young and old (SIRT1, IL-6, CDKN1A, *cosinor-q*≤0.05). Others showed no significant diurnal variation in either cohort (RETN, B2M, adiponectin, IL-10, *cosinor-q*>0.5, *Supplementary Figure 5*). Closer inspection of IL-6 showed varying phase-relationships between transcript and protein abundances (*Supplementary Figure 6*). KEGG pathway analysis of the AB-proteins did not identify cycling pathways that differentiated significantly between young and old (*cosinor-q*>0.05) as evident from their distribution in the p-value histogram (Supplementary Figure 7). Interestingly, 14 pathways cycled in young (*cosinor-q*=<0.05) but not in old (*cosinor-q*>0.05), while 9 pathways cycled exclusively in the old. Among the 14 were DNA damage repair pathways (base excision repair, mismatch repair), complement and coagulation cascades, protein digestion and absorption, and hedgehog signaling; among the 9 were breakdown and secretory processus (*Supplementary Table 4*).

Future characterization of time-specific functional networks relevant to inflammaging, senescence and premature aging may be achieved by examining oscillatory triads, which represent cycling features in the transcriptome, methylome and AB/MS-proteome converging on a single gene locus. These were detected in both young and old (*INPP5D, MAPKAPK2, NFE2, and PDLIM7*); only in the young (*CIRBP, DAPK2, DOK2, LRPAP1, NADK, NID1, PGD, and TALDO1*); and only in the old (*CD44, CMIP, FOXO1, HIP1R, ITGA6, NFATC1, NRP1, SMAD3,* and *ZBTB16*), (*Supplementary Table 5)*.

### Transomic integration of age-specific oscillatory features

We next sought to integrate the oscillatory features identified across the various omics datasets to identify candidates and associated pathways of potential functional relevance that differ between the two age groups. We confirmed that age-specific oscillatory differences were preserved in the transcriptome when the dimensionality of the omics datasets was reduced, while other omes showed less defined separation (*Supplementary Figure 8A, B, C*). However, when integrating data across omes, young and old clearly separated in their amplitudes (*Figure 4 d*). This is quite striking as the distinction was discernable in the absence of perturbation to test resilience, known to decline with age. The transomic analyses were accomplished by identifying the amplitudes of oscillatory features in each omic dataset, and simultaneously correlating features across modalities and age discrimination via the mixOmics platform (see methods, *Figure 4a,b*, *Supplementary Figure 9*).

**Figure 4.**
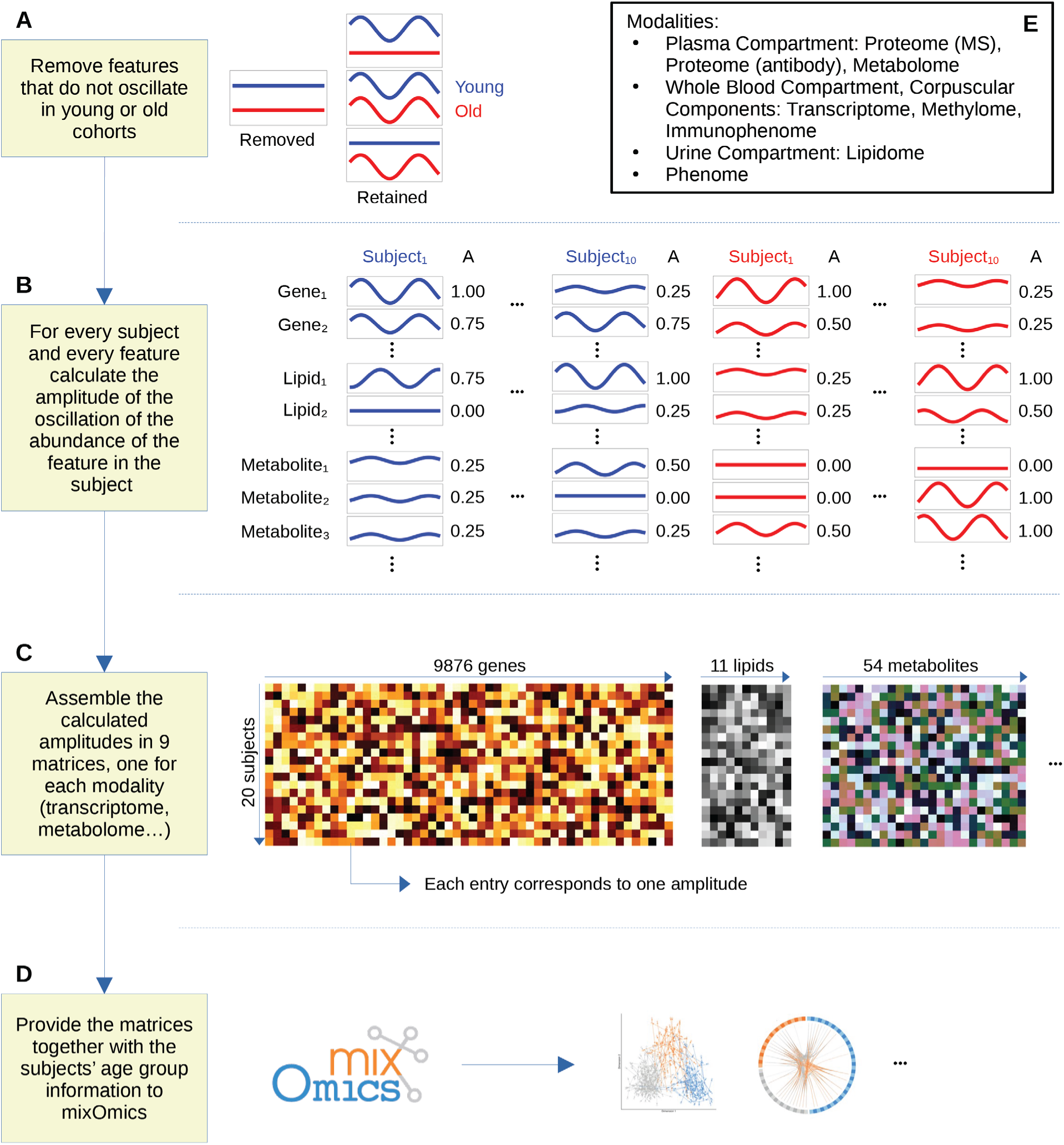
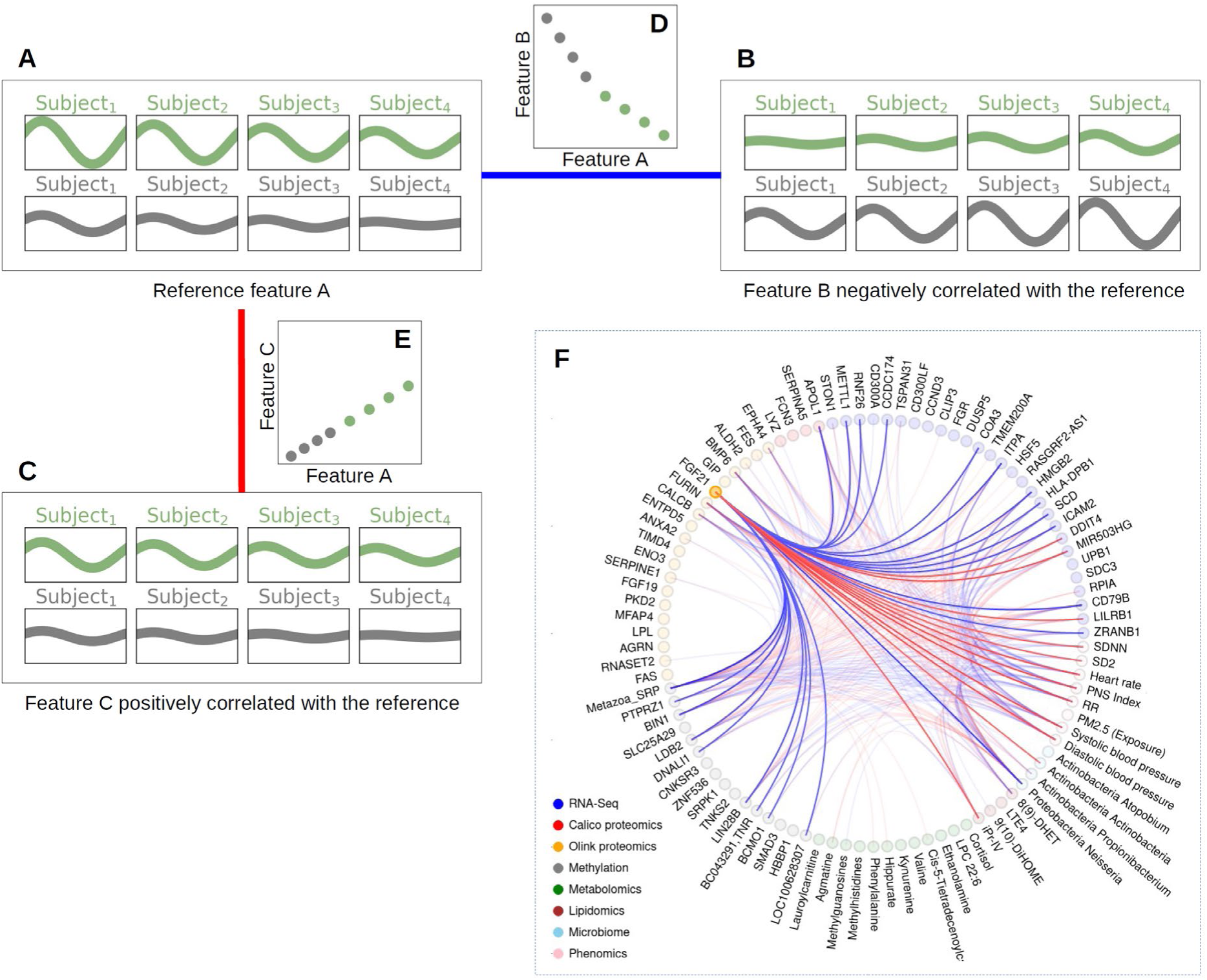
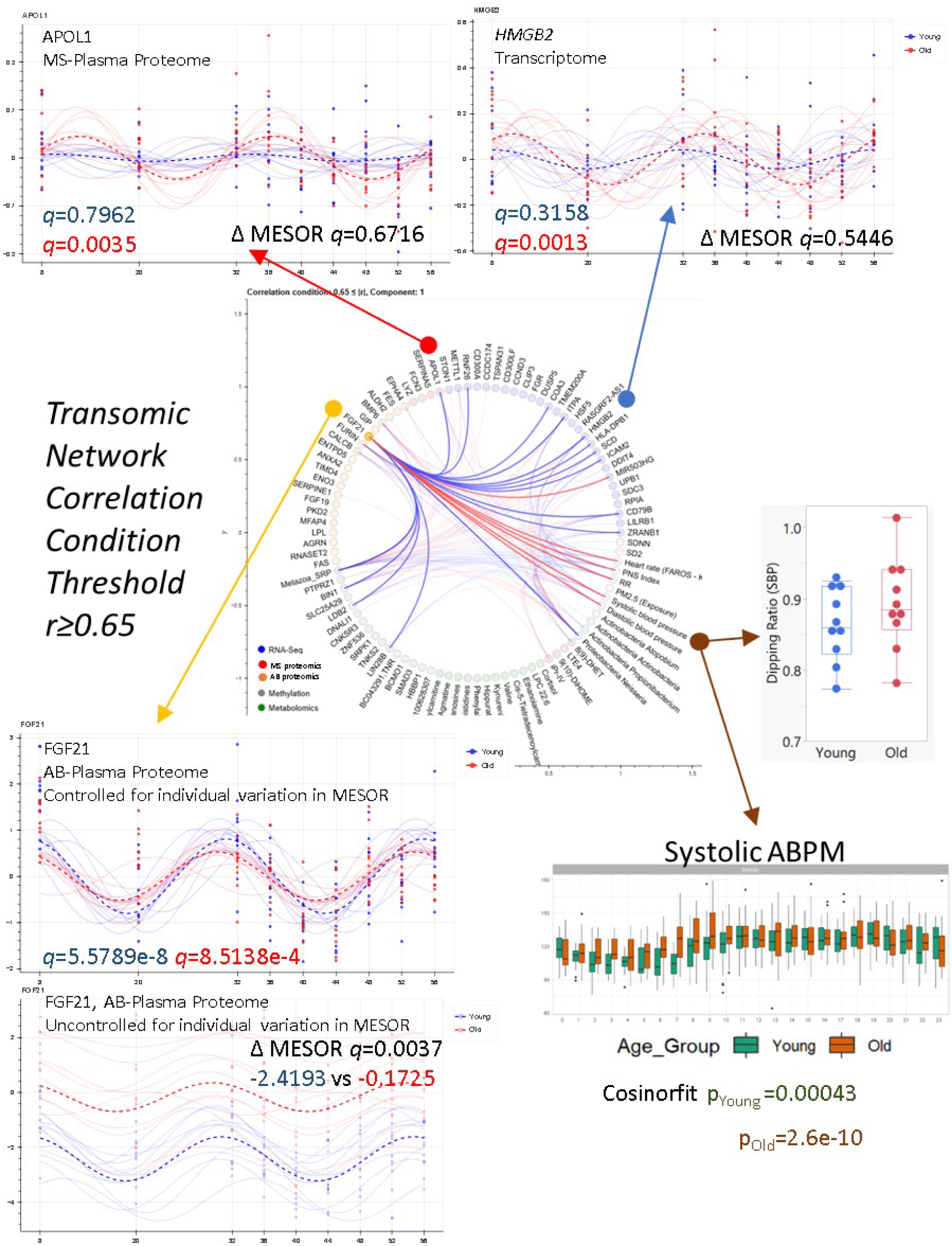
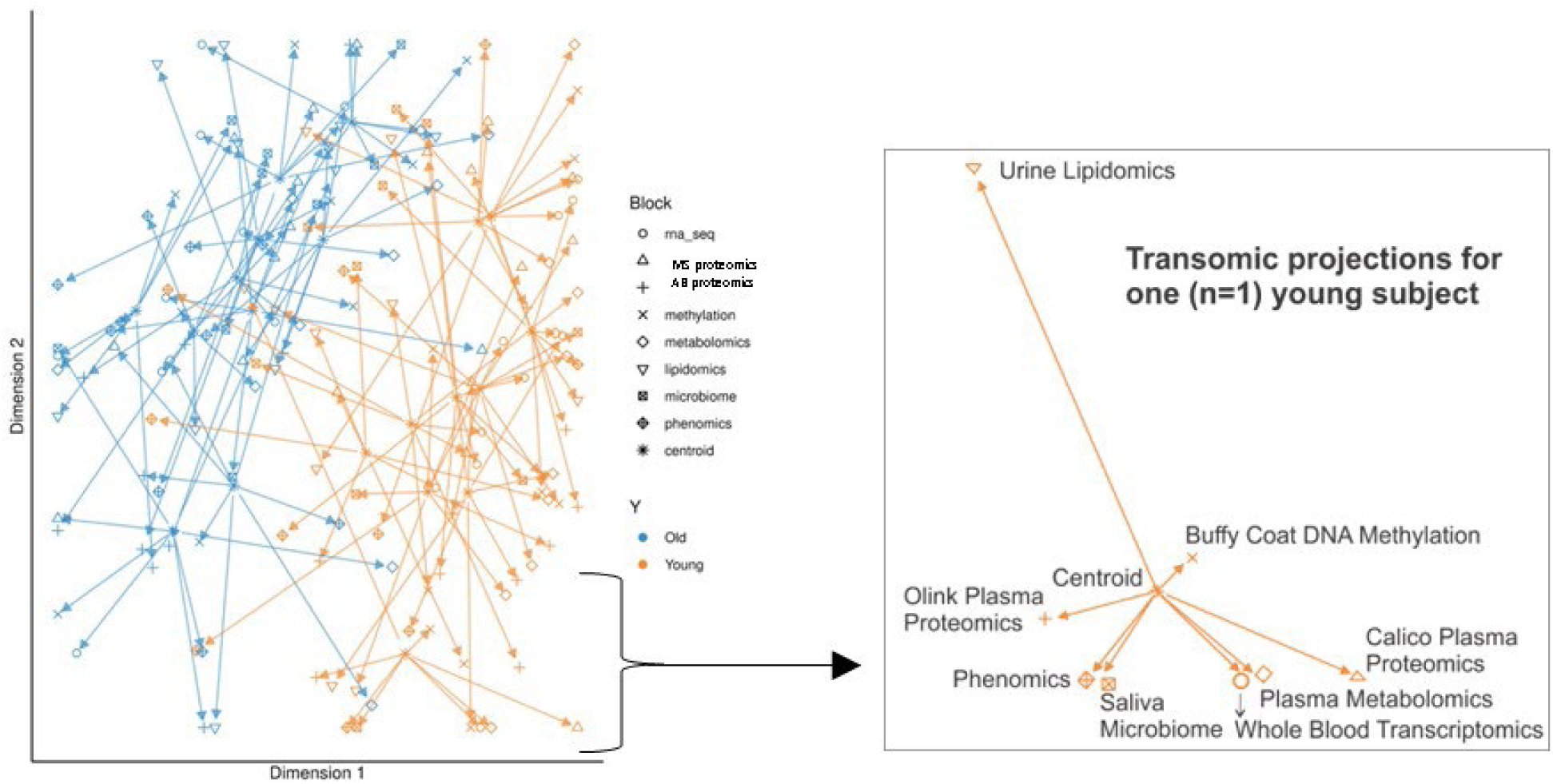
**a) Data preparation for integrative analysis of oscillatory patterns.** (A) Features that don’t exhibit circadian oscillation in young (q ≥ 0.1) and old (q ≥ 0.1) cohorts were removed since they would only contribute noise in this analysis. The calculation of cohort level q-values using cosinor fits is described in the Methods. (B) Amplitudes are calculated per feature and per subject using cosinor fits. MESOR and phase information are not used for this analysis. (C) Calculated amplitudes are assembled into matrices, one matrix per data type. (D) Matrices are provided to the mixOmics platform, along with the information about each subject’s age group (young or old). The output of the mixOmics analysis is discussed separately. (E) Modalities of data present in the integrative analysis. Protein data are separated into two modalities based on the assay used: mass-spectroscopy and antibody-based assay (Olink). **b) Circos plots representing the first integrated latent variables determined by mixOmics.** (A) Illustration of oscillatory patterns of a feature included in the Circos plot. Features included in the Circos plot distinguish between young (green) and old (grey) cohorts in terms of amplitude, i.e., the oscillation in one of the cohorts is dampened (or even absent) when compared to the other cohort. Reference feature A illustrates the case when the oscillations are stronger in the young cohort. Only four out of ten subjects in each cohort are being presented to conserve space. (B) Features with amplitudes of oscillation that are negatively correlated with the amplitudes of oscillation of feature A. Such pairs of features are connected by blue arcs in the Circos plot. (C) Features whose amplitudes of oscillation are positively correlated with the amplitudes of oscillation of feature A. Such pairs of features are connected by red arcs in the Circos plot. (D) Scatter plot depicting negative correlation of amplitudes of oscillation of features A and B. (E) Scatter plot depicting positive correlation of amplitudes of oscillation of features A and C. (F) Circos plot containing features in first latent variables with arcs connecting features with Pearson correlation coefficient (of amplitudes) lower than −0.6 (blue) or higher than 0.6 (red). Arcs connecting FGF21 to other features are rendered opaque, the remaining arcs exhibit a degree of transparency. c) Circos plot with Pearson correlation condition of r≥0.65 to examine transomic linkages between FGF21 (antibody proteome feature, lower left), APOL1 (mass-spec proteome feature, upper left), HMGB2 (transcriptome feature, upper right), and systolic blood pressure (phenome feature, lower right) including as reference the dipping ratio for systolic blood pressure (center right). Where applicable, q-values are provided for each feature as significance measures for rhythmicity in the young and old cohorts and the Δ mesor q-value indicates significance in the rhythm-adjusted means between young and old. d) Transomic projections derived from the molecular and phenomic features separate young and old in both dimensions on the group-level (left) with an example for subject-level transomic projections (right).

We sought oscillatory hub features to facilitate interpretation of the resulting cross-correlated omic and phenomic features, displayed in the Circos plot (features selected to optimize high correlations of amplitudes within and across modalities and whose amplitudes separate between young and old cohorts). These hubs show significant diurnal variability (in either young or old) and display high cross-correlations with most features from other modalities. At a relaxed stringency level (Pearson correlation condition of |r|≥0.55 in the Circos plot), these oscillatory hub features are predominantly present in the phenome (8 out of 8 features) and the transcriptome (17 out of 28), while only half of the AB-proteins display a high degree of transomic connectedness (10 out of 20) and much lower numbers for the metabolites (2 out of 12 features). At the most stringent level of cutoff (Pearson correlation condition of |r|≥0.8), FGF21 protein abundance, as measured by the AB-platform, emerged as a prominent oscillatory hub (Supplementary Figure 10). To examine a broader sample of transomic linkage across multiple features, we balanced the stringency at midlevel (Pearson correlation condition of |r|≥0.65) and observed that FGF21 cross-correlated with several features in the transcriptome (n=10), phenome (n=5), methylome (n=4), saliva microbiome (n=1) and MS proteome (n=1) (*Figure 4c*). Here, a majority of these transomic linkages (n=15), based on the flowchart in *Figure 4b*, indicated an age-dependent inversion of oscillatory patterns (negative cross-correlations, blue arcs in *Figure 4c*) while preserved oscillatory patterns (positive cross-correlations, red arcs in *Figure 4c*) were evident in a smaller number of molecular features (n=6) as well as in the phenomic features related to blood pressure and heart rate variability (n=5). The age-dependent inversion of transomic oscillatory patterns is dominated by pairs of features (FGF21 is the reference feature) with detectable oscillations in the old but not the young (*Supplementary Figure 11*). FGF21 protein abundance oscillated in both groups but the amplitude was greater in the young; it negatively correlated with the abundance of APOL1 MS protein and HMGB2 gene transcript. Thus, the subject-level amplitudes of FGF21 oscillation negatively correlated with the amplitudes of these two proteins, both of which oscillated in the old, but not the young.

Several other oscillatory hub features emerged at the Pearson correlation condition of |r|≥0.8, with FURIN (AB-protein), Metazoa-SRP, 8(9)-DHET, Proteobacteria Neisseria, systolic and diastolic blood pressure and PNS index (*Supplementary Figure 10*). Relaxing the Pearson correlation condition to |r|≥0.7, adds BMP6 (AB-protein), ITPA, HMGB2, ICAM2, CD79B, ZRANB1, heart rate, and RR to the list of candidate oscillatory hub features (*Supplementary Figure 12*). We discuss these in more detail in the supplementary data. Several features, however, remain islands without transomic cross-correlations even at the Pearson correlation condition of |r|≥0.55. These include, for example, GIP, ENTPD5, TIMD4, ENO3, FGF19, LPL, AGRN, and FAS in the AB proteome (*Supplementary Figure 13*).

The strength of our integrative approach is that we produce a candidate list of oscillatory hub features which show substantial diurnal variability (in either young or old) and are highly correlated with features across the chronobiome that distinguish between young and old. This sets the stage to validate experimentally the functional contribution of these oscillatory hub features to physiological aging in follow-up studies.

### Transcriptomic integration of the chronobiome with the Genotype-Tissue Expression (GTEx) Project

The Adult Genotype-Tissue Expression (GTEx) Project comprehensively quantified gene expression across 54 non-diseased tissue from about 1,000 deceased donors ^12^. In a subcohort of 914 donors (grouped as <50 vs >60 years of age), a previous study detected oscillatory gene expression patterns from 46 human tissues, and observed that circadian clock genes maintained their temporal relationships across tissues ^2^. We determined the extent of overlapping oscillatory transcripts between deceased GTEx donors and living chronobiome volunteers. We found that transcripts oscillating in the plasma of our aging chronobiome volunteers (*q*<0.01) aligned with the cycling transcripts detected in 26 different donor tissues of the deceased GTEx patients (*q*<0.01, *Supplementary Figure 14 A*), with the highest overlap observed in body fat. Specifically, we detected n=172 oscillatory transcripts in the young chronobiome which shared their diurnal phenotype with the expression rhythms in subcutaneous adipose GTEx tissue. This overlap was more pronounced for the old chronobiome, where n=246 shared the rhythmic gene expression. Oscillatory patterns in several core clock genes (*PER2, PER3, NR1D1, NR1D2*) and clock-related genes (*TEF, FKBP5)* showed substantial concordance between GTEx and the chronobiome of both young and old (*Supplementary Figure 14 B top*). Other transcripts showed an age-specific oscillatory overlap only between the young chronobiome and GTEx (*DDIT4* and *CCND3*, *q*<0.01) or between the old chronobiome and GTEx (*PER1*, *q*<0.01).

Similarly high numbers of overlapping oscillatory transcripts between the plasma chronobiome and GTEx were apparent in tissues from the omental visceral adipose (263 young, n=190 old), tibial nerve (n=164 young, n=205 old), and skeletal muscle (n=92 young, n=285 old) specifically for several core clock genes (*PER* and *NR1D1* homologs) and clock-related genes (*TEF, FKBP5) (*Supplementary Figure *14 B, C)*.

Little overlap (n<10 oscillatory transcripts) was evident, as expected, for GTEx tissues spatially removed from plasma, such as vagina, prostate, and the different brain regions (Supplementary Figure *14 A magnification*). For GTEx liver tissue the overlap was skewed towards cycling transcripts from the chronobiome in the old (*Supplementary Figure 14 D top)*. Here, many of these features have been associated in the literature with modulating the risk of hepatocellular carcinoma (HCC), such as *STIP* ^13,14^, *PPARD* ^15^, *PHB2* ^16^, *NDRG2* ^17^, *HDAC6* ^18^, *OCIAD2* ^19^, and *DCXR* ^20^, though future studies are necessary to understand the age-specific relationship to diurnal variability. Interestingly, an age-specific enrichment of n=10 cycling transcripts was evident in the cerebellum where clock genes (*PER* and *NR1D1* homologs) and several genes associated with neurological phenotypes (*TSC22D1* ^21^, *IRF2BPL* ^22^, *DNAJB5* ^23^) overlapped only with the old chronobiome (*Supplementary Figure 14 D bottom)*.

### Proteomic integration of the chronobiome with the UK Biobank (UKBB)

The same Olink proximity extension assays were applied to proteomic analysis in both this study and the UKBB. Leveraging this convergence with published data ^6,7^, we found that age dependent differences in protein abundance aligned between the two data sets. Thus, the 85 proteins that differed significantly in abundance (MESOR) with age in the chronobiome aligned well with those that differed with age in the UKBB ^6^. This significant overlap (Pearson correlation coefficient *r*=0.58) included proteins more abundant in young compared to old (PDXL2, FLT3, LTA, BOC, CD1C, PTPRR, PRL, CTSV, LPO, CR2 and PLB1) and proteins more abundant in old compared to young (GDF15, LTBP2, HSPB6, ACTA2, CDCP1, ELN, MLN, GIP, LMOD1, DTX3, EDA2R, NEFL, WFDC2, MSR1, WNT9A, EFHD1, NT5C1A, PRELP, *Figure 5* A).

**Figure 5.**
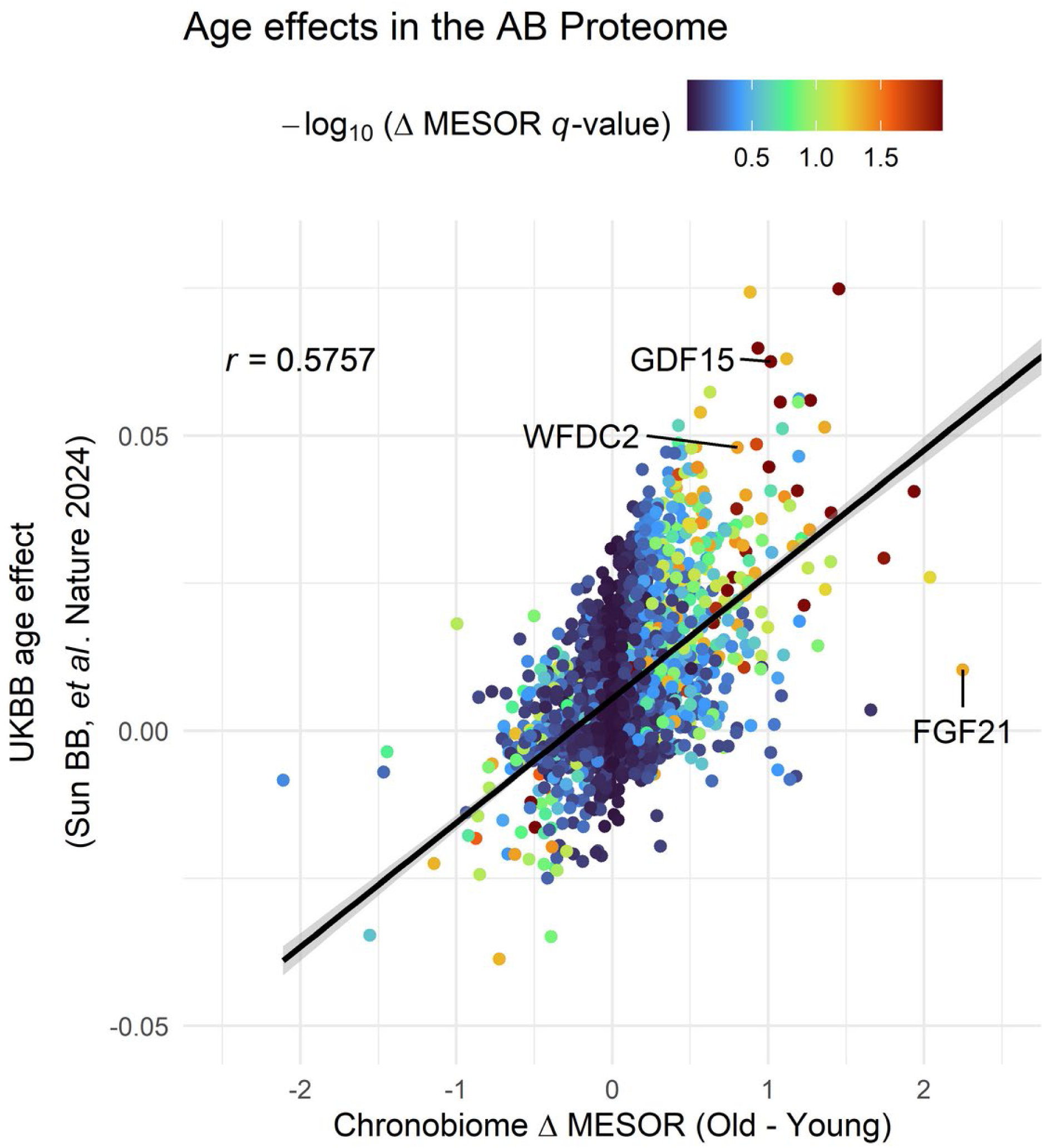
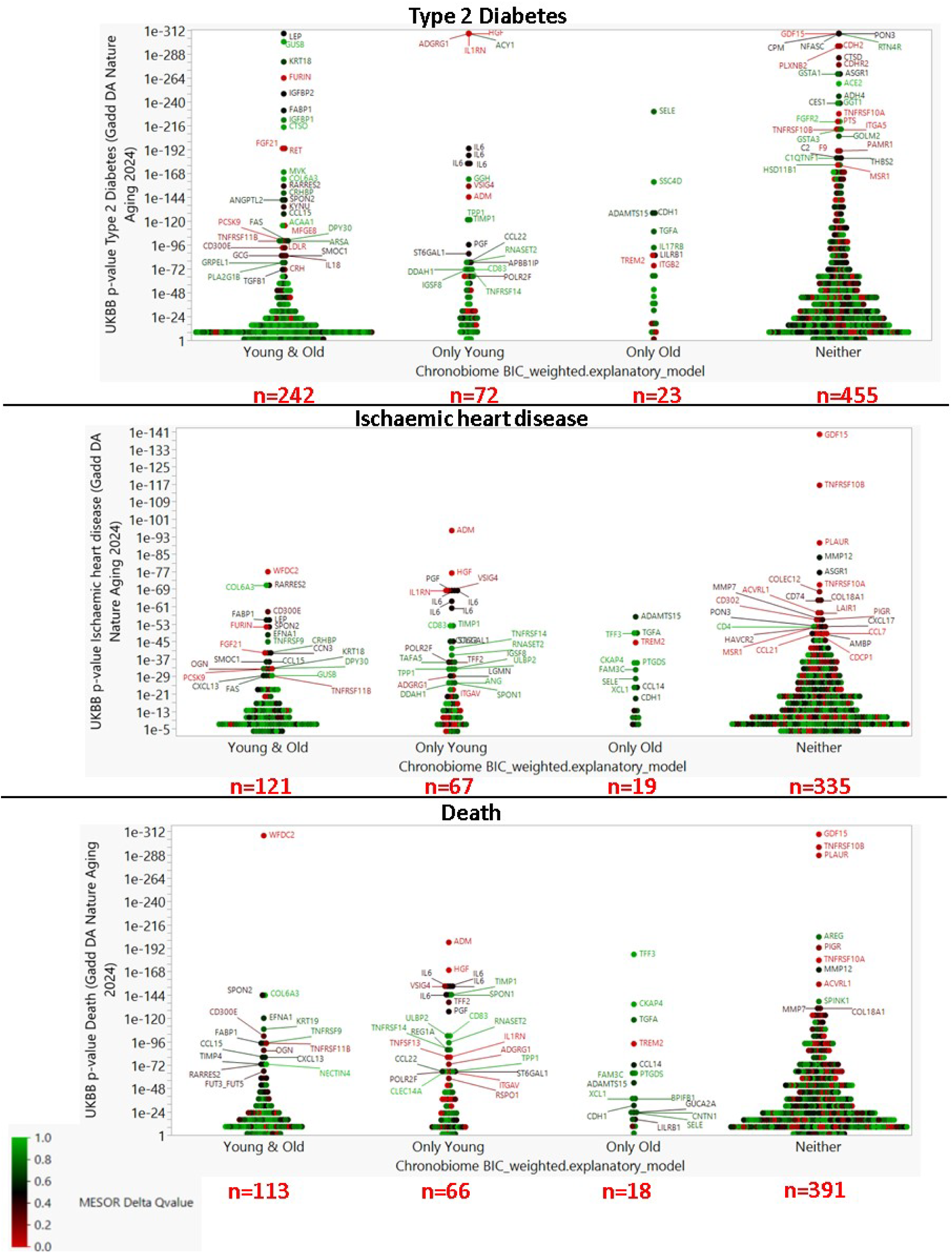
(A) The age-specific difference in AB-protein abundance in the chronobiome correlates with the age-specific AB-protein abundance in the UKBB as reported in the Extended Data Figure 3a and Supplementary Table 5 of ^6^.The effect size of the age-dependent difference in the chronobiome is color-coded using the −log_10_ (delta MESOR *q*-values). Age predictors, such as GDF15, align in this matrix as expected. (B) The overlap between the cycling AB-proteome (four categories on the x-axis: cycling in young and old, cycling in only the young, cycling in only the old, and cycling in neither) as determined by the chronobiome and the protein-disease association (y-axis) as determined by the UKBB ^7^ is shown for (top) type 2 diabetes, (center) ischemic heart disease, and (bottom) death. All proteins shown satisfy the following cut-off values for significance: chronobiome BIC>0.75, UKBB Bonferroni-adjusted p<3.1e-6. The sample sizes located under each graph indicate the number of cycling proteins in each category. The color overlay indicates the chronobiome mesor delta q-value which highlights in red the proteins with an age-specific difference in abundance in the cycling and non-cycling proteome. Note, some features (like IL6) are present in each of the inflammation, neurology, oncology and cardiometabolic Olink panels. These features appear multiple times (once for each panel) in the plots.

Furthermore, 796 proteins associated in the UKBB with 10 common disease outcomes and mortality ^7^ (Bonferroni-adjusted *P* value threshold =3.1×10^-6^) were cyclers in the chronobiome in both young and old (BIC>0.75), while 433 predictive features were cyclers only in the young compared to 129 cycling features in the old chronobiome observing the same stringent statistical thresholds. A total of 2,506 features predictive of UKBB outcomes were characterized as neither oscillating in the young nor old chronobiome (*Supplementary Table 6*).

Overall, the disease related proteins in the UKBB were more likely to be nonrhythmic in the old compared to the young chronobiome, evident, for example, in type 2 diabetes where 32 features cycling in old compared to 72 in young. This pattern held across a range of proteins predictive of age-related diseases. These included ischemic heart disease, death, liver disease, Parkinson’s disease, COPD, ischemic stroke), rheumatoid arthritis, lung cancer, and systemic lupus erythematosus (*Figure 5 B* and *Supplementary Figure 15 A-G*).

Amongst the disease related proteins in the UKBB that lose their oscillation in the chronobiome with age are those reflective of immune response programs. For example, IL6 is predictive in the UKBB for type 2 diabetes, ischemic heart disease, liver disease, death, COPD, ischemic stroke, rheumatoid arthritis, lung cancer, and systemic lupus erythematosus (*Supplementary Figure 15*).

### The chronobiome dashboard on the World Wide Web

The multiomics data is available at https://chronobiome.org/, which offers an interactive platform to explore time- and age-specific characteristics of molecular features across datasets with statistics derived from fitting a mixed linear model. Representative screenshots are provided in *Supplementary Figure 16* to facilitate navigation. [*This resource becomes accessible once the peer review process concludes.*]

## Discussion

This study compares the chronobiome in apparently healthy younger versus older individuals as a first step towards addressing the hypothesis that deconsolidation of oscillatory rhythms may contribute to age dependent emergence of disease. We identified oscillatory features in both multiomic and behavioral data in both groups. The study participants were distinct in both chronological and biological age. Differences in physiological features – grip strength, auditory responsiveness, blood pressure – were apparent, but within the physiological range. Integrating the individual multiomics data using the mixOmics platform afforded a clear distinction between the younger and older group, even in the absence of a test of resilience ^24^. The failure to detect interactions between age and time of day within distinct omic datasets is likely reflective of a lack of power.

Age-dependent functional insights may be gleaned from oscillatory triads, which integrate cycling features in the transcriptome, methylome and proteome converging on a single gene locus. CIRBP, for example, has a role in recovery from sleep deprivation ^25^, so that the disintegration of an oscillatory CIRBP triad in the old may suggest a higher susceptibility to sleep fragmentation in the old compared to the young. In contrast, the switch to an oscillatory triad for CD44 and NRP1 in the old compared to the young may indicate time-of-day-dependent loss of endothelial integrity in the vasculature ^26,27^.

To identify changes in oscillatory features of potential functional relevance to the differences between the groups, we conducted multivariate mixOmics analyses of features where oscillation differed (present/absent or dampened) by age and sought their interactions. As a sanity check, we applied the most stringent statistical correlation conditions to reveal the first set of such relationships. This was the negative correlation in amplitude of FGF21 protein abundance with pro-inflammatory features as well as its positive correlation in amplitude with blood pressure and heart rate. FGF21, a protein generated by the liver, pancreatic beta cells, adipose tissue and skeletal muscle, has been related to beneficial metabolic effects on thermoregulatory functions, glucose and lipid metabolism ^28^ and has been positively correlated with lifespan ^29^. APOL1 protein abundance and HMBG2 gene transcript abundance, both negatively correlated with FGF21 protein abundance, have been associated with inflammation. APOL1 is known to activate the inflammasome by binding NLRP3 with caspase-1 as a host defense mechanism which plays a role in hypertensive kidney diseases ^30^. Of interest for future interrogation is how oscillatory transomic relationships of APOL1 are modulated by high-risk APOL1 genotypes associated in Black individuals with end-stage kidney disease ^31^. HMGB2 has been shown to orchestrate cellular senescence and is required for AIM2 inflammasome activation ^32^. FGF21 has been shown to inhibit directly the NLRP3 inflammasome ^33^. Thus, these inter-relationships raise the possibility that erosion of the mitigating actions of FGF21 on age related inflammation is a candidate oscillatory pathway that discriminates between healthy younger and older subjects. Correspondingly, the positive correlation between FGF21 and systolic blood pressure and other cardiovascular parameters is consistent with its previously reported correlation with blood pressure and its renoprotective capabilities ^34^. In the literature, this is consistent with the mitigating effect of FGF21 on the inflammatory features of aging ^29,35^ and its association with cardiovascular outcomes ^34^.

Characterization of the age related human chronobiome affords an opportunity complementary to human genetics, forced desynchrony protocols and postmortem data in the study of human circadian biology. Although the differences with age amongst these healthy individuals are modest, the integrated transomic data both permit segregation of the age groups and identification of a candidate mechanism (age dependent erosion of FGF21 dependent mitigation of inflammation) by which oscillatory function might contribute to age dependent distinctions between these apparently healthy groups.

While the sample size of the chronobiome is small, the participants have been phenotyped at a depth unprecedented in studies of human chronobiology. This creates a unique resource that can be interrogated by the community (https://chronobiome.org/). Furthermore, the integration of such cross-sectional studies with data at scale, such as GTEx and UKBB, is an opportunity to derive novel biological insights. While statistical methodology for longitudinal / cross-section data integration is still evolving, we show that subsets of proteins predictive of disease in the UKBB that differ in abundance with age align nicely with age dependent differences in the chronobiome. For example, disease and death related proteins in the UKBB more likely oscillate in the young than in the old in the chronobiome. Furthermore, many transcripts and proteins that displayed oscillatory behavior in this study, also showed time-of-day variation in GTEx. Age related changes of cycling in disease proteins in the chronobiome align with changes of protein oscillation coincident with disease in the UKBB described in the accompanying paper ^36^. This is consistent with the hypothesis that progressive circadian disruption predicts the emergence of age-related disease. The longitudinal acquisition of biosamples and phenotypic information planned for the UKBB and other biobanks will allow this hypothesis to be tested further.

## Data Availability Statement

This study is registered with dbGap under accession number [pending]. dbGap provides secure storage of and controlled access to: Anonymized participant metadata, raw RNA-Seq FASTQ files, DNA methylation array data, antibody plasma proteomics, urine metabolome and saliva microbiome data.

Anonymized metabolomics data were deposited under accession number [pending] in MetaboLights (EMBL-EBI) and are available at the following URL [pending]. Anonymized mass spectrometry proteomics data were deposited to the ProteomeXchange Consortium (http://proteomecentral.proteomexchange.org) via the PRIDE partner repository ^37^ with the dataset identifier PXDxxxxxx [pending].

Statistics and fits to ascertain oscillatory characteristics of the anonymized molecular data collected are available via the Chronobiome online data portal located at https://chronobiome.org/.

## Code Availability

The codes used for the analysis were deposited at Github under the URL [pending].

## Acknowledgments

We are indebted to the volunteer participants of this study who supported our efforts with their engagement and feedback. We thank the staff of ITMAT’s Center for Human Phenomic Science (CHPS) who adapted to our research needs to implement this comprehensive chronobiome research. C.S. is the Robert L. McNeil Jr. Fellow in Translational Medicine and Therapeutics. G.A.F. holds the McNeil Professor of Translational Medicine and Therapeutics.

We like to thank the following investigators for facilitating the adoption of the TracMyAir Smartphone Application for monitoring air quality: Michael Breen, PhD, from the Center for Public Health and Environmental Assessment, U.S. Environmental Protection Agency, Research Triangle Park, North Carolina, 27711, USA; Catherine Seppanen,, PhD, from the Institute for the Environment, University of North Carolina at Chapel Hill, Chapel Hill, NC 27514, USA; Vlad Isakov, PhD, and Sarav Arunachalam, PhD, from the Center for Environmental Measurement and Modeling, U.S. Environmental Protection Agency, Research Triangle Park, North Carolina, 27711, USA.

We acknowledge Georgios K. Paschos, PhD, for helpful discussions and LaVenia Banas, RN, for excellent clinical study logistics and subject recruitment.

