## Supplemental Material for "Age and the Diurnal Oscillatory Features of the Human Chronobiome"

*Corresponding Authors:

**The PDF file includes:**

Materials and Methods
Supplementary Tables 1 to 6
Supplementary Figures 1 to 16
Supplementary References

#### Materials and Methods

##### *Study Subjects, Study Design*

Apparent healthy young (18-30 years of age) and old (55-75 years of age) volunteers with a BMI ≤27.4 and who owned a smartphone were enrolled into this study (University of Pennsylvania IRB# 834156, NCT04225442). Exclusion criteria included smoking, shift work, transmeridian travel across ≥3 time zones in the two weeks prior to the start of study activities and a history of severe psychiatric illness or cognitive conditions, clinically significant obstructive sleep apnea, diabetes mellitus, gastroesophageal reflux disease, bilateral mastectomy, or Raynaud's phenomenon. Participants with serum creatinine > 1.5 mg/dl in men or >1.3 mg/del in women, significant liver disease (>3x upper limit of normal), use of oral or intravenous antibiotics in the 6 months prior to study activities, received an experimental drug, used an experimental medical device within 30 days prior to screening, or who gave a blood donation of ≥ one pint within 8 weeks prior to screening, > 2 drinks of alcohol per day, positive urine test for illicit drugs, known allergy against natural latex rubber, use of alpha-blockers, or nursing or pregnant were excluded. Safety was determined through blood laboratory screens, pregnancy tests, screen test for substance abuse, and documentation of adverse events.

Study assessments occurred under physiological conditions in the wild with sparse biosampling in the outpatient setting on day 1 (12-hour collection time intervals at 08:00 and 20:00), followed by frequent biosampling (4-hour collection time intervals at 08:00, 12:00, 16:00, and 20:00 on day 2 followed by 24:00, 04:00 and 08:00 on day 3) in the inpatient setting of the Center for Human Phenomic Science (CHPS), Institute for Translational Medicine and Therapeutics (ITMAT) at the University of Pennsylvania. Biosamples were processed after each collection time point and stored at -80°C until further analyses.

During the biosampling over 48 hours, grip strength (Jamar Smart Hand Dynamometer, Chicago, IL, US; data collection standardized by means of the NIH Toolbox) and cognitive function (Penn Computerized Neurocognitive Battery, CNB [1], University of Pennsylvania, Philadelphia, PA, US) were assessed in the mornings and evenings (12-hour intervals). Synchronous continuous assessments through wearable devices included accelerometry

(ActiGraph wGT3X-BT, Actigraph, Pensacola, FL, US), ECG monitoring (Bittium Faros 180, Bittium Corporation, Oulu, Finland) and smartphone-based digital phenotyping (Beiwe, Harvard University). Ambulatory blood pressure monitoring (Spacelab 90207, Spacelabs Healthcare, Snoqualmie, WA, US) was deployed asynchronous to guard the biosampling period against sleep disruption induced by the blood pressure cuff, as was the smartphone app TracMyAir (Environmental Protection Agency, US) [2] and PurpleAir monitor (PurpleAir Inc., Draper, UT, US) to determine hourly PM2.5 and O3 environmental exposure metrics. Surveys included standard health questionnaire, the Munich Chronotype Questionnaire (MCTQ) [3], and the Big Five Inventory (BFI).

Study data were collected and managed using REDCap electronic data capture tools hosted at the University of Pennsylvania [4,5]. REDCap (Research Electronic Data Capture) is a secure, web-based software platform designed to support data capture for research studies, providing 1) an intuitive interface for validated data capture; 2) audit trails for tracking data manipulation and export procedures; 3) automated export procedures for seamless data downloads to common statistical packages; and 4) procedures for data integration and interoperability with external sources.

##### *Whole Blood RNA-Seq*

Aliquots of blood were collected from study participants and stored in PAXgene Blood RNA Tubes (BD Biosciences; Franklin Lakes, New Jersey). Total RNA was extracted from PAXgene blood aliquots using the Maxwell RSC simplyRNA Blood Kit (Promega; Madison, Wisconsin). Purified RNA samples were assessed for quality using a TapeStation System (Agilent Technologies; Santa Clara, California). All extracted RNA samples had RNA Integrity Number (RIN) values greater than or equal to seven. Total RNA was prepared for Illumina sequencing using the TruSeq Stranded mRNA library preparation kit (Illumina; San Diego, California). Libraries were prepared in two batches because Illumina adapters were only available in plates of 96 unique barcodes. The completed libraries were sequenced in a 2x100 bp, paired end configuration on a NovaSeq 6000 Xp workflow, with a v1.5 S4 flowcell (Illumina). All RNA extractions, library preparations, and sequencing were performed according to the manufacturers' protocols by the Center for Applied Genomics (CAG) Sequencing Core at the Children's Hospital of Philadelphia (Philadelphia, Pennsylvania). The samples were provided to the CAG with randomized, blinded identifiers to prevent any biases or batching in sample preparation that were confounded by participant, collection time, or age cohort.

Raw sequencing reads were aligned to the GRCh38 build of the human reference genome using v2.7.6a of STAR [6]. These STAR alignments were run with the following command line parameters: '--outSAMtype BAM Unsorted --outSAMunmapped Within KeepPairs --outFilterMismatchNmax 33 --seedSearchStartLmax 33 --alignSJoverhangMin 8'. Next, the data were normalized and quantified at the gene-level using v0.8.5e-beta of the Pipeline Of RNA-Seq Transformations (PORT; https://github.com/itmat/Normalization). Both STAR and PORT were provided with gene models from v104 of the Ensembl reference annotation [7]. All downstream analyses were performed using the PORT-normalized, gene-level read counts.

***Antibody plasma proteomics***

EDTA plasma, 80 uL per biosample, was aliquoted in-house to 96 well plate in randomized order, blinded and shipped on dry ice for analysis of Olink® Explore 1536 and Olink® Explore Expansion to Olink Proteomics, Waltham, MA, US.

***LC-MS plasma proteomics***

EDTA plasma biosamples were shipped on dry ice to Calico Labs, San Francisco, CA.

**LC–MS Sample Preparation for human plasma**

Plasma samples were prepared using an in-house automated multiplexed proteome profiling platform (AutoMP3) protocol [8] with some minor modifications. In brief, 15 µL of human plasma was diluted with 285 µL of SDS lysis buffer (3% SDS, 50 mM HEPES (pH 8.5), 75 mM NaCl, protease inhibitor (cOmplete ULTRA, Roche), and phosphatase (PhosSTOP, Roche) inhibitor tablet. 40 µL of the diluted plasma was then taken out for further processing. Proteins were reduced with dithiothreitol (DTT, 5 mM, 37 °C for 30 min) and then alkylated with iodoacetamide (15 mM, room temperature for 20 min in the dark). DTT was then used to quench excess iodoacetamide (5 mM, room temperature for

15 min in the dark). Protein cleanup was performed using a 1:1 mixture of E3:E7 Sera-Mag Carboxylate-Modified Magnetic Particles (Cytiva Life Sciences, Marlborough, MA, USA). In brief, the beads were first washed with water 3 times, and then the sample was added onto the beads. For protein binding, acetonitrile was added to a concentration of 75%, and the mixture was incubated for 20 min at room temperature without shaking. The beads were then washed twice with 70% ethanol, followed by 2 washes with 100% acetonitrile. The beads were resuspended in digestion buffer (100 mM EPPS, pH 8.5, 10 mM $CaCl_2$), and then Trypsin/LysC (Promega) was added at a ratio of 1:25 (enzyme: substrate), followed by incubation for 1 h at 37 °C (1000 rpm) to digest the proteins. The peptide digests were stored at -80℃ until further preparation.

For all digested plasma samples,18 µL (~20 µg digest) was taken out and mixed with 62 µL of digestion buffer (100 mM EPPS, 10 mM $CaCl_2$) for a final volume of 80 µL which was used for TMT-labeling with 24 µL of TMTpro reagents (0.2 mg/ml, in acetonitrile). In addition, a pool consisting of equal volumes from all the individual samples in the study was made. This pool was labeled with the TMTpro126-tag and then acted as bridge samples in all the plexes. All samples were incubated at 25 °C for one hour after the addition of the TMTpro-tags. Excess TMT was quenched by adding 11 µL 5% hydroxylamine solution before they were combined. The combined samples were then split into multiple SP3 peptide cleanup reactions due to % organic solution constraints. In brief, 50 µL prewashed beads were mixed with 50 µL of the combined sample, and then the solution was immediately brought to 95% isopropanol by addition of 100% isopropanol. Samples were incubated at room temperature for 18 minutes to enable peptide binding to the beads. Tubes were then placed on the magnet for 2 min, and the supernatant removed followed by two rounds of washes with 95% isopropanol, and then one with 100% acetonitrile. The peptides were eluted in three fractions using 80% acetonitrile, 50% acetonitrile and finally 5 % acetonitrile. The cleaned-up peptides were dried down in a speedvac (Labconco) and finally reconstituted in 5% formic acid, 5% acetonitrile and 1 µg was injected for analysis.

**TMT Data Acquisition**

All data were obtained on an Orbitrap Eclipse mass spectrometer with FAIMS (Field Asymmetric Ion Mobility Spectrometry) Pro Interface. The mass spectrometer was coupled to an UltiMate 3000 HPLC operating in DDA mode (Thermo Fisher Scientific). Peptides were separated on an Aurora Series emitter column (25 cm × 75 µm i.d., 1.6 µm, 120 Å pore size, C18; IonOpticks) using a 165 min gradient from 8 to 30% acetonitrile in 0.125% formic acid. FAIMS was enabled during data acquisition with compensation voltages set as −40, −50, −60 and -70 and each voltage had a cycle time of 1.25 s. A high-resolution MS1 scan was performed in the Orbitrap at a 120,000 resolving power, m/z range 400–1600, RF lens 30%, standard AGC target, and “Auto” max. injection time. The parameters for MS2 acquisition were as follows: 0.7 m/z isolation window, CID, 35% normalized collision energy with a CID activation time of 10 ms, AGC $1 \times 10^4$, 35 ms max. injection time). Real-time search (RTS) [9] with the Uniprot database UP000005640 was used for data collection. Depending on the RTS, the ions were then analyzed with SPS MS3. For the RTS, the minimum Xcorr needed to pass was set to 1, the minimum dCn was set to 0.1, and the maximum missed cleavages allowed was set to two. MS3 analysis was performed in the Orbitrap with the following parameters: HCD, 50 k resolution, 45% normalized collision energy, AGC $1 \times 10^5$, m/z range 100–500 m/z, and 200 ms max. injection time. A maximum of 10 fragment ions from each MS2 spectrum were selected for MS3 analysis using SPS; 10 ppm was set as the mass tolerance for the target masses.

**Data Analysis**

An in-house software pipeline (version 3.12) was used to process all mass spectrometry data [10]. Raw files were first converted to mzXML files using the MSConvert program (version 3.0.45) to generate peak lists from the RAW data files, and spectra were then assigned to peptides using the SEQUEST (version 28.12) algorithm [11]. Spectra were queried against a “target-decoy” protein sequence database consisting of human proteins (Uniprot database UP000005640 containing 75776 features and common contaminants) in forward and reversed decoys of the above [12]. The parent mass error tolerance was set to 20 ppm and the fragment mass error tolerance to 0.6 Da. Trypsin specificity was required allowing for up to two missed cleavages. Carbamidomethylation of cysteine (+57.02 Da), TMTpro-labeled N terminus and lysine (+304.20 Da) were set as static modifications. Methionine oxidation (+15.99 Da) was set as variable modification. Following database searching, linear discriminant analysis (LDA) was performed to filter peptide spectral matches to a 1% false discovery rate (FDR) [10]. Following peptide filtering, non-unique peptides were assigned to proteins that comprised the largest number of matched redundant peptide sequences using the principle of Occam's razor. The quantification of TMT reporter ions was performed by extracting the most intense ion at the predicted m/z value for each reporter ion (within a 0.003 m/z window) and isotopic purity correction of reporter quant values was applied. Known false positives (i.e., decoys and contaminants) were excluded from further analysis steps and peptide intensities and signal-to-noise ratios were exported for further analysis and subsequently analyzed using the msTrawler statistical software package [13]. Protein fold changes were exported from msTrawler and then further processed using R and Nitecap [14].

**Data Availability Statement**

The mass spectrometry proteomics data have been deposited to the ProteomeXchange Consortium via the PRIDE [15] partner repository with the dataset identifier PXD-TBD.

***Buffy Coat Methylome***

Whole blood buffy coat samples were submitted to blood DNA extraction method of Agencourt GenFind V2 Protocol (Protocol 001072v001) on the 1st attempt and AS1400 Maxwell RSC-Blood-DNA-Kit protocol on the 2nd attempt. The DNA quality cut off applied were: DNA concentration >20ng/ul and 260/280>1.7 on Nano QC with a combination criterion on applying Qubit QC on high Nano reading/non-optimal 260/280 DNA samples. Methylation status was quantified on the Illumina Infinium MethylationEPIC BeadChip (850K chip) at the Center for Applied Genomics, Children's Hospital of Philadelphia.
DNA extraction was performed on the Biomek FX using the Agencourt Genfind Chemistry and methodologies for 96 samples. Blood samples were typically stored at 4C and were

gently inverted before transferring 0.3ml of its contents into one well of a 96 well plate. The DNA was eluted in 10mM Tris-HCL, 0.1mM EDTA with a PH of 8.0 in a typical volume of 0.18ml. Concentration and quality metrics were performed on a Nanodrop 8000 shortly after extraction. The eluted DNA was stored locally at 4C and stored long term at -20C. Samples were then prepared for bisulfite conversion at a concentration of 12.5ng/ul in a 40ul solution.

DNA was treated with sodium bisulfite using the Qiagen EpiTect Bisulfite Conversion kit (cat# 59104). After the treatment, unmethylated cytosines convert to uracil, while methylated cytosines remain unchanged. For all methylation genotyping samples run at CAG, the Illumina Methylation EPIC Array (cat#20087706) technology was utilized to interrogate SNP loci across an individual's genome in a streamlined three-day process using state-of-the-art Tecan Freedom EVO liquid handlers. Day one consisted of denaturing and neutralizing bisulfite converted DNA samples to prepare them for amplification.
The denatured DNA was isothermally amplified in an overnight step. Day two began by fragmenting the amplified product using end-point fragmentation to avoid over-fragmenting the sample. After an isopropanol precipitation, the fragmented DNA was collected by centrifugation at 4 degrees C. The precipitated DNA was then resuspended in hybridization buffer. The array was then prepared for hybridization in a capillary flow-through chamber. Samples were applied to an array; the loaded array was incubated overnight in the Illumina Hybridization Oven. Day three began with "washing" the hybridized array; unhybridized and non-specifically hybridized DNA was washed away to prepare the chip for staining and extension. Extend and Stain (Xstain) consisted of single-base extension of the oligos on the array, using the captured DNA as a template, incorporates detectable labels on the array and determines the genotype call for the sample. When finished, the beadchips were then loaded into the Illumina iScan, which scans the array using a laser to excite the fluorophore of the single-base extension product on the beads. The scanner recorded high-resolution images of the light emitted from the fluorophores.

The iScan Reader used a laser to excite the fluor of the single-base extension product on the beads of the BeadChip sections. Light emissions from these fluors were then recorded in high-resolution images of the BeadChip sections. Data from these images were analyzed using Illumina's GenomeStudio Methylation Module.

The Illumina MethylationEPIC Array used Cy3 (green) and Cy5 (red) labels to detect cytosine methylation in genomic DNA:

Cy3: Used to label methylated probes

Cy5: Used to label unmethylated probes

Biotin: Incorporated into the primer during allele-specific single base extension

The bisulfite-treated DNA was amplified and loaded onto an Infinium methylation array for the Infinium methylation assay and scanned on the iScan System.

EPICv2 Infinium BeadChip data preprocessing

Preprocessing, quality control, and analysis of the Infinium MethylationEPIC v2 array IDATs files were processed using the SeSAMe package [16]. The standard openSesame workflow was employed to process raw signal data to beta values. Briefly, the openSesame workflow first calculated probe detection P value using the pOOBAH algorithm, which leverages the fluorescence of out-of-band (OOB) probes. It then performed normalization using noob, which uses OOB probes to perform a normal exponential deconvolution of fluorescent intensities, followed by a dye bias correction using the dyeBiasNL function. Signal intensities were then summarized into beta values using the getBetas function. Probes are optionally collapsed to cg-numbers using getBetas function with the collapseToPfx = TRUE option.

The SeSAMe R package was used to load the Illumina EPICv2 array IDAT data files to generate a Beta values matrix with rows including each cg_probe corresponding to a CpG site on the human genome and with columns including each sample ID.

The key Illumina definition files for the Methylation EPIC v2 array can be downloaded at https://support.illumina.com/downloads/infinium-methylationepic-v2-0-product-files.html.

The Sesame R library tool was used for analyzing the different Illumina Methylation Array version data available at https://github.com/zwdzwd/sesame.

The Betas matrix’ "NA" values were used to calculate a per probe "call rate" or "missingness" to create an exclude/quality filter probes with call rate < 90%.

To remove duplicate probes the Sesame function
getBetas(IDATS, collapseToPfx = TRUE)
was used [17].

**Biological age estimation**

Horvath [8], Hannum [9], SkinHorvath [10], BLUP [11], and EN [11] epigenetic clocks were used via the R methyclock package [17], with missing values imputed with KNN [18], and replacing all missing values with 0, 1 and 0.5. A large portion of significant CpG sites for each clock were found to be missing in subject data. This was due to differences in measured CpG sites between earlier generation Methylation BEADchips, used to train and develop the epigenetic clocks, and the methylation 850k beadchip used in this study. The Horvath epigenetic age estimates were trained on 27k and 450k methylation arrays and missingness of CpG site coverage in the 850k methylation array used for the present study may affect predictions. Furthermore, several CpG age predictor sites forming the aging clock did not survive quality control where roughly 250k CpG sites out of 850k configured the final methylation array dataset [16].

We trained a biological age estimator on the AltumAge data set [18] using elastic net modeling to overcome several limitations of using published biological age predictors. The training set was filtered to include only healthy individuals above the age of 18 and sampled via buffy coat or equivalent. PCA analysis was then performed using Scikit-learn on a modified training set that replaces measured values with “1” and missing values with “0” to identify clustering within subject and CpG site measurement. Four series in the training set were then removed due to differences in measured CpGs within subjects.

***Plasma Metabolome***

EDTA plasma, 100 uL per biosample, was randomized, blinded, and analyzed in our in-house metabolomics MS platform as described before [19].

***Urine Lipidome***

Urine samples were obtained at each blood draw and were frozen at -80℃ until analysis. Aliquoted biosamples were randomized, blinded, and submitted for analysis as established in [20]. Urinary eicosanoid metabolites were quantified using ultra-performance liquid chromatography/tandem mass spectrometry (UPLC/MS/MS) as previously described by Meng et al. In brief, PGEM, PGIM, TxM, iPF$_{2\alpha}$-VIII and 8,12-*iso*-iPF$_{2\alpha}$-VI were detected in 0.5 ml urine as follows. The d6 -PGEM (25 ng), d6 -PGDM (25 ng), d3-PGIM (5 ng), d$_3$-2,3-dinor-6k-PGF1a (5 ng), d4-TxM (5 ng), d4-iPF2a-III (5 ng), and d11-8,12-iso-iPF2a-VI (5 ng) stable isotope-labelled internal standards in 50 µl of acetonitrile were added. The sample was equilibrated in methoxyamine (MO) HCl solution (250 µl of 100g MO HCl solid in 100 ml water) for 30 min. Then 200 µl of milliQ water was added to a final volume of 1 ml prior to solid phase extraction (SPE). The samples were passed through a Strata-X 33 µm polymeric reversed phase cartridges (Phenomenex, 8B-S100-TAK) that were preconditioned with 1 ml acetonitrile and 0.25 ml water. The eluate was collected and dried using an Eppendorf Vacufuge. The dried lipid residue was reconstituted in 100 µl of 50% methanol in water prior to analysis. HPLC-MS/MS separation was achieved using a Waters ACQUITY ultra-performance liquid chromatography (UPLC) system with a 2.1 x 150 mm UPLC column with 1.7 µm particles (Waters ACQUITY UPLC CSH C18) with mobile phase A consisting of water with 0.5% ammonium acetate at pH 5.7 and mobile phase B consisting of acetonitrile-methanol mixture (95:5). A flow rate of 350 µl/min, and a linear solvent gradient from 5% to 45% mobile phase B over 30 min was used to separate metabolites.

The second set of eicosanoid metabolites analyzed include 14(15)-DHET, 11(12)-DHET, 8(9)-DHET, 5(6)-DHET, LTE$_4$, 5(S)-HETE, 12-HETE, 15-HETE, 9(10)-DiHOME, 9-HODE, and 13-HODE. To another 0.5 ml aliquot of urine, 5 ng of deuterated standards of d11-14(15)-DHET, d11-11(12)-DHET, d11-8(9)-DHET, d5-LTE4, d8-5-HETE, d8-12-

HETE, d8-15-HETE, and d4-13-HODE in 50 µl of acetonitrile. The sample was brought to 1 ml with the addition of 450 µl acetonitrile and passed through SPE and eluted, dried, and reconstituted as previously described. Lipid separation was also conducted using a Waters ACQUITY UPLC and a 2.1 x 150 mm UPLC column with 1.7 µm particles (Waters ACQUITY UPLC BEH C18). Mobile phase A was prepared from water containing 0.1% formic acid and mobile phase B consisted of acetonitrile:methanol (95:5; v/v) containing 0.1% formic acid. The flow rate was set to 350 µl/min in which separations were conducted using various linear solvent gradients. Quantification was achieved using a single point calibration curve with the standard mixes and all the analyte's concentration levels in the samples were below the concentration where the calibration curve starts to bend. The multiple reaction monitoring (MRM) LC–MS method has a linear range of 4 magnitudes. Values below the lowest level of a typical calibration curve, but above the limit of detection (LOD) were also reported.

All urinary metabolite concentrations were normalized by quantifying creatinine using LC-MS. The stable isotope-labelled internal standard d3-creatinine, 10 µg/ml in 3% water/acetonitrile, was added to 10 µl of each urine sample. This mixture was then diluted with 200 µl acetonitrile. A 2.1 × 50 mm UPLC column with 2.5 µm particles (Waters XBridge BEH HILIC) was used to perform the separations. Mobile phase A was composed of acetonitrile (100%), and mobile phase B consisted of a 5 mM ammonium formate in water solution (pH = 3.98) with a flow rate of 350 µl/min.

***Saliva Microbiome***

Saliva, 200uL per biosample, was transferred into a bead tube containing 600 uL lysis buffer and stored at -80◦C until analysis at the PennCHOP Microbiome Center.

DNA Purification

DNA was extracted from approximately 200 mg of stool or tissue using the Qiagen DNeasy PowerSoil Pro kit. Extracted DNA was quantified with the Quant-iT PicoGreen Assay Kit.

Library Preparation and Sequencing

DNA was quantified using the Quant-iT PicoGreen dsDNA assay kit (Thermo Fisher Scientific) before library generation. Shotgun libraries were generated from 0.52 ng DNA using Illumina Nextera XT Library Prep kit and Nextera unique dual indexes at 1:4 scale reaction volume. Library success was assessed by Quant-iT PicoGreen dsDNA assay and samples with library yields < 1 ng/ul were re-prepped as needed. After all samples for a given pool were prepped, an equal volume of library was pooled from every sample and then the pool was sequenced using a 300 cycle Nano kit on the Illumina MiSeq. Libraries were then repooled based on the demultiplexing statistics of the MiSeq Nano run. Final libraries were QCed on the Agilent BioAnalyzer to check the size distribution and absence of additional adaptor fragments. Libraries were sequenced on an Illumina NovaSeq 6000 v1.5 flow cell, producing 2x150 bp paired-end reads. Extraction blanks and nucleic acid-free water were processed along with experimental samples to empirically assess environmental and reagent contamination. A laboratory-generated mock community consisting of DNA from Vibrio campbellii and Lambda phage were included as a positive sequencing control.

Bioinformatics processing

Shotgun metagenomic data were analyzed using Sunbeam, a user-extendable bioinformatics pipeline that we developed for this purpose [21]. Quality control steps were performed by the default workflows in Sunbeam, which are optimized to remove host-derived sequences and reads of low sequence complexity. The abundance of bacteria were estimated using Kraken [22]. Reads were mapped to the KEGG database [23] using Diamond [24] to estimate the abundance of bacterial gene orthologs, as well as to curated databases of genes involved in butyrate production, polysaccharide utilization, and secondary bile acid production. Sample similarity were assessed by Bray-Curtis and Jaccard distances, and community-level differences between sample groups were assessed using the PERMANOVA test.

***Bioinformatics Pipeline***

**Unsupervised principal component analysis**

To address the question whether age-specific oscillatory differences are preserved when the dimensionality of the omics data sets is reduced to enable integrative transomic modeling. Here, principal component analysis (PCA) of the oscillatory amplitudes derived from each separate omic data set revealed that the first two principal components, PC1 and PC2, explained large parts of the total variability. Thus, PC1 and PC2 explained 17.4% and 8.4%, respectively, of the observed variance in the transcriptome, 26.9% and 6.0% in the AB-proteome, 15.9% and 9.1% in the methylome, and 47.3% and 15.2%, respectively, in the phenome. Sex did not fully explain the observed variance in PC1 (Figure 3b). Unsupervised projections onto the amplitudes of the oscillating transcriptome clearly separated young from old participants in PC2 (p=0.035 for the Mann-Whitney test to ascertain whether one group has, generally, greater PC2 values than the other one, Figure 3b top left). Projections onto the amplitudes of the oscillating AB-proteome and methylome showed little age-specific divergence in PC2 (p=0.4359 and p=0.7959, respectively, in the Mann-Whitney test). Projections onto the amplitudes of the oscillating features in the phenome overlapped largely between young and old in PC2 (p=0.1655 in the Mann-Whitney test, Figure 3b bottom right). PCA enriched for oscillating features from each omic data set showed overall larger variance explained in PC1, for example, 24.5% in the transcriptome and 44.1% in the AB-proteome compared to the respective 17.4% and 26.9% variance explained in the full data set described above (Figure 3c). For PC2, the projections on the amplitudes of the oscillating features showed similar age-specific divergence (transcriptome, AB-proteome and methylome) and convergence (phenome) (Figure 3c). Taken together, this shows that age-specific characteristics were retained among high dimensional oscillating principal components derived, most evidently, from the transcriptome.

**Unsupervised canonical correlation analysis (CCA)**

To determine whether this structure is conserved when we relate the amplitudes of features from the transcriptome with amplitudes of features from the AB-proteome, we deployed canonical correlation analysis (CCA). The new canonical variables, CC1 and

CC2, show variance like PC1 and PC2 in the PCA. Age associated with CC1. However, neither CC1 nor CC2 was associated with sex (Figure 3d). Taken together, we found that age-specific characteristics in the data structure were not only retained in the transomic integration but explained the largest part of the observed variance.

**Omics Data Preprocessing**

The datasets we collected in this study are heterogeneous and require a range of normalizations and transformations before they are suitable for analysis. Here's the list of preprocessing operations we performed on each dataset:

- *RNA-Seq*: Removed genes with low expression. Specifically, we only kept genes with PORT-normalized read counts > 0 for all samples in at least one combination of age group and time of day.
- *DNA methylation*: None beyond the QC filtering we performed while processing the raw array data.
- *Olink Proteomics*: Assays for six proteins (CXCL8, IDO1, IL6, LMOD1, SCRIB, TNF) are repeated across multiple Olink panels. To avoid analyzing these proteins multiple times within the same subject, we identified the assay for each protein with the highest variability. Briefly, within each repeated assay and subject we calculated the mean-subtracted abundance values for these six proteins. Then, we calculated the standard deviation of these mean-subtracted values across all subjects within each assay. Finally, for each of the six proteins we selected the assay with the highest standard deviation as the one to retain for all downstream analyses and discarded the remaining repeats.
- *MS proteomics*: Removed features missing data from all subjects in more than two timepoints, missing all data from more than five subjects within one age cohort, or missing all data from one group of an experimental factor of interest (age cohort, time of day, sex).
- *MS untargeted metabolomics*: None.
- *Urine eicosanoids*: Replaced all missing abundance values (NA's in the spreadsheet) with the lower limit of detection for each lipid.

- *Saliva microbiome*: Removed low abundance microbial genera with mean relative proportions less than 0.001. We also log2 transformed the relative proportion values.

**Two-Factor Analysis for Age and Time of Day Effects**

To identify features with significant differences in abundance by age (old vs young) or time of day (morning; 0800 vs evening; 2000) we performed an ANOVA-style two-factor analysis. Briefly, we limited these analyses to those samples collected at the 0800 timepoint (n=3 per subject) and the 2000 timepoint (n=2 per subject). Using these samples, across both age cohorts, we fit a mixed-effects linear model to the abundance of each feature which included fixed-effect terms for age group, time of day, and their interaction. This model also included a random effects term to account for repeated measurements from each subject. Within each omic dataset, we applied a Benjamini-Hochberg multiple testing correction to the p-values testing for significant age, time of day, or interaction effects across all features. For the RNA-Seq and DNA methylation (M-values) datasets, we performed these fits in R v4.4.1 using v3.60.3 of the limma package. For the Olink dataset, we used the *olink_lmer* function from v3.8.2 of the OlinkAnalyze package to perform these fits against the NPX abundance values. For all other omics datasets, we performed fits with the *lme* function from v3.1-164 of the nlme package. We used the Benjamini-Hochberg (BH) method [25] to correct the resulting *p*-values for multiple testing, as implemented by the *p.adjust* function from the base R stats package. Note, the limma and OlinkAnalyze packages apply a BH correction internally, so we did not need to apply any additional corrections.

**Detecting Oscillating Features at the Cohort Levels Using Cosinor Fits**

We identified features with circadian oscillations in abundance using a cosinor analysis [26-28]. Briefly, we fit a cosine curve to the abundance measurements for each feature using a linear model. This model included fixed effects terms for sex, age group, two cosinor parameters, and interactions between age group and the cosinor parameters. The model also included a random effects term to account for repeated measurements from each subject. From this model, we tested both cosinor terms simultaneously to identify features with significant oscillations. We tested the interaction terms to identify any features that show significant differences in oscillation between the two age cohorts. We also tested the age group term to identify features with an age-dependent change in MESOR. Within each omic datatset, we applied a Benjamini-Hochberg multiple testing correction to these p-values testing for significant oscillations and age-related changes. Additionally, we used this model to calculate cosinor parameters (MESOR, amplitude, and phase) for features with significant oscillations within each age group. Note, for these analyses we fixed the period of our cosinor fits to 24 hours, to focus on those features with circadian oscillations in abundance. For the RNA-Seq and DNA methylation datasets, we performed these fits and hypothesis testing using v3.60.3 of the limma package. For all other omics datasets, we performed cosinor fits with the *lmer* function from v1.1-35.5 of the lme4 package, and calculated *p*-values for significant cycling/interaction/MESOR terns with the *KRmodcomp* function from v0.5.3 of the pbkrtest package. As with the two-factor analyses, we used the *p.adjust* function to apply a BH correction to the cosinor *p*-values. The limma-based analyses performed this correction internally and did not require us to use the *p.adjust* function.

**Bayesian Information Criterion to Compare Explanatory Models of Rhythmicity**

As an alternative method to characterize cycling and non-cycling features across age groups we implemented an approach based around the Bayesian Information Criterion (BIC). This is the approach taken by the dryR package [29]. In each omics dataset, we started by fitting four models to each feature: 1) feature abundance cycles in both young and old subjects; 2) feature abundance cycles in old, but not young subjects; 3) feature abundance cycles in young, but not old subjects; 4) feature abundance cycles in neither young nor old subjects. We performed these fits using linear mixed-effects models (similar to the one we used for the cosinor fits) that accounted for repeated measures within each subject. Next, we compared the results across these four fits to determine which model was most likely to explain the data. We use the Bayesian Information Criterion (BIC) value from each fit to assess its quality. Lastly, we applied a weighting scheme to the BIC values from all four models using a softmax function, so that they sum to 1 within each feature.

The model with the highest weighted BIC value was considered the one with the best fit and most likely to explain the data. As with the cosinor fits, we performed these fits using v3.60.3 of the limma package for the RNA-seq and DNA methylation datasets, or the *lmer* function from v1.1-35.5 of the lme4 package for all other omics datasets.

**Multi-Block Data Analysis**

We submitted feature amplitudes as proxy for oscillatory patterns to the generalized canonical correlation analysis (gCCA) using published frameworks [30]. In short, CCA generalizes PCA to two datasets, and generalized CCA to multiple data sets. We use generalized CCA because this algorithm affords to include additional datasets.

**Integration of modalities with mixOmics**

We first performed integration focusing on circadian oscillatory patterns present in the data. We removed features that lack significant circadian oscillations in abundance in both young and old cohorts (Figure 4a, panel A). Removal criterion was set to be population level oscillation q-value ≥ 0.1 in both young and old groups. The q-values were calculated by applying a Benjamini-Hochberg multiple testing correction to p-values obtained from the mixed-effect cosinor model described in (Detecting Oscillating Features at the Cohort Levels Using Cosinor Fits).

For each retained feature and each subject, we used cosinor fit to calculate the amplitude of oscillation of the abundance of the feature in the subject (Figure 4a, panel B). These amplitudes were then assembled into matrices (Figure 4a, panel C), one matrix per modality, which were provided to mixOmics (Figure 4a, panel D & E) [31]. The information about the age group of each subject (young/old) was also provided to mixOmics.

Multiblock sPLS-DA (DIABLO) approach is used by mixOmics to find latent variables (components) which are combinations of input variables (amplitudes of oscillation of abundances of measured features) that are highly correlated within and across modalities (Figure 4b, panel A-E) and which separate between young and old cohorts (i.e., there is a difference in population level amplitude between young and old cohorts for features that comprise the components). These two optimization goals are oftentimes competing (*Supplementary Figure 4*, panel A vs. B) and their importance was weighted using the following design matrix in which both rows and columns represent modalities (Figure 4a, panel E):

For every modality mixOmics outputs one first latent variable (t1, u1, v1 in *Supplementary Figure 4*), one second latent variable (t2, u2, v2 in *Supplementary Figure 4*), etc. The set of all first latent variables across all modalities (t1, u1, v1 in *Supplementary Figure 4*) is represented visually as a Circos plot in Figure 4b, panel F.

#### Supplementary Tables

##### *Supplementary Table 1*

Demographics of Study Participants

| Age Group | Subject ID# | Age Range* | Sex | Race | Ethnicity | BMI (kg/m^2) |
|---|---|---|---|---|---|---|
| Young | 1 | 26-30 | Female | White | Not Hispanic or Latino | 23.9 |
| Young | 2 | 26-30 | Male | White | Not Hispanic or Latino | 29.4 |
| Young | 3 | 26-30 | Male | White | Not Hispanic or Latino | 20.8 |
| Young | 4 | 20-25 | Male | White | Not Hispanic or Latino | 19.8 |
| Young | 5 | 26-30 | Female | Chose not to answer | Chose not to answer | 19.1 |
| Young | 6 | 26-30 | Female | White | Not Hispanic or Latino | 23.5 |
| Young | 7 | 20-25 | Male | White | Hispanic or Latino | 22.4 |
| Young | 8 | 20-25 | Female | White | Not Hispanic or Latino | 25.2 |
| Young | 9 | 20-25 | Male | White | Not Hispanic or Latino | 21.7 |
| Young | 10 | 26-30 | Male | White | Hispanic or Latino | 19.8 |
| Old | 11 | 71-75 | Male | White | Not Hispanic or Latino | 28.0 |
| Old | 12 | 71-75 | Female | White | Not Hispanic or Latino | 27.9 |
| Old | 13 | 71-75 | Male | White | Not Hispanic or Latino | 27.8 |
| Old | 14 | 66-70 | Male | White | Not Hispanic or Latino | 27.8 |
| Old | 15 | 66-70 | Female | White | Not Hispanic or Latino | 24.7 |
| Old | 16 | 66-70 | Female | Black or African American | Not Hispanic or Latino | 27.1 |
| Old | 17 | 51-55 | Male | White | Not Hispanic or Latino | 28.0 |
| Old | 18 | 66-70 | Female | Black or African American | Not Hispanic or Latino | 25.1 |
| Old | 19 | 61-65 | Female | White | Hispanic or Latino | 23.8 |
| Old | 20 | 61-65 | Male | White | Not Hispanic or Latino | 26.6 |

* 20-25, 26-30, 31-35, 36-40, 41-45, 46-50, 51-55, 56-60, 61-65, 66-70, 71-75 yr

***Supplementary Table 2***

Breakdown of data points collected.

| Dataset | Observation Period | Measurement Resolution | Aggregated Resolution | Number of Data Points | Number of Features |
|---|---|---|---|---|---|
| **Actigraphy** | 1361 hours (+/- 342) | 60 hz | every minute | 10,842,906 | 6 |
| **EKG** | 96.1 hours (+/- 53) | 1000 hz | every minute (Kubios results) | 4,516,429 | 49 |
| **ABPM** | 29.91 hours (+/- 17.46) | every 15 mins (30 mins at night) | no aggregation | 1,499 | 5 |
| **Grip strength** | 48 hours | every 12 hours | no aggregation | 990 | 10 |
| **Auditory function** | 48 hours | every 12 hours | no aggregation | 396 | 4 |
| **Cognitive function tests** | 48 hours | every 12 hours | no aggregation | 16,264 | 983 |
| **Air quality** | 95.06 hours (+/- 62.12) | every hour | no aggregation | 5,596 | 4 |
| **RNA-seq (Whole blood)** | 48 hours | Day 1 - every 12 hours; Day 2 - every 4 hours | no aggregation | 6,765,126 | 37,794 |
| **AB proteomics (Plasma)** | 48 hours | Day 1 - every 12 hours; Day 2 - every 4 hours | no aggregation | 523,575 | 2,925 |
| **MS proteomics (Plasma)** | 48 hours | Day 1 - every 12 hours; Day 2 - every 4 hours | no aggregation | 71,562 | 503 |
| **DNA methylation (Buffy coat)** | 48 hours | Day 1 - every 12 hours; Day 2 - every 4 hours | no aggregation | 120,683,066 | 866,553 |
| **Metabolomics (Plasma)** | 48 hours | Day 1 - every 12 hours; Day 2 - every 4 hours | no aggregation | 25,760 | 153 |
| **Lipidomics (Urine)** | 48 hours | Day 1 - every 12 hours; Day 2 - every 4 hours | no aggregation | 3,204 | 18 |
| **Microbiome (Saliva)** | 48 hours | Day 1 - every 12 hours; Day 2 - every 4 hours | no aggregation | 359,100 | 2,052 |
| ***Total*** | *NA* | *NA* | *NA* | ***143,815,473*** | ***911,059*** |

#### ***Supplementary Table 3***

Cycling Molecular Features in the Aging Landscape (*cosinor-q*<0.05; *delta-mesor-q*<0.05)

| DATA TYPE | TISSUE | QCED FEATURES | YOUNG | OLD | EITHER | BOTH | OLD ONLY | YOUNG ONLY | MESOR DIFFERENCES |
|---|---|---|---|---|---|---|---|---|---|
| RNA-SEQ | Whole blood | 18728 | 3945 (21.06%) | 5444 (29.07%) | 7148 (38.17%) | 2241 (11.97%) | 3203 (17.1%) | 1704 (9.1%) | 2844 (15.19%) |
| EPIC DNA-METHYLATION | Buffy coat | 644928 | 0 (0%) | 758 (0.12%) | 758 (0.12%) | 0 (0%) | 758 (0.12%) | 0 (0%) | 140623 (21.8%) |
| AB PROTEOMICS | Plasma | 2925 | 1262 (43.15%) | 1144 (39.11%) | 1438 (49.16%) | 968 (33.09%) | 176 (6.02%) | 294 (10.05%) | 79 (2.7%) |
| MS PROTEOMICS | Plasma | 394 | 102 (25.89%) | 101 (25.63%) | 137 (34.77%) | 66 (16.75%) | 35 (8.88%) | 36 (9.14%) | 20 (5.1%) |
| METABOLOMICS-UNTARGETED | Plasma | 153 | 45 (29.41%) | 57 (37.25%) | 72 (47.06%) | 30 (19.61%) | 27 (17.65%) | 15 (9.8%) | 0 (0%) |
| LIPIDOMICS | Urine | 18 | 9 (50%) | 0 (0%) | 9 (50%) | 0 (0%) | 0 (0%) | 9 (50%) | 0 (0%) |
| MICROBIOME | Saliva | 39 | 14 (35.9%) | 11 (28.21%) | 18 (46.15%) | 7 (17.95%) | 4 (10.26%) | 7 (17.95%) | 0 (0%) |
| KEGG PATHWAYS: RNA-SEQ | Whole blood | 340 | 162 (47.65%) | 155 (45.59%) | 209 (61.47%) | 108 (31.76%) | 47 (13.82%) | 54 (15.88%) | NA |
| KEGG PATHWAYS: AB PROTEOMICS | Plasma | 330 | 300 (90.91%) | 298 (90.3%) | 308 (93.33%) | 290 (87.88%) | 8 (2.42%) | 10 (3.03%) | NA |
| KEGG PATHWAYS: MS PROTEOMICS | Plasma | 193 | 116 (60.1%) | 132 (68.39%) | 137 (70.98%) | 111 (57.51%) | 21 (10.88%) | 5 (2.59%) | NA |
| KEGG PATHWAYS: METAGENOMICS | Saliva | 186 | 28 (15.05%) | 16 (8.6%) | 35 (18.82%) | 9 (4.84%) | 7 (3.76%) | 19 (10.22%) | NA |

How to read this table: In the first row "RNA-seq", cyclic features for gene transcripts measured in whole blood using 0.05 as the q-value cutoff were evident in 21% (3945/18728) of the genes in young and 29% (5444/18728) in old. About 38% of the transcriptome oscillated in either young or old (7148/18728), while 12% unique genes cycled in both young and old (2241/18728). Only in old, 17% (3203/18728) were oscillatory genes, and 9% (1704/18728) only in young.

#### *Supplementary Table 4*

KEGG pathway analysis

**Rhythmic in Young, Nonrhythmic in Old (*Cosinor-q* Young < 0.05, *Cosinor-q* Old > 0.05)**

| **Pathway & ID** | Cosinor-*p* Young | Cosinor-*q* Young | **Cosinor-*p* Old** | **Cosinor-*q* Old** |
|---|---|---|---|---|
| **Hedgehog signaling pathway \| path:hsa04340** | 0.0002 | 0.0004 | 0.953 | 0.965 |
| **Various types of N-glycan biosynthesis \| path:hsa00513** | 0.0005 | 0.001 | 0.247 | 0.270 |
| **N-Glycan biosynthesis \| path:hsa00510** | 0.001 | 0.001 | 0.208 | 0.229 |
| **Glycosaminoglycan biosynthesis - keratan sulfate \| path:hsa00533** | 0.001 | 0.001 | 0.146 | 0.165 |
| **PPAR signaling pathway \| path:hsa03320** | 0.001 | 0.002 | 0.168 | 0.188 |
| **Viral protein interaction with cytokine and cytokine receptor \| path:hsa04061** | 0.002 | 0.002 | 0.045 | 0.052 |
| **Ascorbate and aldarate metabolism \| path:hsa00053** | 0.003 | 0.004 | 0.306 | 0.331 |
| **Sphingolipid metabolism \| path:hsa00600** | 0.006 | 0.007 | 0.055 | 0.064 |
| **Complement and coagulation cascades \| path:hsa04610** | 0.006 | 0.007 | 0.289 | 0.314 |
| **Glycosphingolipid biosynthesis - ganglio series \| path:hsa00604** | 0.006 | 0.008 | 0.050 | 0.058 |
| **Mismatch repair \| path:hsa03430** | 0.008 | 0.010 | 0.412 | 0.441 |
| **Homologous recombination \| path:hsa03440** | 0.014 | 0.016 | 0.385 | 0.412 |
| **Nucleocytoplasmic transport \| path:hsa03013** | 0.023 | 0.027 | 0.442 | 0.469 |
| **RNA polymerase \| path:hsa03020** | 0.032 | 0.038 | 0.283 | 0.308 |

**Nonrhythmic in Young, Rhythmic in Old (*Cosinor-q* Young > 0.05, *Cosinor-q* Old < 0.05)**

| **Pathway & ID** | Cosinor-*p* Young | Cosinor-*q* Young | **Cosinor-*p* Old** | **Cosinor-*q* Old** |
|---|---|---|---|---|
| **Pyruvate metabolism \| path:hsa00620** | 0.068 | 0.077 | 0.004 | 0.006 |
| **Proteasome \| path:hsa03050** | 0.267 | 0.288 | 0.008 | 0.011 |

| | | | | |
|---|---|---|---|---|
| **Collecting duct acid secretion \| path:hsa04966** | 0.092 | 0.103 | 0.009 | 0.011 |
| **Bile secretion \| path:hsa04976** | 0.417 | 0.441 | 0.012 | 0.015 |
| **Proximal tubule bicarbonate reclamation \| path:hsa04964** | 0.240 | 0.261 | 0.016 | 0.020 |
| **Vitamin digestion and absorption \| path:hsa04977** | 0.607 | 0.626 | 0.026 | 0.031 |
| **Morphine addiction \| path:hsa05032** | 0.051 | 0.058 | 0.033 | 0.039 |
| **Non-homologous end-joining \| path:hsa03450** | 0.404 | 0.429 | 0.034 | 0.041 |
| **Neuroactive ligand-receptor interaction \| path:hsa04080** | 0.708 | 0.728 | 0.037 | 0.043 |

***Supplementary Table 5***

Oscillatory triads of transcriptomic, proteomic and methylated features converging on a specific gene in young and old

| **Ome** | **Feature** | **Gene** | **Location** | **Young ~ q** | **Old ~ q** | **Δ ~ q** | **Young A** | **Old A** | **Young Φ** | **Old Φ** | **Young MESOR** | **Old MESOR** | **Δ MESOR q** |
|---|---|---|---|---|---|---|---|---|---|---|---|---|---|
| Transcriptome | ENSG00000026508 | CD44 | | 0.1584 | **0.0004** | 0.45 | 0.04 | 0.10 | 1.06 | 2.91 | 12.26 | 12.29 | 0.79 |
| Methylome | cg05313151 | CD44 | 11:35104878 | 0.3733 | **0.0516** | 0.76 | 0.07 | 0.15 | 5.12 | 11.52 | -2.18 | -2.19 | 0.95 |
| MS-Proteome | sp\|P16070\|CD44_HUMAN | CD44 | | 0.5926 | **0.0166** | 0.88 | 0.02 | 0.08 | 18.10 | 12.99 | 0.05 | -0.09 | 0.38 |
| Transcriptome | ENSG00000099622 | CIRBP | | **0.0049** | 0.1631 | 0.71 | 0.12 | 0.07 | 3.01 | 1.90 | 12.57 | 12.48 | 0.25 |
| AB-Proteome | OID31503 | CIRBP | | **2.41E-06** | 4.38E-05 | 0.95 | 1.23 | 1.06 | 16.63 | 17.16 | 0.90 | 0.54 | 0.65 |
| Methylome | cg02917867 | CIRBP-AS1 | 19:1265999 | **0.1434** | 0.0768 | 0.98 | 0.18 | 0.15 | 21.99 | 18.75 | -0.51 | -0.62 | 0.64 |
| Transcriptome | ENSG00000153815 | CMIP | | 0.0760 | **0.0060** | 0.35 | 0.10 | 0.10 | 14.26 | 10.27 | 9.99 | 10.05 | 0.60 |
| Methylome | cg23004006 | CMIP | 16:81664541 | 0.5838 | **0.0485** | 0.93 | 0.04 | 0.10 | 23.91 | 22.61 | 1.00 | 0.94 | 0.50 |
| AB-Proteome | OID31406 | CMIP | | 7.80E-07 | **3.71E-05** | 0.92 | 1.48 | 1.20 | 16.37 | 17.18 | 2.59 | 2.37 | 0.84 |
| Transcriptome | ENSG00000035664 | DAPK2 | | **0.0039** | 0.0067 | 0.93 | 0.19 | 0.16 | 13.28 | 12.49 | 9.57 | 9.54 | 0.87 |
| Methylome | cg09052865 | DAPK2 | 15:64219504 | **0.1468** | 0.2543 | 1.00 | 0.15 | 0.09 | 18.96 | 20.59 | 1.74 | 1.64 | 0.46 |
| AB-Proteome | OID30505 | DAPK2 | | **8.44E-07** | 0.0302 | 0.92 | 0.74 | 0.36 | 16.65 | 17.36 | -1.43 | -1.54 | 0.85 |
| Transcriptome | ENSG00000147443 | DOK2 | | **0.0064** | 0.0562 | 0.24 | 0.13 | 0.07 | 13.02 | 17.50 | 10.27 | 10.43 | 0.20 |
| AB-Proteome | OID20138 | DOK2 | | **1.60E-07** | 2.83E-05 | 0.92 | 1.83 | 1.40 | 16.35 | 17.10 | 3.33 | 3.06 | 0.84 |
| Methylome | cg16410556 | DOK2,GFRA2 | 8:21598308 | **0.1468** | 0.1262 | 1.00 | 0.10 | 0.07 | 18.09 | 19.31 | 1.33 | 1.23 | 0.07 |
| Transcriptome | ENSG00000150907 | FOXO1 | | 0.0002 | **0.0025** | 0.81 | 0.15 | 0.12 | 23.63 | 0.61 | 9.12 | 8.74 | 0.0010 |
| Methylome | cg19828062 | FOXO1 | 13:41172346 | 0.4819 | **0.0292** | 0.88 | 0.10 | 0.30 | 10.13 | 10.93 | 1.81 | 1.93 | 0.52 |
| AB-Proteome | OID20515 | FOXO1 | | 4.39E-07 | **3.41E-05** | 0.92 | 1.11 | 0.87 | 16.22 | 17.17 | 1.94 | 1.35 | 0.32 |
| Transcriptome | ENSG00000130787 | HIP1R | | 0.0025 | **0.0006** | 0.31 | 0.21 | 0.20 | 2.59 | 23.92 | 9.31 | 8.89 | 0.09 |
| Methylome | cg27534833 | HIP1R | 12:123347897 | 0.2131 | **0.0584** | 1.00 | 0.07 | 0.11 | 18.68 | 20.42 | 1.23 | 1.05 | 0.0056 |
| AB-Proteome | OID31141 | HIP1R | | 0.0338 | **0.0097** | 0.92 | 0.14 | 0.13 | 15.93 | 19.41 | 0.31 | 0.70 | 0.51 |

| | | | | | | | | | | | | | |
|---|---|---|---|---|---|---|---|---|---|---|---|---|---|
| Transcriptome | ENSG00000168918 | INPP5D | | 0.0010 | **0.0087** | 0.76 | 0.11 | 0.08 | 12.82 | 11.48 | 11.97 | 12.02 | 0.65 |
| Methylome | cg12315466 | INPP5D | 2:233923814 | 0.1468 | **0.0995** | 1.00 | 0.10 | 0.08 | 20.01 | 20.77 | 0.97 | 0.87 | 0.31 |
| AB-Proteome | OID30259 | INPP5D | | 3.11E-05 | **0.0001** | 0.92 | 0.77 | 0.62 | 16.55 | 18.73 | 1.92 | 1.92 | 1.00 |
| Transcriptome | ENSG00000091409 | ITGA6 | | 0.0008 | **0.0000** | 0.81 | 0.25 | 0.28 | 2.29 | 1.45 | 9.55 | 8.94 | 0.0045 |
| Methylome | cg22061832 | ITGA6 | 2:173293627 | 0.3287 | **0.0580** | 1.00 | 0.12 | 0.22 | 19.10 | 20.46 | -1.04 | -0.91 | 0.58 |
| AB-Proteome | OID20528 | ITGA6 | | 4.09E-05 | **0.0460** | 0.92 | 0.41 | 0.26 | 17.52 | 16.27 | -2.29 | -2.61 | 0.31 |
| Transcriptome | ENSG00000163956 | LRPAP1 | | **0.0006** | 0.4604 | 0.34 | 0.11 | 0.04 | 11.24 | 13.28 | 9.73 | 9.83 | 0.42 |
| Methylome | cg02846431 | LRPAP1 | 4:3526685 | **0.1148** | 0.5040 | 0.89 | 0.24 | 0.08 | 18.55 | 20.64 | 2.16 | 2.03 | 0.54 |
| Methylome | cg07955126 | LRPAP1 | 4:3526676 | **0.1321** | 0.0757 | 1.00 | 0.23 | 0.19 | 19.17 | 21.41 | 1.80 | 1.70 | 0.72 |
| AB-Proteome | OID21059 | LRPAP1 | | **7.32E-06** | 0.0093 | 0.92 | 0.32 | 0.20 | 16.31 | 16.90 | 0.26 | 0.50 | 0.83 |
| Transcriptome | ENSG00000162889 | MAPKAPK2 | | **1.04E-06** | 9.52E-06 | 0.95 | 0.13 | 0.11 | 12.10 | 11.80 | 10.68 | 10.66 | 0.76 |
| Methylome | cg06262043 | MAPKAPK2 | 1:206904153 | **0.1452** | 0.0787 | 1.00 | 0.10 | 0.09 | 20.66 | 21.13 | 0.44 | 0.26 | 0.28 |
| AB-Proteome | OID31293 | MAPKAPK2 | | **2.00E-07** | 3.92E-05 | 0.92 | 1.02 | 0.76 | 16.32 | 17.23 | 1.99 | 1.67 | 0.63 |
| Transcriptome | ENSG00000008130 | NADK | | **0.0014** | 0.0172 | 0.83 | 0.16 | 0.11 | 13.95 | 13.10 | 12.04 | 12.20 | 0.25 |
| Methylome | cg24163210 | NADK | 1:1713944 | **0.1482** | 0.2410 | 1.00 | 0.14 | 0.09 | 20.39 | 19.59 | -0.85 | -1.25 | 0.0035 |
| AB-Proteome | OID20178 | NADK | | **0.0117** | 0.3309 | 0.92 | 0.20 | 0.12 | 20.93 | 22.82 | -0.58 | -0.22 | 0.37 |
| Transcriptome | ENSG00000131196 | NFATC1 | | 0.3670 | **0.0050** | 0.72 | 0.05 | 0.10 | 0.46 | 0.10 | 8.38 | 8.26 | 0.23 |
| Methylome | cg27106643 | NFATC1 | 18:77257352 | 0.4223 | **0.0403** | 0.93 | 0.10 | 0.27 | 8.81 | 10.91 | 1.62 | 1.76 | 0.59 |
| Methylome | cg15260951 | NFATC1 | 18:77165150 | 0.1669 | **0.0292** | 1.00 | 0.15 | 0.24 | 7.50 | 9.52 | 1.38 | 1.40 | 0.92 |
| AB-Proteome | OID20545 | NFATC1 | | 1.72E-07 | **1.69E-05** | 0.92 | 1.31 | 1.04 | 16.27 | 17.17 | 2.16 | 2.16 | 0.99 |
| Transcriptome | ENSG00000123405 | NFE2 | | **0.0064** | **0.0251** | 0.92 | 0.20 | 0.16 | 14.74 | 14.06 | 11.15 | 11.56 | 0.0072 |
| Methylome | cg17230002 | NFE2 | 12:54689504 | **0.1321** | **0.0815** | 1.00 | 0.23 | 0.19 | 19.47 | 18.93 | 0.49 | 0.09 | 0.0151 |
| AB-Proteome | OID30268 | NFE2 | | **2.11E-07** | **3.41E-05** | 0.92 | 1.24 | 0.92 | 16.23 | 17.41 | 2.64 | 2.50 | 0.88 |
| Transcriptome | ENSG00000116962 | NID1 | | **0.0064** | 0.0054 | 0.51 | 0.21 | 0.22 | 4.12 | 1.75 | 7.14 | 7.36 | 0.60 |
| Methylome | cg12303769 | NID1 | 1:236107252 | **0.1468** | 0.0692 | 1.00 | 0.13 | 0.12 | 18.26 | 20.59 | 1.63 | 1.36 | 0.17 |
| AB-Proteome | OID20362 | NID1 | | **1.66E-06** | 0.3352 | 0.92 | 0.29 | 0.09 | 16.39 | 15.55 | 0.52 | 0.64 | 0.72 |
| Transcriptome | ENSG00000099250 | NRP1 | | 0.1315 | **0.0036** | 0.52 | 0.16 | 0.32 | 8.49 | 4.53 | 4.44 | 3.80 | 0.06 |
| MS-Proteome | sp\|O14786\|NRP1_HUMAN | NRP1 | | 0.2226 | **0.0236** | 0.88 | 0.07 | 0.13 | 16.33 | 14.34 | 0.16 | -0.03 | 0.18 |

| | | | | | | | | | | | | | |
|---|---|---|---|---|---|---|---|---|---|---|---|---|---|
| Methylome | cg20127733 | NRP1 | 10:33635645 | 0.4571 | **0.0320** | 0.90 | 0.08 | 0.23 | 8.75 | 10.21 | 1.15 | 1.05 | 0.63 |
| Methylome | cg12054892 | NRP1 | 10:33607723 | 0.1627 | **0.0547** | 1.00 | 0.20 | 0.24 | 22.10 | 22.00 | -1.73 | -2.22 | 0.0182 |
| Transcriptome | ENSG00000196923 | PDLIM7 | | **0.0159** | **0.0248** | 0.97 | 0.16 | 0.15 | 11.84 | 12.47 | 10.25 | 10.43 | 0.23 |
| Methylome | cg06449191 | PDLIM7 | 5:176912100 | **0.1439** | **0.0333** | 1.00 | 0.25 | 0.28 | 19.06 | 20.73 | 1.77 | 1.48 | 0.08 |
| Methylome | cg01235607 | PDLIM7 | 5:176912137 | **0.1468** | **0.0750** | 1.00 | 0.13 | 0.12 | 19.05 | 21.33 | 1.08 | 0.96 | 0.14 |
| AB-Proteome | OID20729 | PDLIM7 | | **1.33E-07** | **2.39E-05** | 0.92 | 2.05 | 1.56 | 16.20 | 16.98 | 4.10 | 3.95 | 0.93 |
| Transcriptome | ENSG00000142657 | PGD | | **0.0014** | 0.0052 | 0.95 | 0.16 | 0.13 | 11.45 | 11.04 | 11.36 | 11.46 | 0.60 |
| Methylome | cg02916418 | PGD | 1:10480153 | **0.1468** | 0.2296 | 1.00 | 0.18 | 0.10 | 18.99 | 20.52 | 0.80 | 0.59 | 0.0442 |
| AB-Proteome | OID30344 | PGD | | **0.0010** | 0.0005 | 0.96 | 0.46 | 0.46 | 17.20 | 17.91 | -0.16 | 0.29 | 0.43 |
| Transcriptome | ENSG00000166949 | SMAD3 | | 0.0363 | **0.0007** | 0.86 | 0.13 | 0.19 | 1.24 | 0.89 | 9.77 | 9.67 | 0.46 |
| Methylome | cg15490565 | SMAD3 | 15:67390907 | 0.3699 | **0.0491** | 0.51 | 0.08 | 0.18 | 23.96 | 13.04 | -3.13 | -3.22 | 0.45 |
| Methylome | cg04620612 | SMAD3 | 15:67336616 | 0.5732 | **0.0110** | 0.64 | 0.05 | 0.14 | 15.85 | 21.20 | 1.60 | 1.49 | 0.07 |
| AB-Proteome | OID31418 | SMAD3 | | 6.38E-06 | **1.96E-05** | 0.94 | 0.78 | 0.74 | 16.61 | 17.35 | 0.83 | 0.73 | 0.90 |
| Transcriptome | ENSG00000177156 | TALDO1 | | **0.0115** | 0.0152 | 0.99 | 0.17 | 0.15 | 12.66 | 12.34 | 12.19 | 12.35 | 0.18 |
| Methylome | cg25043021 | TALDO1 | 11:763133 | **0.1452** | 0.2780 | 0.97 | 0.17 | 0.10 | 19.26 | 22.39 | 2.00 | 1.87 | 0.38 |
| AB-Proteome | OID30414 | TALDO1 | | **2.46E-05** | 0.0002 | 0.92 | 0.55 | 0.46 | 16.52 | 17.48 | 0.91 | 0.92 | 0.98 |
| Transcriptome | ENSG00000109906 | ZBTB16 | | 0.0034 | **1.23E-07** | 0.54 | 0.25 | 0.41 | 9.52 | 8.76 | 7.48 | 7.01 | 0.0298 |
| Methylome | cg03725573 | ZBTB16 | 11:113962901 | 0.8725 | **0.0301** | 0.77 | 0.01 | 0.09 | 22.39 | 21.04 | 1.28 | 1.41 | 0.0274 |
| AB-Proteome | OID21205 | ZBTB16 | | 9.99E-08 | **2.23E-05** | 0.92 | 1.05 | 0.79 | 16.36 | 17.17 | 1.57 | 1.58 | 0.99 |

Each oscillatory triad consists of three features, the gene transcript, CpG sites for this gene locus and the protein, latter quantified either on the mass spec (MS) or antibody (AB) analytical platforms. For some gene loci, more than one CpG site is reported. Features testing statistically significant for oscillatory behavior are marked as green cells, and oscillatory triads are marled in bold. Cut-off significant levels are $q<0.017$ for the transcriptome, $q<0.2$ for the methylome, and $q<0.05$ for the proteomics.

***Supplementary Table 6***

Oscillating proteins in the chronobiome associated with disease phenotypes and mortality in the UKBB

| **UKBB Phenotype** | **Total associated proteins UKBB** | **Chronobiome Explanatory model (BIC > 0.75)** | | | |
|---|---|---|---|---|---|
| | | **Cycling in Young & Old** | **Cycling in Only Young** | **Cycling in Only Old** | **Cycling in Neither** |
| Type 2 diabetes | 1008 | 242 | 66 | 23 | 453 |
| Death | 810 | 116 | 69 | 19 | 397 |
| Liver disease | 769 | 99 | 58 | 16 | 406 |
| Ischemic heart disease | 716 | 121 | 61 | 19 | 332 |
| COPD | 676 | 95 | 62 | 17 | 322 |
| Ischemic stroke | 392 | 49 | 40 | 10 | 187 |
| Rheumatoid arthritis | 289 | 27 | 20 | 11 | 153 |
| Lung cancer | 214 | 21 | 25 | 7 | 112 |
| Systemic lupus erythematosus | 178 | 15 | 18 | 3 | 99 |
| Inflammatory bowel disease | 72 | 7 | 11 | 3 | 32 |
| Parkinson's disease | 26 | 4 | 3 | 1 | 13 |
| ***Sum*** | ***5150*** | ***796*** | ***433*** | ***129*** | ***2506*** |

The disease-specific prediction for each AB-protein abundance in the UKBB [32] is merged with the time-specific AB-protein abundance in the chronobiome. UKBB protein disease associations with significance threshold of Bonferroni-adjusted p-values $p<3.1e-6$. The Bayesian Information Criterion (BIC) in the chronobiome denotes the confidence to categorize the diurnal variability of a protein of interest (BIC>0.75 as threshold). Only disease phenotypes are shown with protein associations of $n \geq 20$.

#### Supplementary Figures

##### *Supplementary Figure 1*

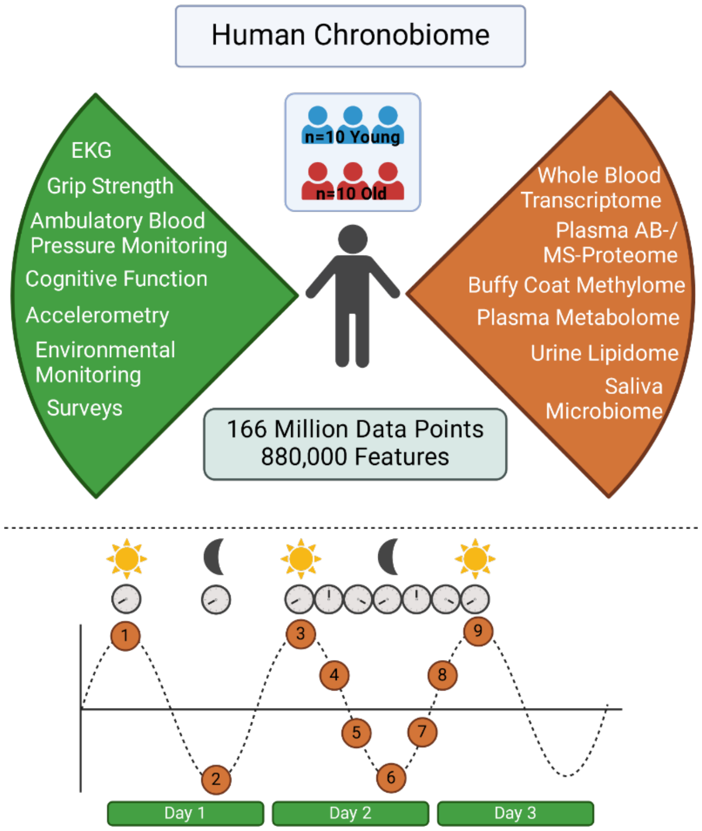

Clinical study design. Cohorts of apparent healthy young and old (n=10 per group) were consented to enroll in this prospective, observational clinical study where a suite of wearable devices, smartphone apps and clinical assessments (green) were aligned with biosampling (orange) (3) over three days with sparse 12-hour sampling on day 1 followed by dense sampling in 4-hour intervals on days 2 and 3 (timepoints 1-9 bottom).

***Supplementary Figure 2***

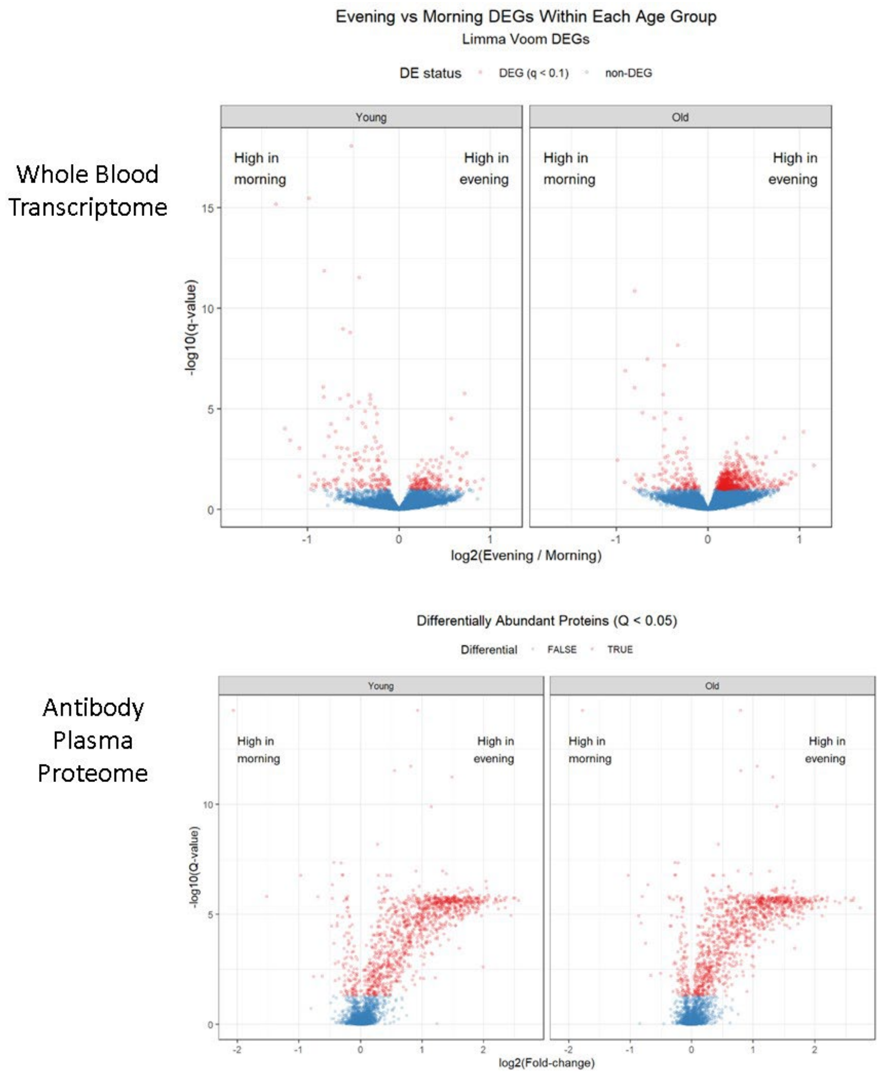

(A) Volcano plots for the whole blood transcriptome (top) and the plasma antibody proteome (bottom) displaying the distribution of differentially expressed genes (DEGs, q<0.01) and differentially expressed proteins (DEPs, q<0.05), respectively, for young (left) and old (right) highlighted in red compared to non-significant features in blue. Within each age cohort's distribution, the tail ends indicate time-of-day specific expression: in the left flank features cluster with higher expression in the morning, while features in the right flank show higher expression in the evening.

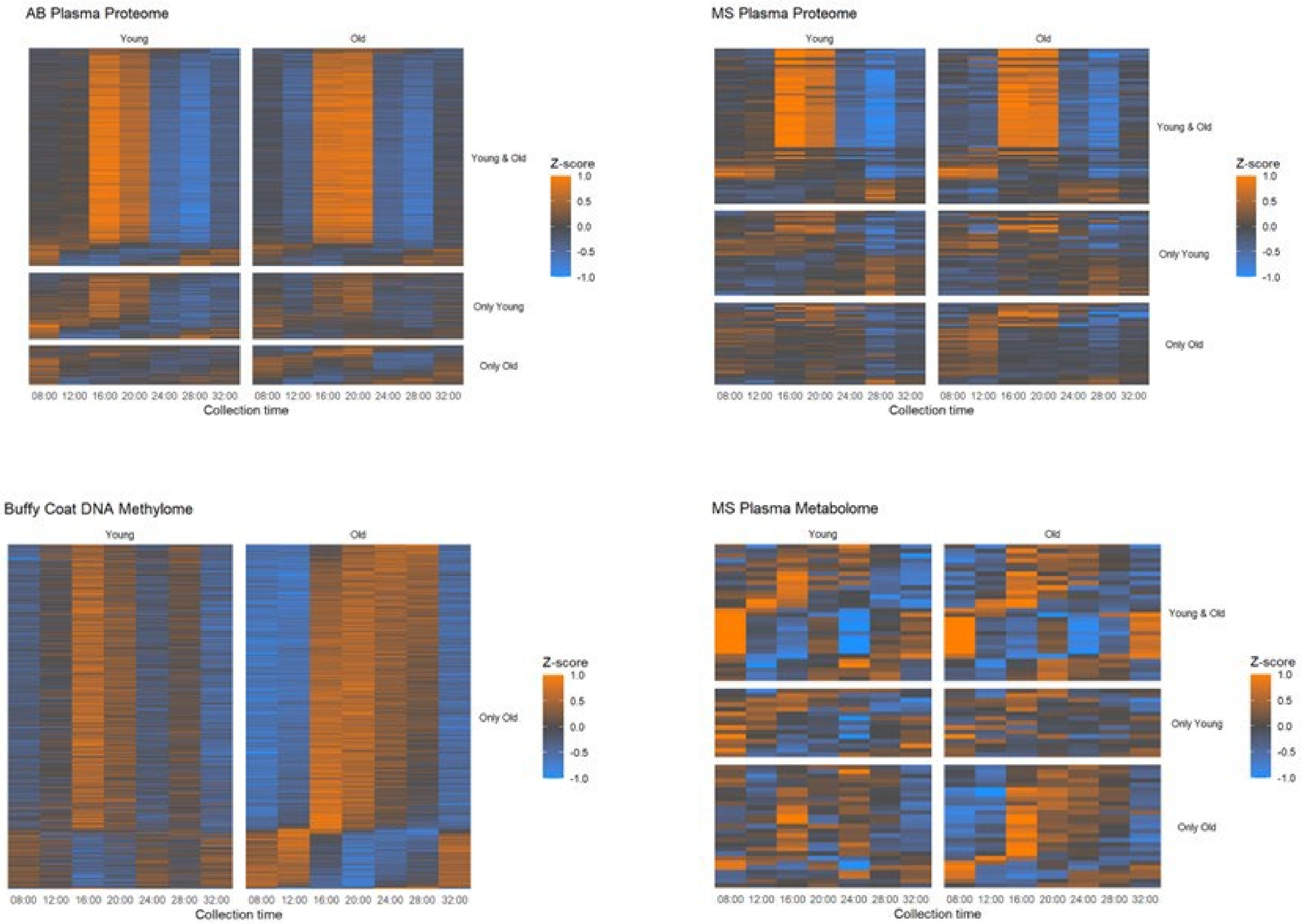

(B) Heatmaps for distinct 'omes in young (left column) and old (right column) visualize the variability in feature abundance quantified in 4hr intervals over 24 hours. Each row in a given heatmap displays abundance data of a single feature. The data have been z-normalized to anchor a feature's mean to zero (0) and the standard deviation to one (1). This normalization achieves values that are comparable across different scales. In each heatmap, features are sorted from top to bottom according to their acrophase, the time of day when peak abundances were quantified. Similarly timed features thus appear side-by-side and enhance appreciation of oscillatory behavior. The top rectangles show oscillatory features in young and old while subsets of features oscillate either in young only (center) or old only (bottom).

*Supplementary Figure 3*

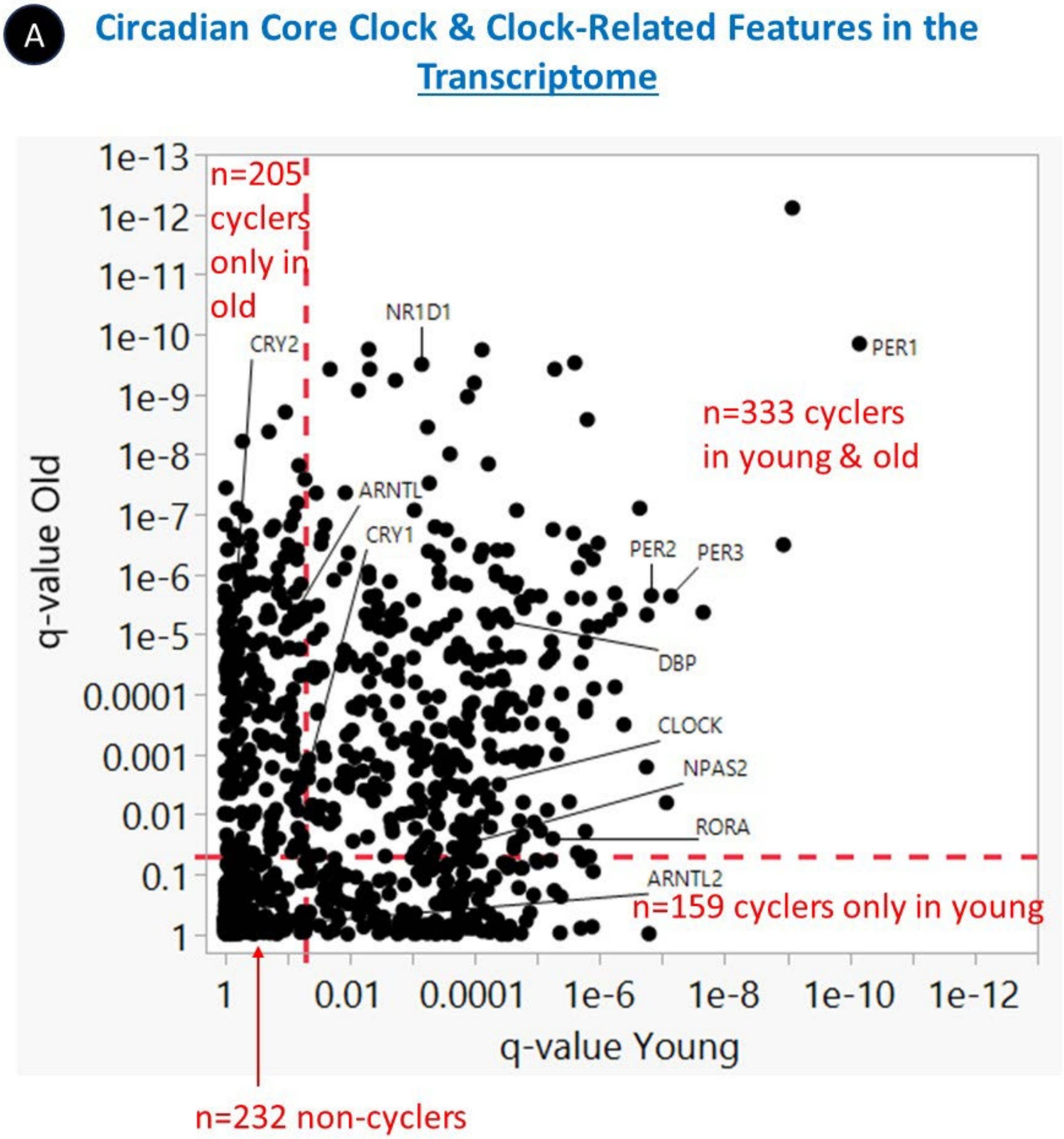

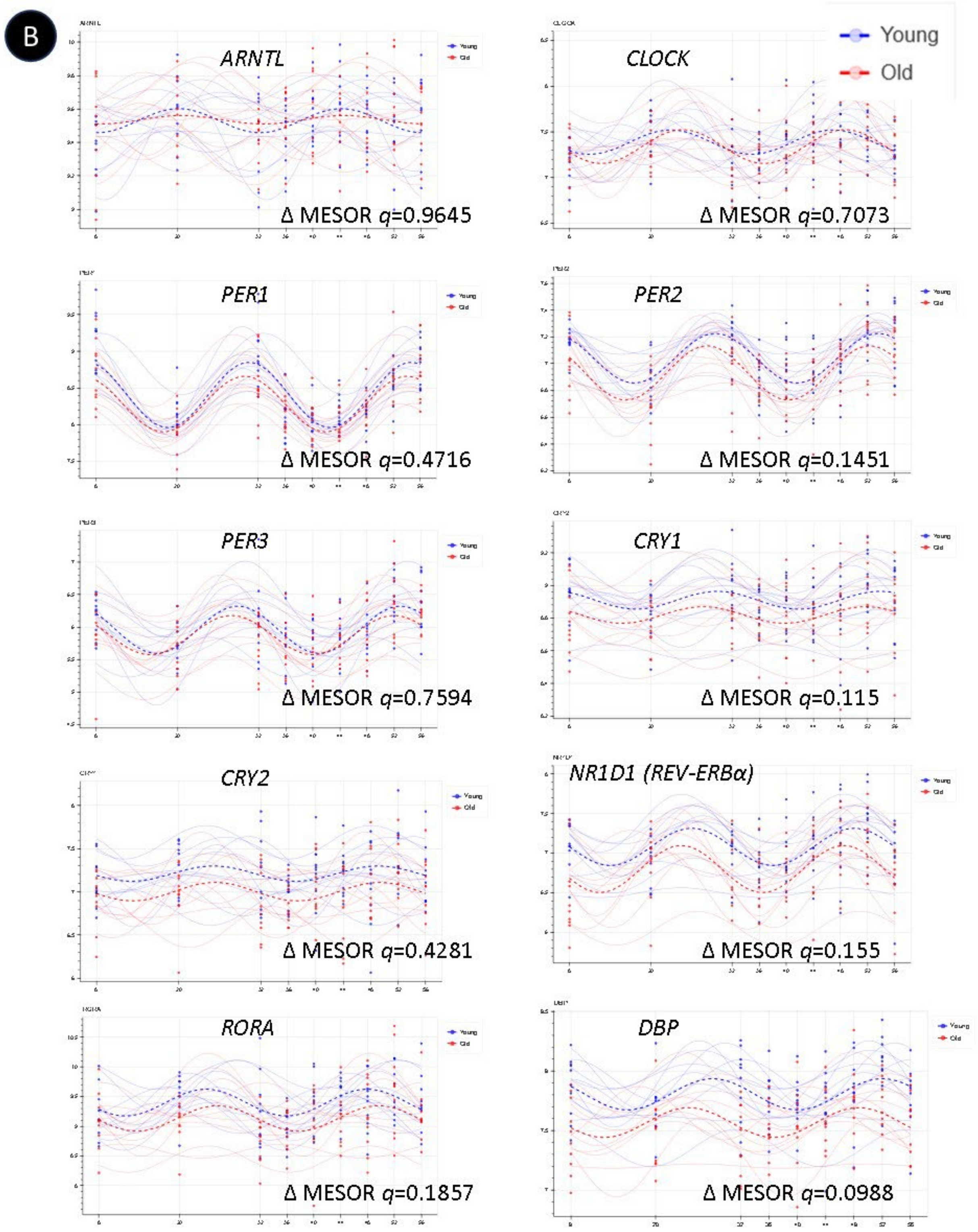
B
ARNTL
Young
Old
Δ MESOR q=0.9645
CLOCK
Δ MESOR q=0.7073
PER1
Δ MESOR q=0.4716
PER2
Δ MESOR q=0.1451
PER3
Δ MESOR q=0.7594
CRY1
Δ MESOR q=0.115
CRY2
Δ MESOR q=0.4281
NR1D1 (REV-ERBα)
Δ MESOR q=0.155
RORA
Δ MESOR q=0.1857
DBP
Δ MESOR q=0.0988

C

#### Circadian Core Clock & Clock-Related Features in the AB-Proteome

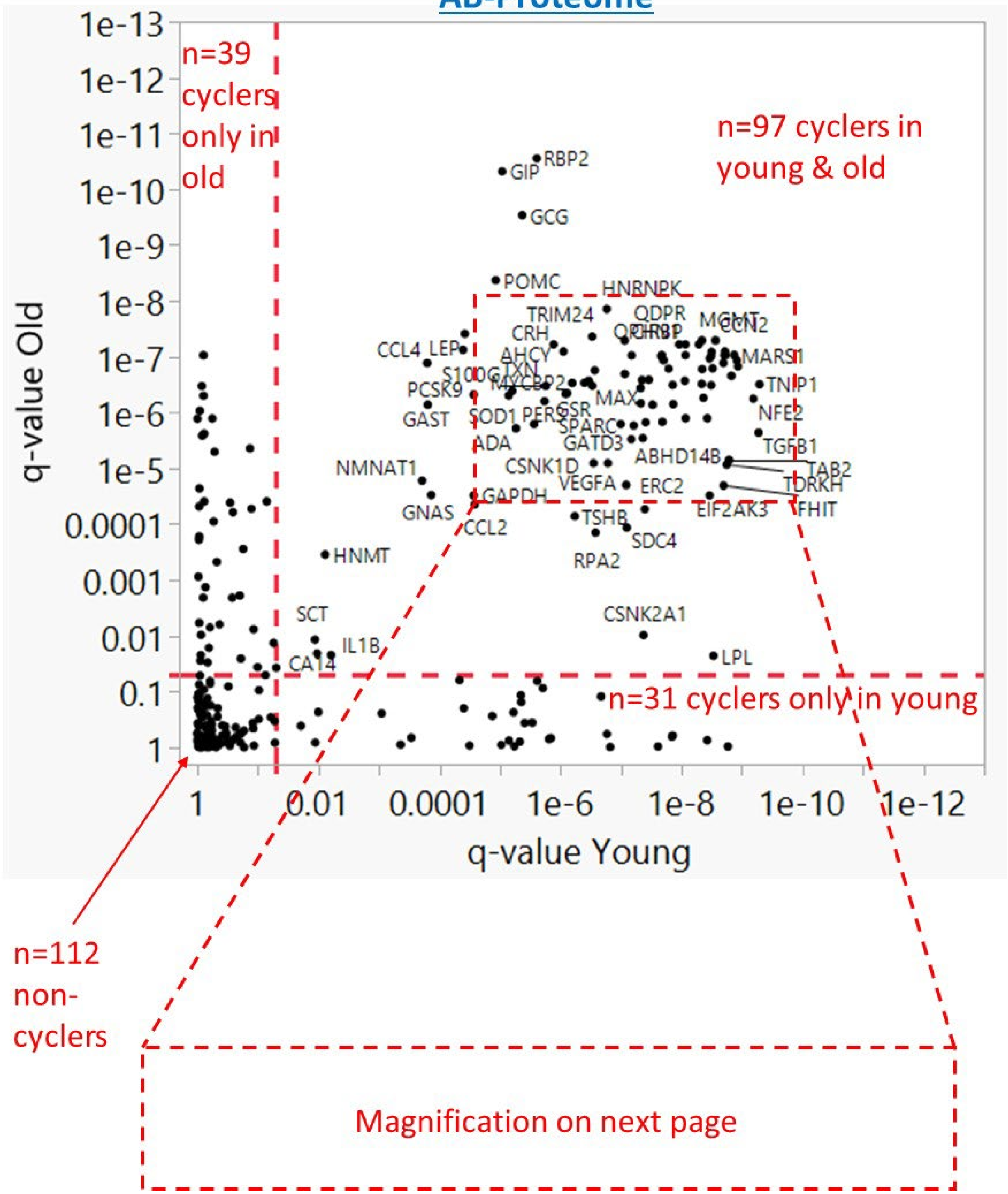

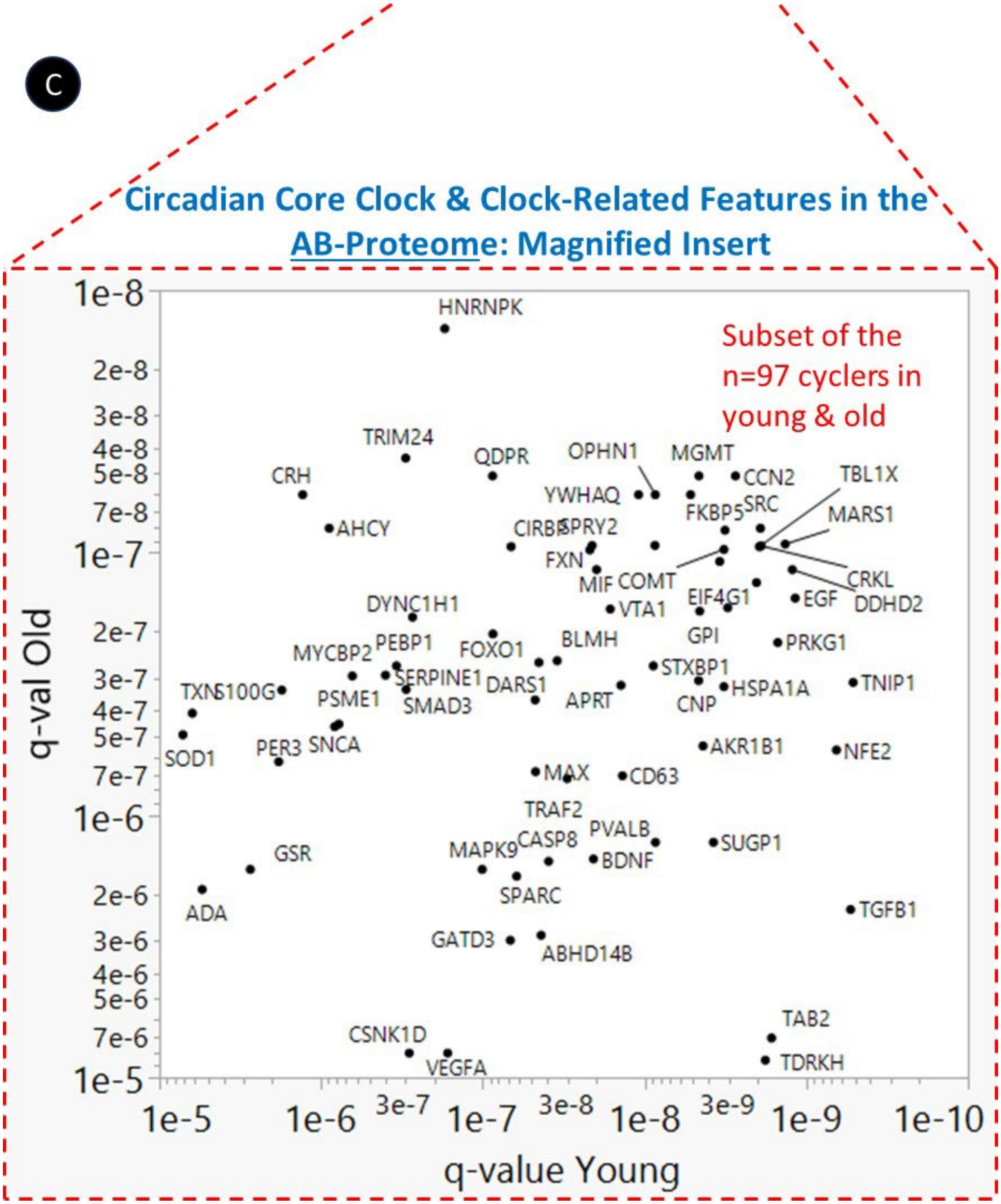
C
Circadian Core Clock & Clock-Related Features in the AB-Proteome: Magnified Insert
Subset of the n=97 cyclers in young & old
q-val Old
q-value Young
HNRNPK
TRIM24
QDPR
OPHN1
MGMT
CCN2
TBL1X
CRH
YWHAQ
FKBP5
SRC
MARS1
AHCY
CIRBP
SPRY2
FXN
MIF
COMT
CRKL
DDHD2
EIF4G1
EGF
DYNC1H1
VTA1
GPI
BLMH
PRKG1
MYCBP2
PEBP1
FOXO1
STXBP1
TXN
S100G
SERPINE1
DARS1
HSPA1A
TNIP1
PSME1
SMAD3
APRT
CNP
SOD1
PER3
SNCA
AKR1B1
NFE2
MAX
CD63
TRAF2
PVALB
CASP8
MAPK9
BDNF
SUGP1
GSR
SPARC
ADA
TGFB1
GATD3
ABHD14B
TAB2
CSNK1D
VEGFA
TDRKH

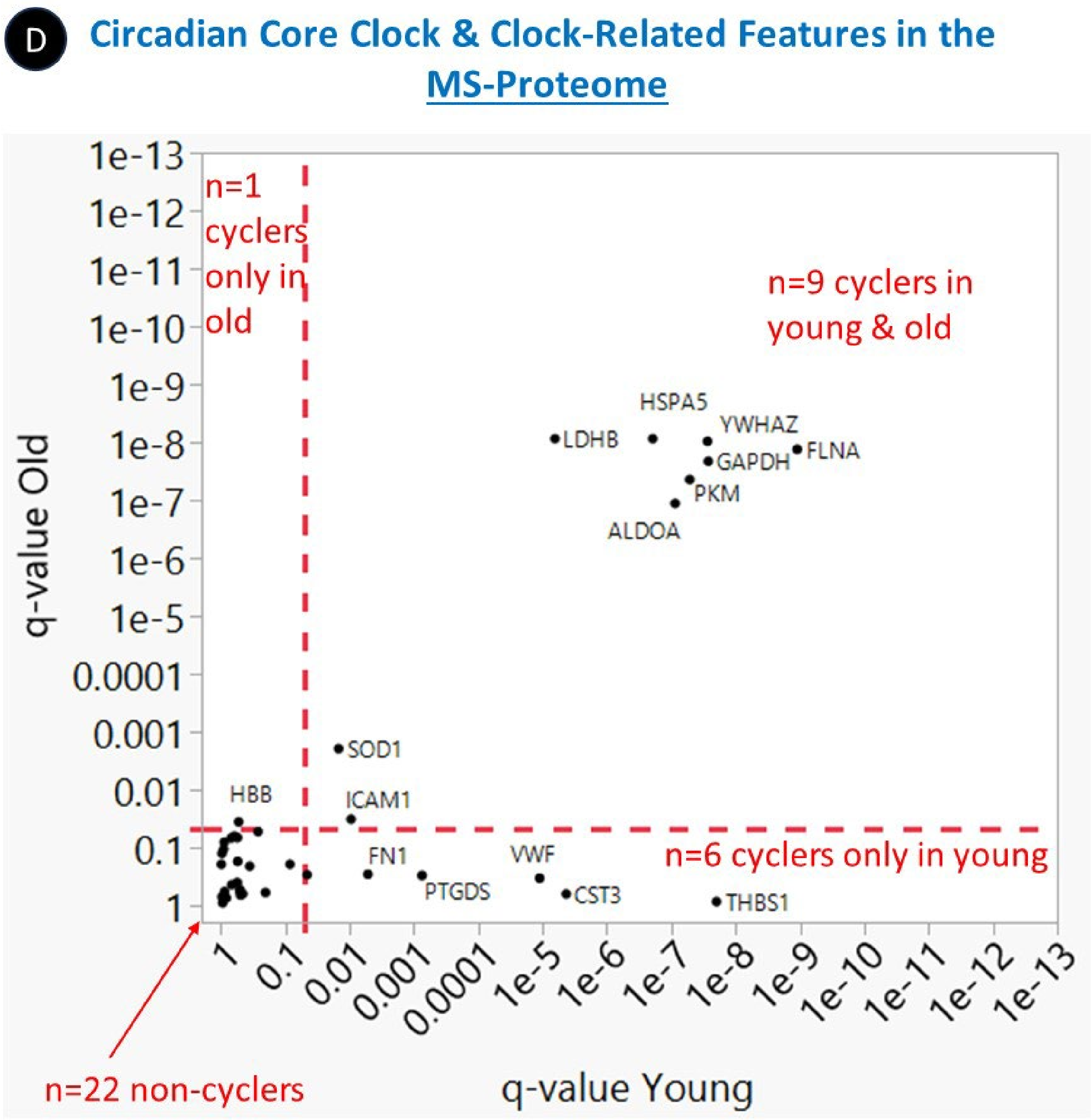

The dashed red lines denote cosinor q=0.05 and divide the dot plot into four quadrants identifying circadian core clock & clock-related features in the (A) transcriptome with (B) diurnal gene expression patterns of molecular clock gene transcripts over 48 hours with age-specific abundance levels indicated by significant Δ mesor q-values, (C) AB-proteome, and (D) MS-proteome, which cycle in both young and old (top right), only in old (top left), only in young (bottom right) and in none (bottom left).

#### *Supplementary Figure 4*

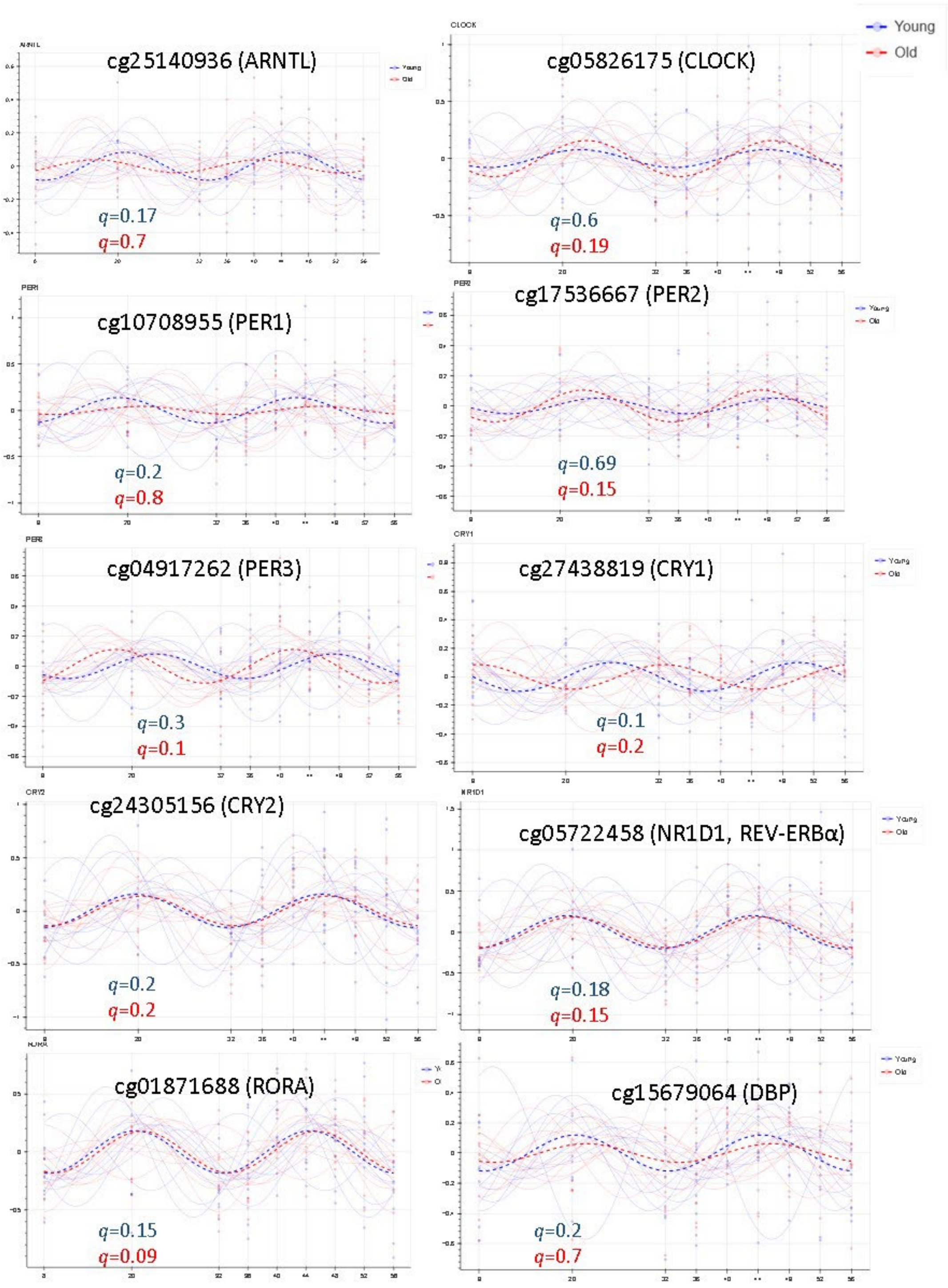

Diurnal variability in methylated gene loci associated by proximity with core clock genes using a cutoff of cosinor-*q*≤0.2.

#### *Supplementary Figure 5*

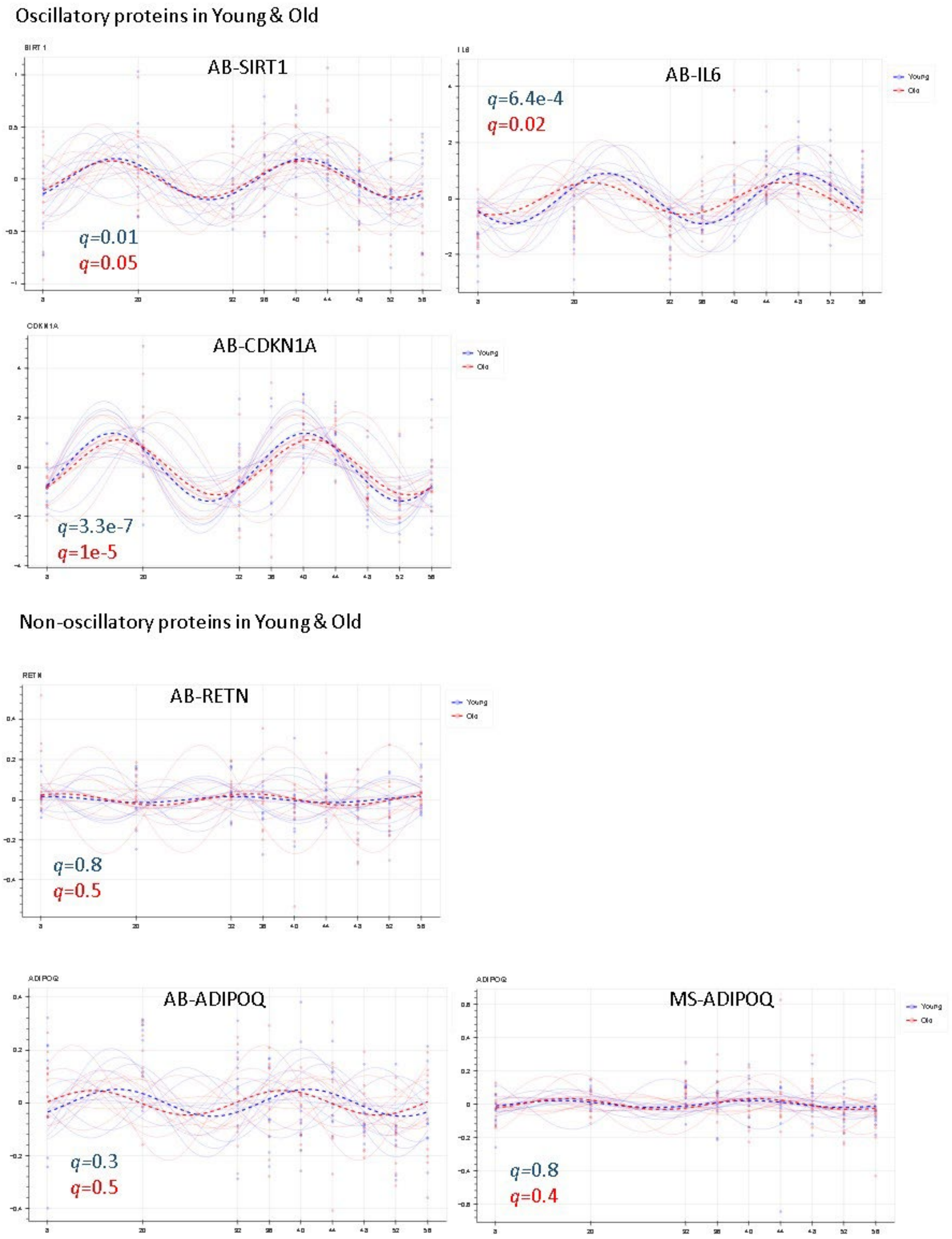

Non-oscillatory proteins in Young & Old - continued

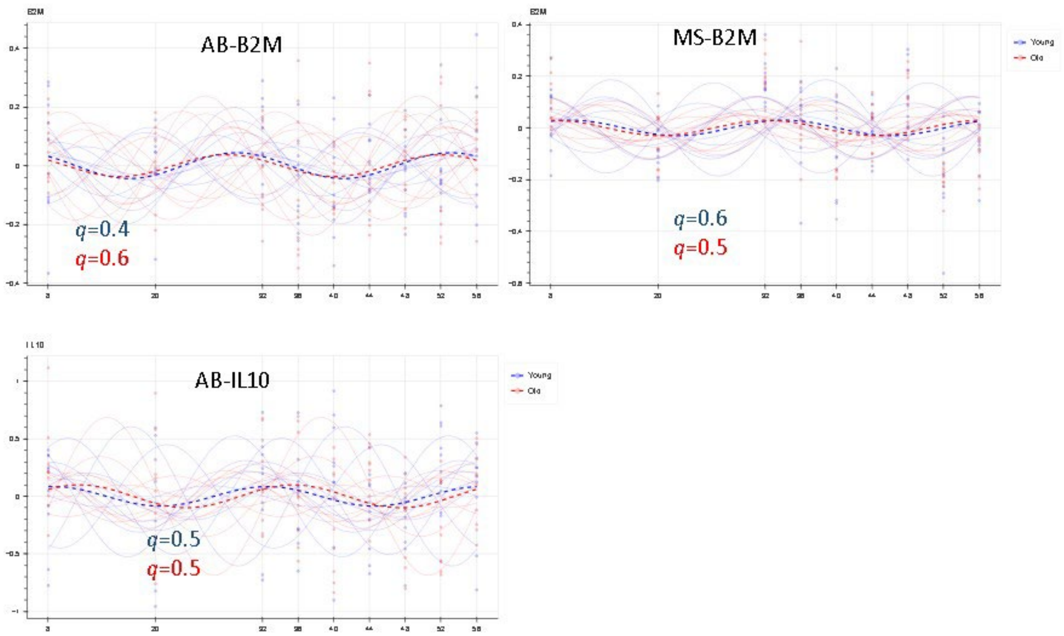

Diurnal variability in proteomic biomarkers associated with inflammaging and circadian clocks.

#### *Supplementary Figure 6*

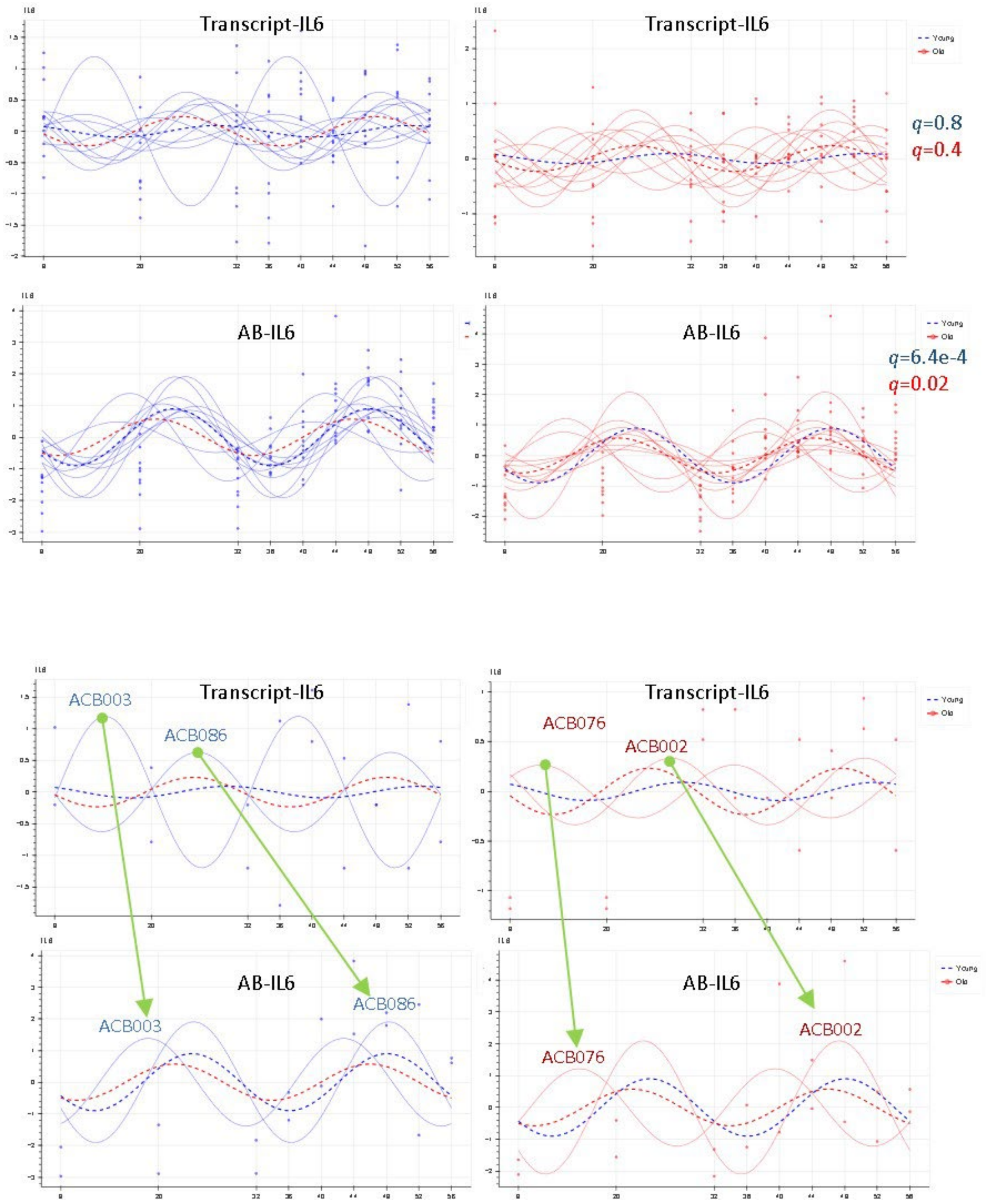

Comparative diurnal variability in IL-6 transcript and AB-protein.

***Supplementary Figure 7***

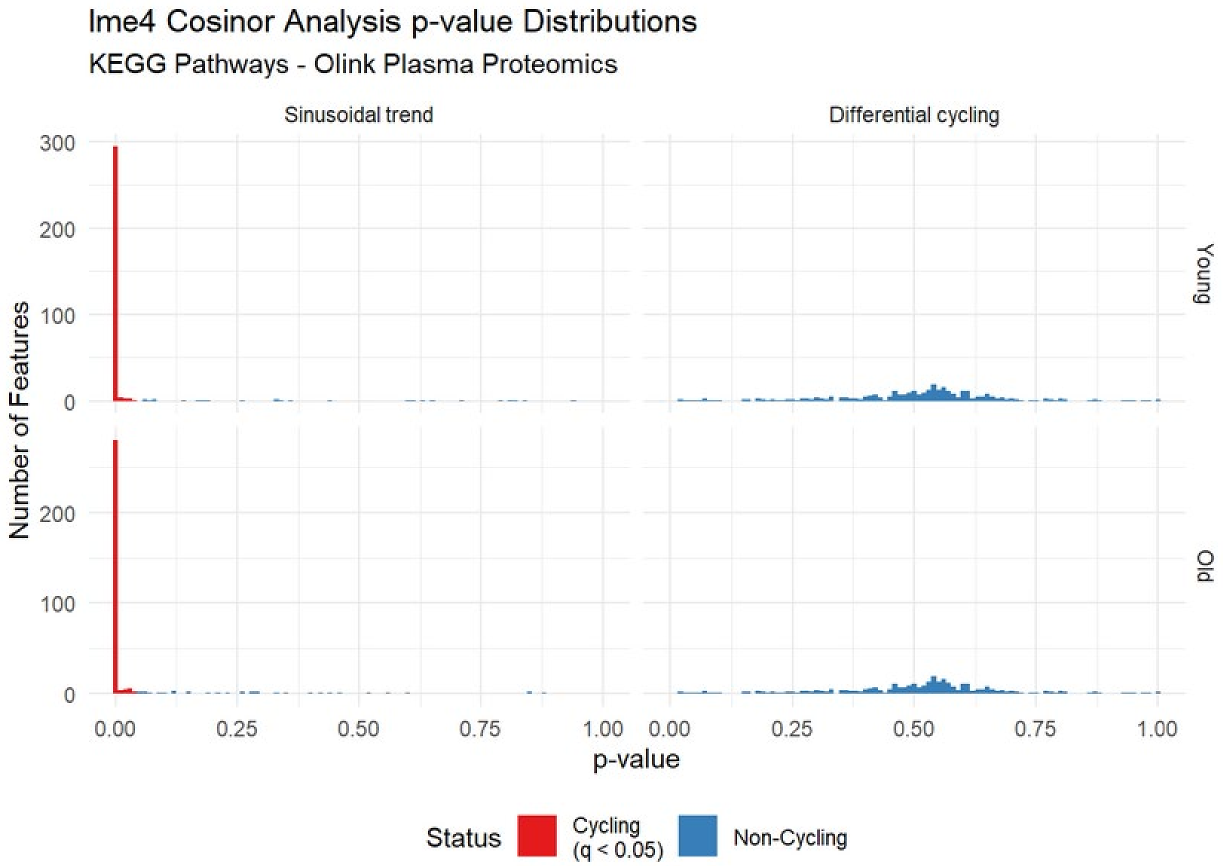

Significant cycler in the AB-proteome (Olink) KEGG Pathway Quantification, Cosinor q-value cutoff of < 0.05.

##### *Supplementary Figure 8*

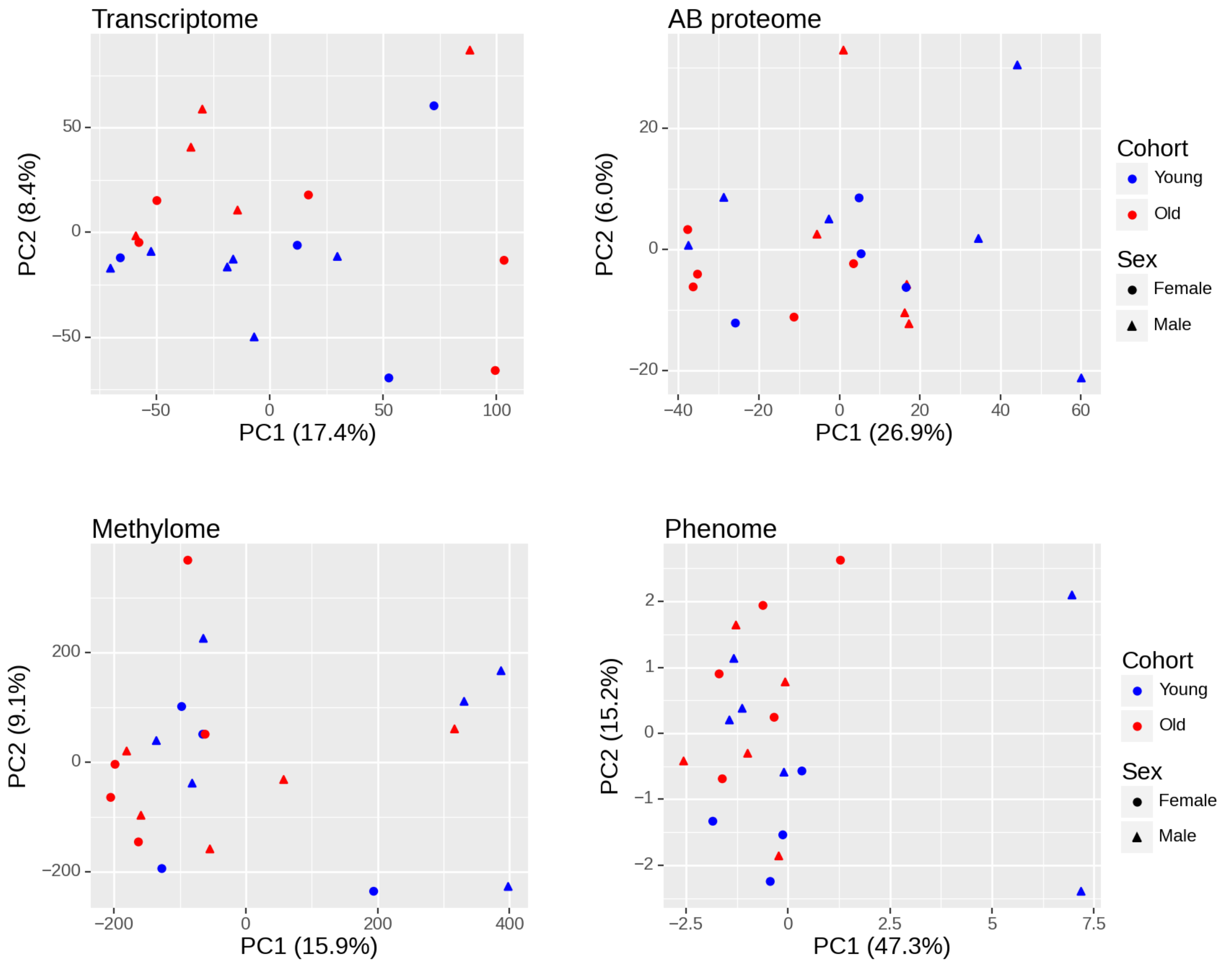

(A) Unsupervised principal component analysis on amplitudes of features from each omic data set.

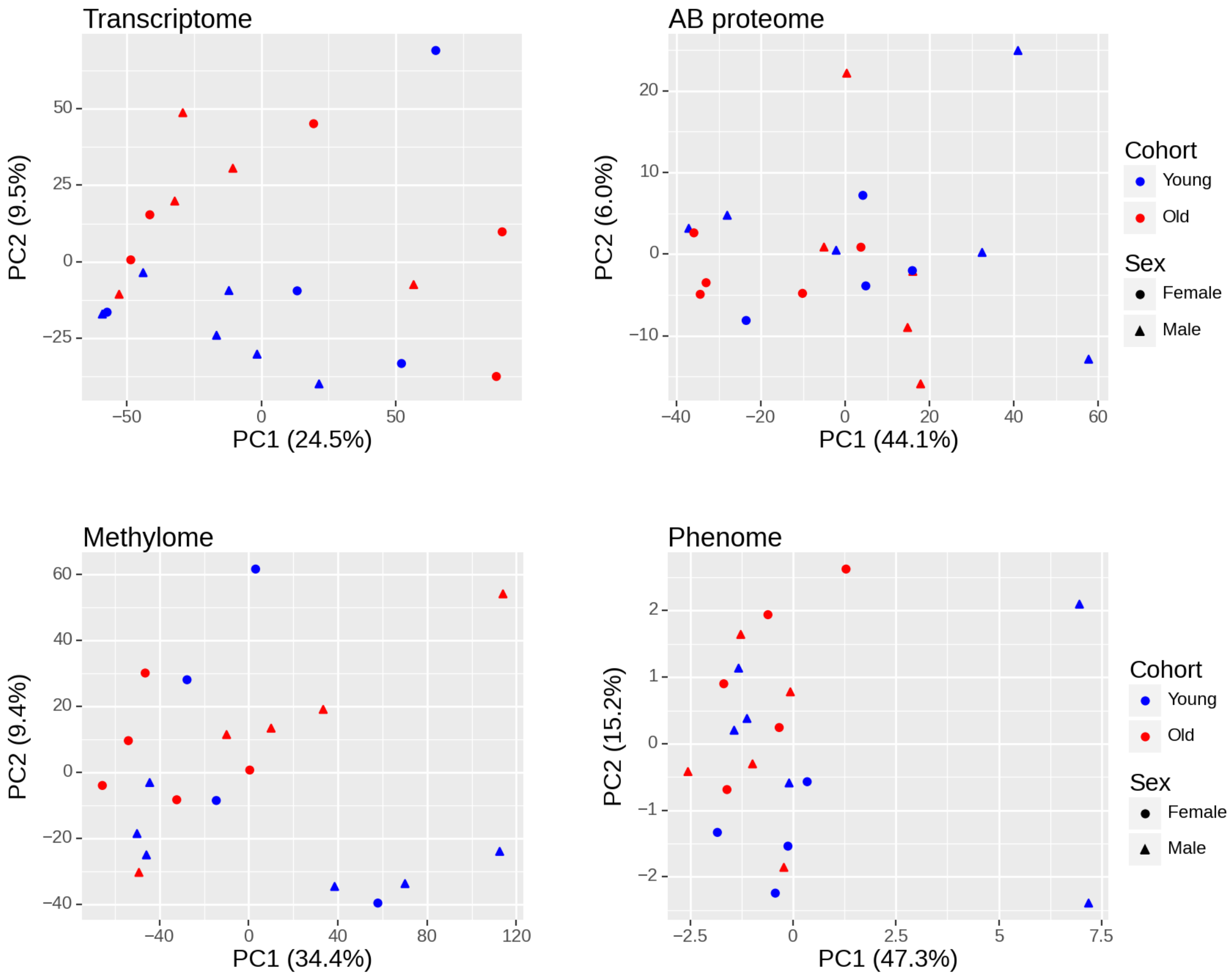

(B) Unsupervised principal component analysis on amplitudes of only the oscillatory features from each omic data set.

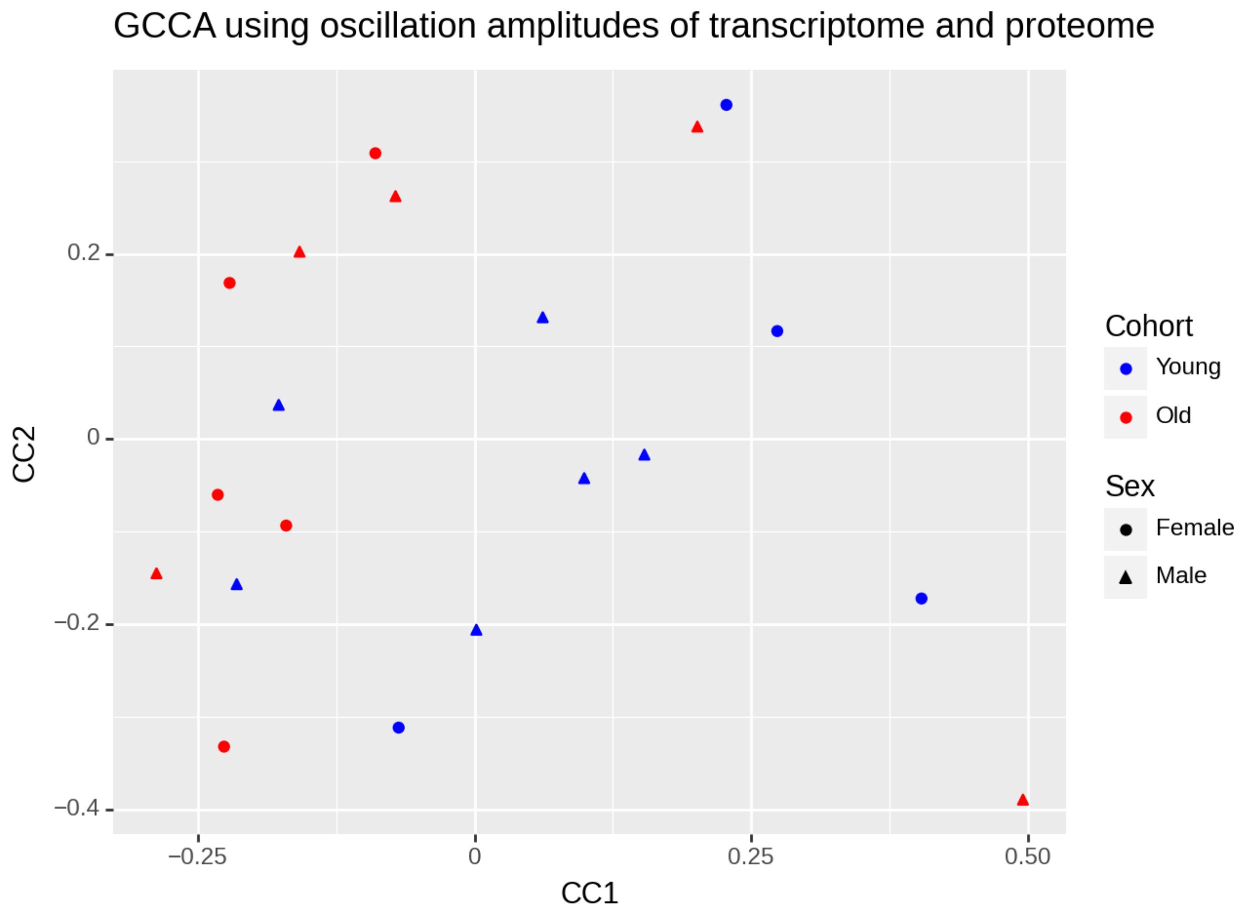

(C) Unsupervised generalized canonical correlation analysis on amplitudes of oscillating features from the transcriptome and the AB-Proteome.

***Supplementary Figure 9***

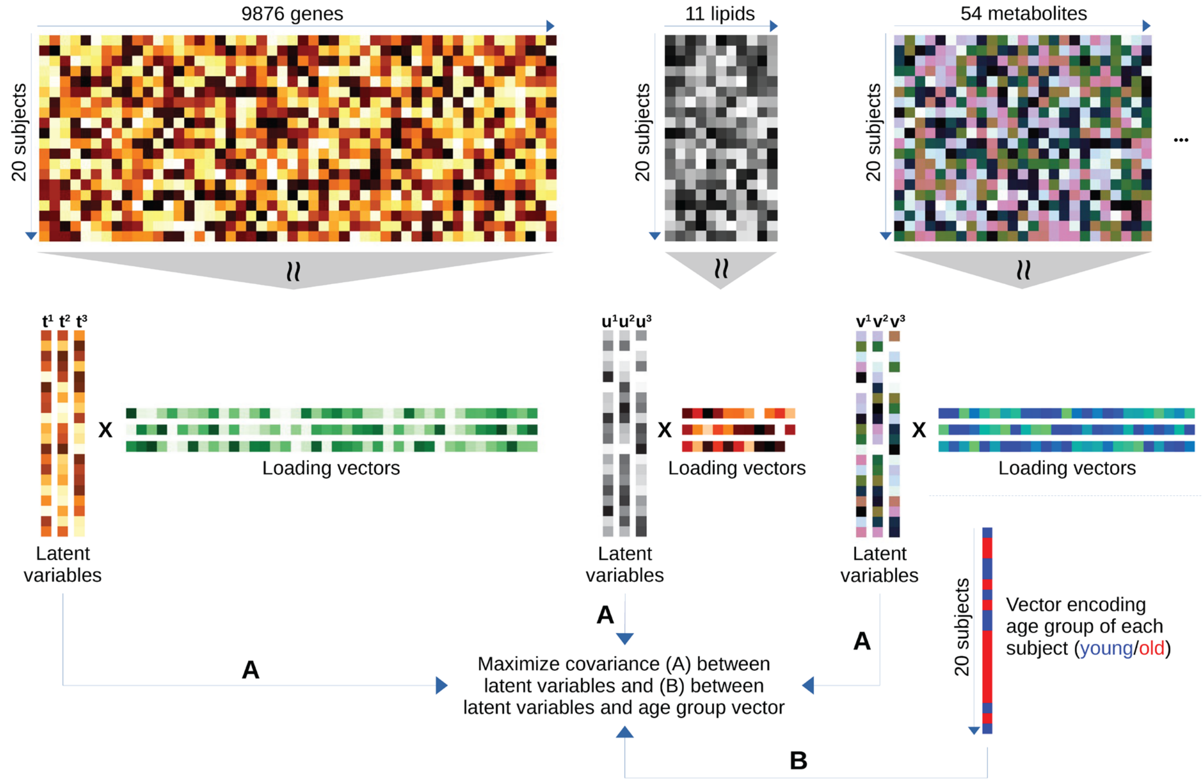

**Latent variables (components) identification by mixOmics.** First latent variables **t1**, **u1**, **v1** are determined by simultaneously maximizing (A) the covariance amongst the latent variables and (B) the covariance between the latent variables and the vector encoding the age group of the subjects. These two optimization goals are usually competing, so their importance must be explicitly weighted. The second latent variables **t2**, **u2**, **v2** are obtained using the same procedure applied on the complement of the space of the first latent variables, and similarly for higher latent variables. The LASSO optimization used aims to produce latent variables comprising of a small number of features (e.g., genes), to aid interpretation.

#### *Supplementary Figure 10*

Hub features with Pearson correlation condition of

|r|≥0.8 |r|≥0.65

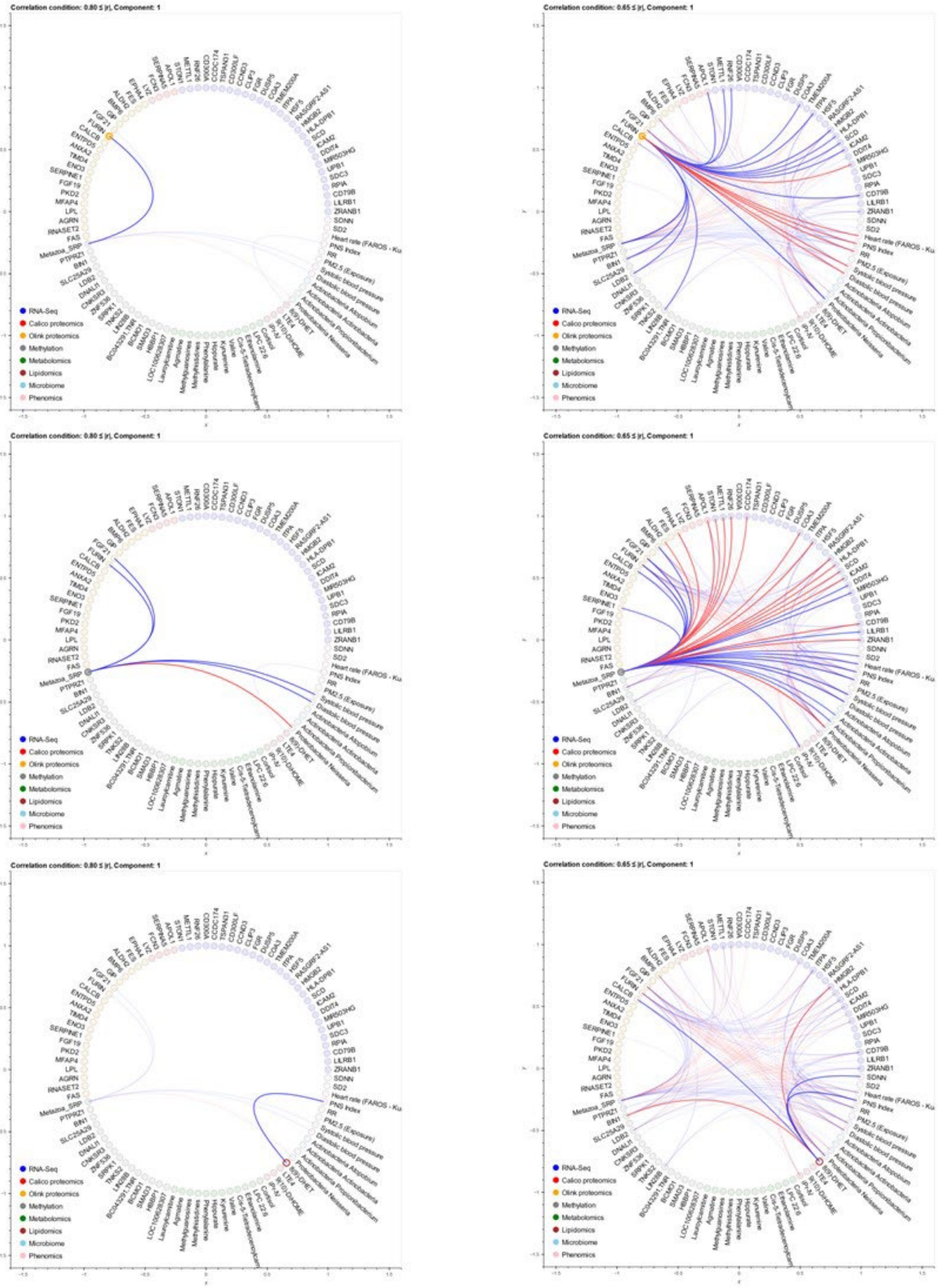

Hub features with Pearson correlation condition of

|r|≥0.8 |r|≥0.65

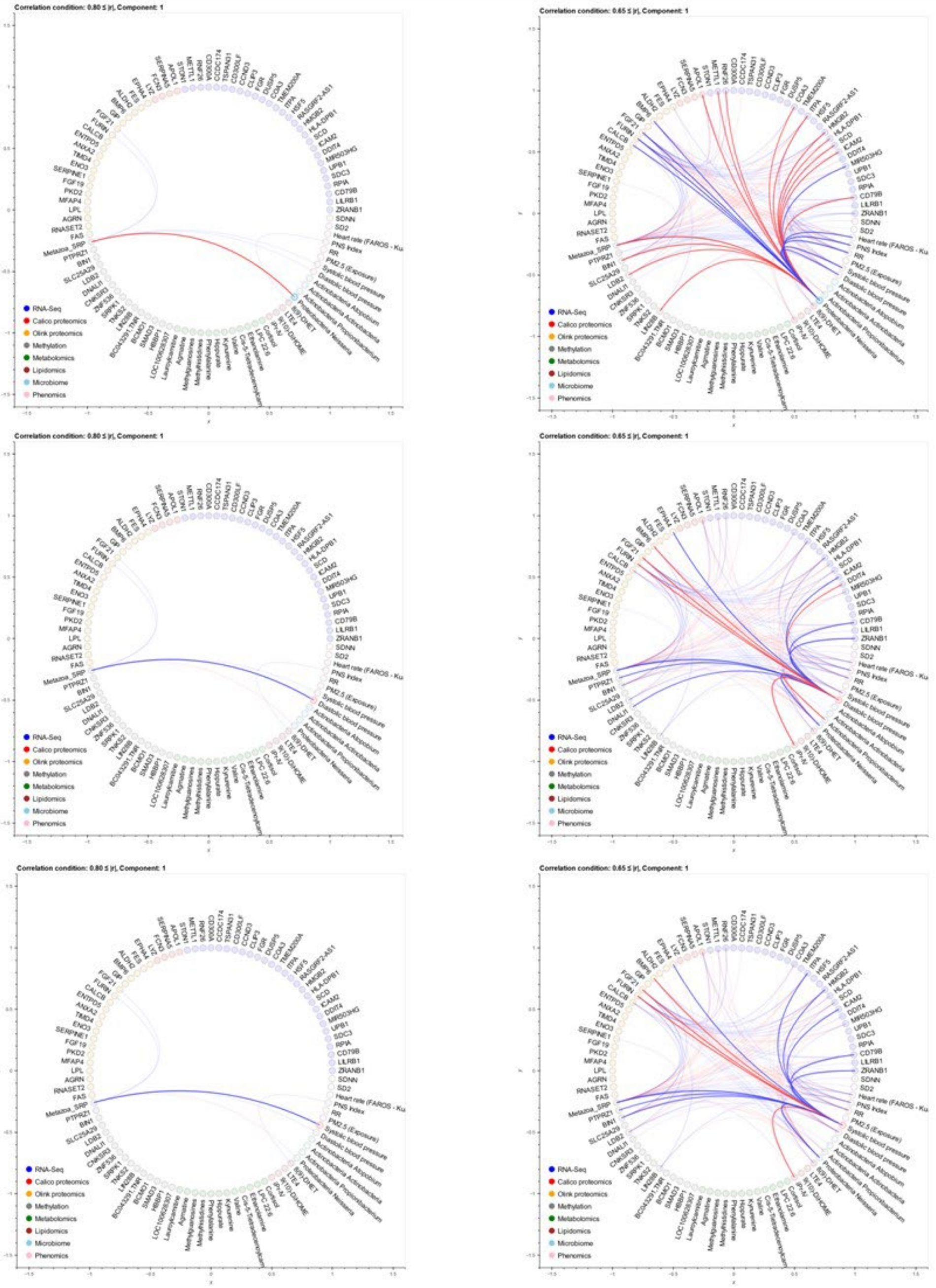

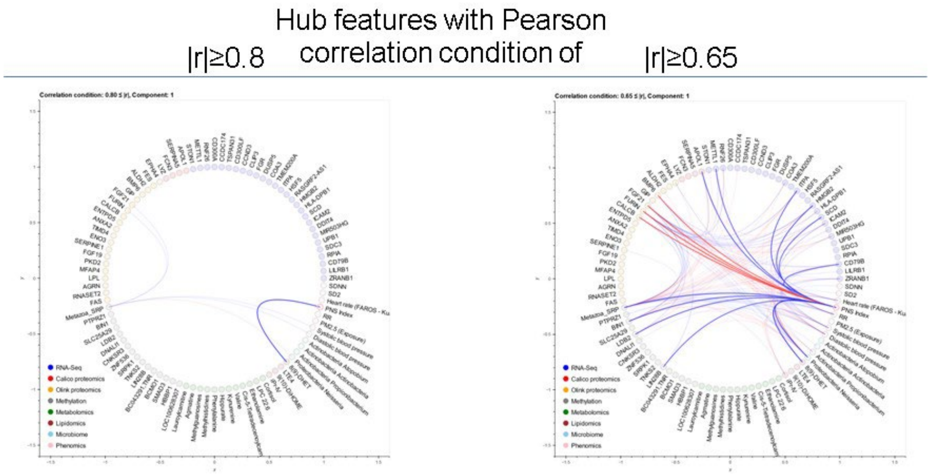

Hub features with Pearson correlation condition of |r|≥0.8 (left) to show transomic correlation linkage at a high stringency level, and of |r|≥0.65 to visualize the high number of transomic correlations.

***Supplementary Figure 11***

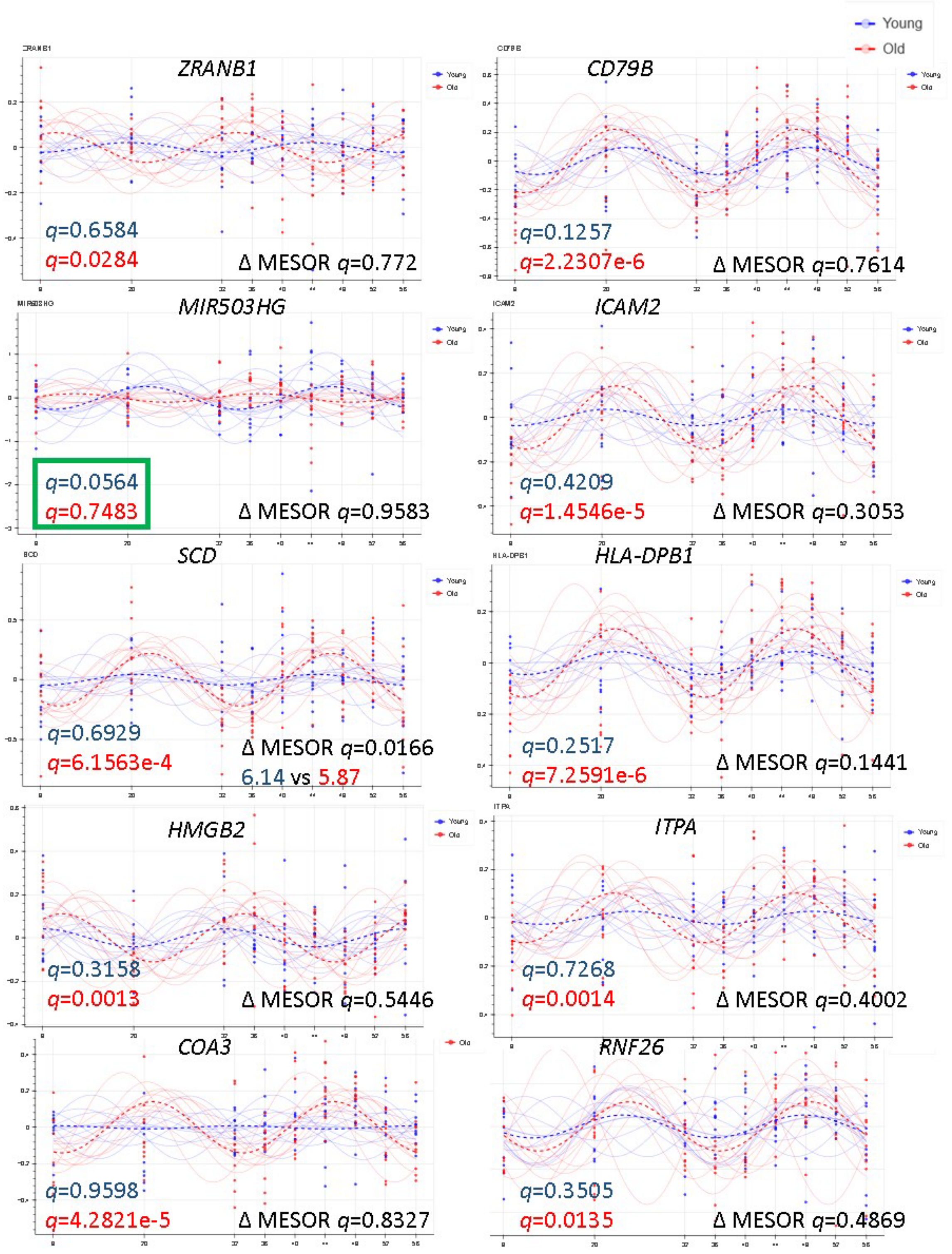

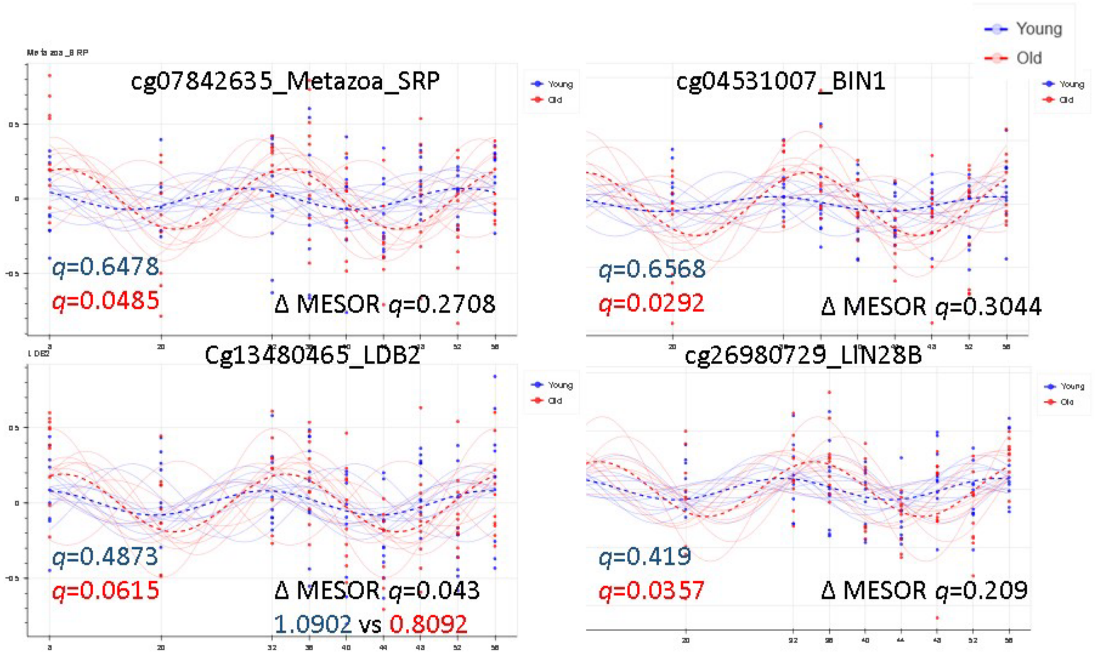

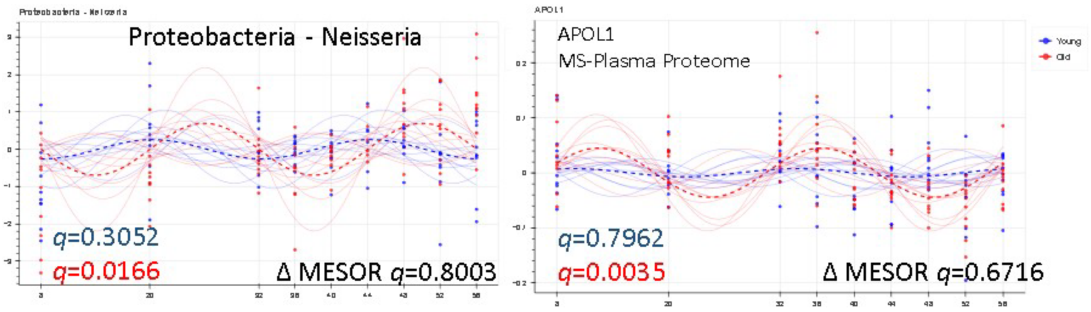

Age-dependent inversion of transomic oscillatory patterns is dominated by comparative features (FGF21 is the cross-correlated reference feature) with cyclic features detected in the old but not the young.

***Supplementary Figure 12***

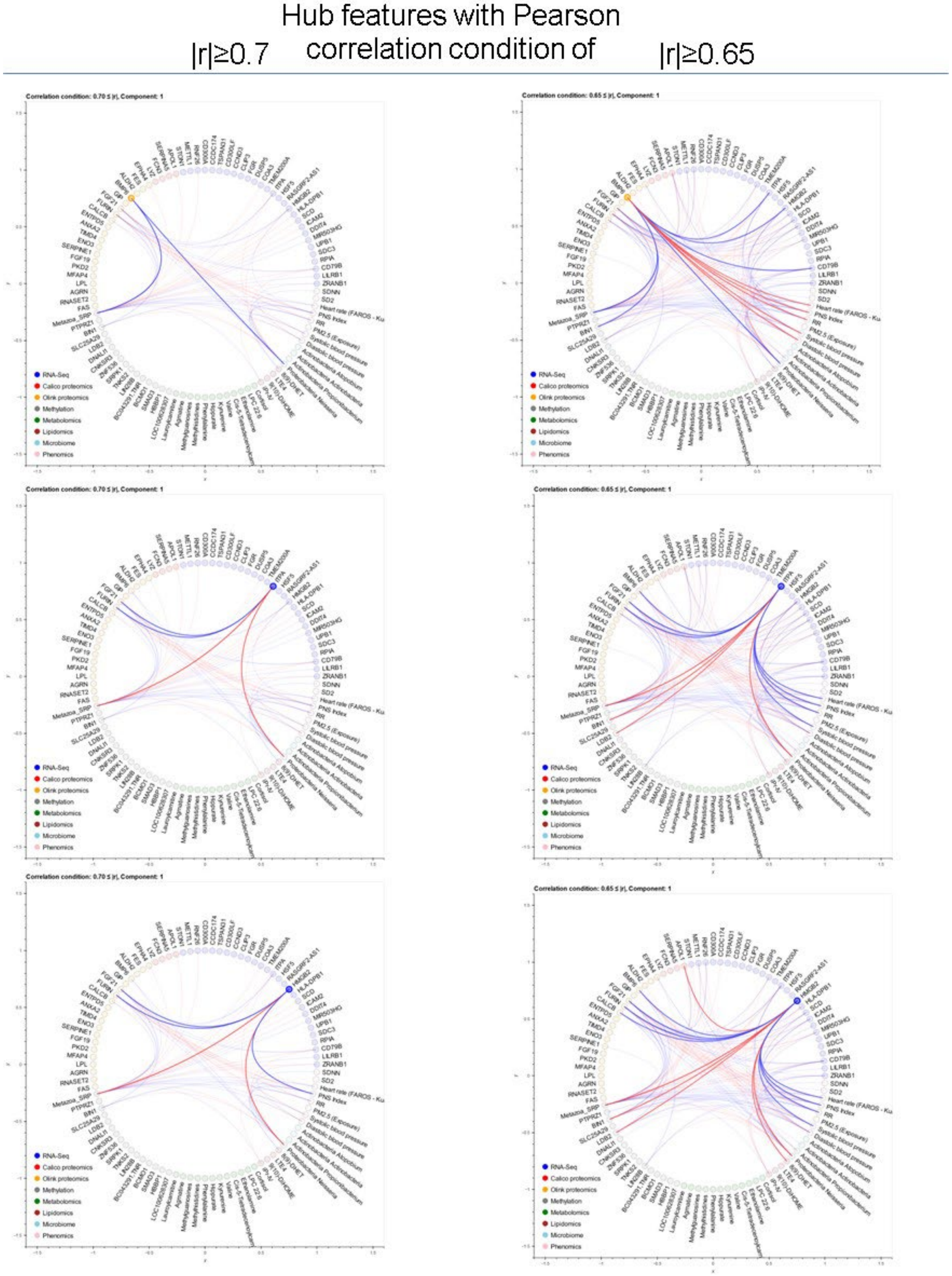

Hub features with Pearson correlation condition of

|r|≥0.7 |r|≥0.65

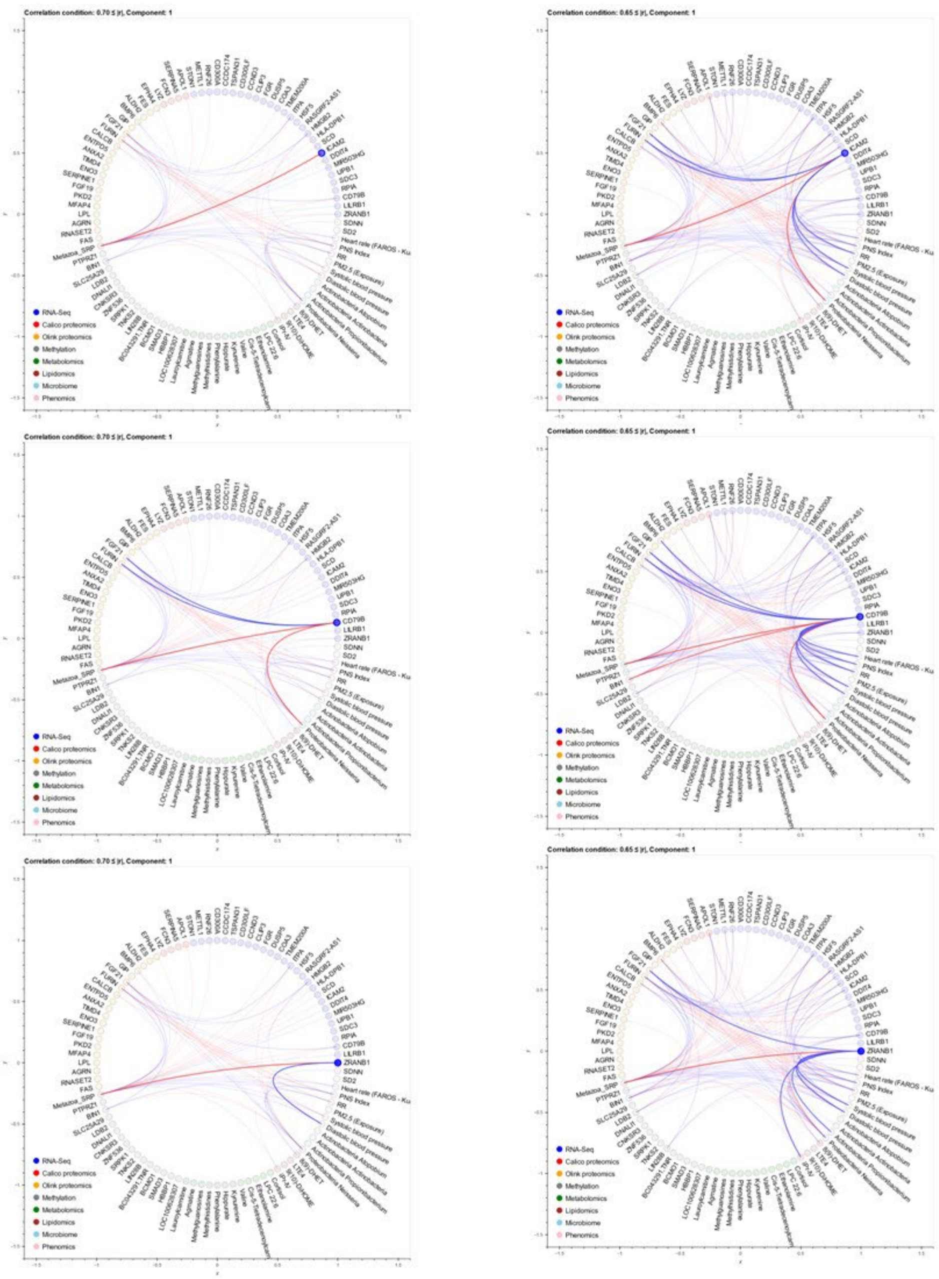

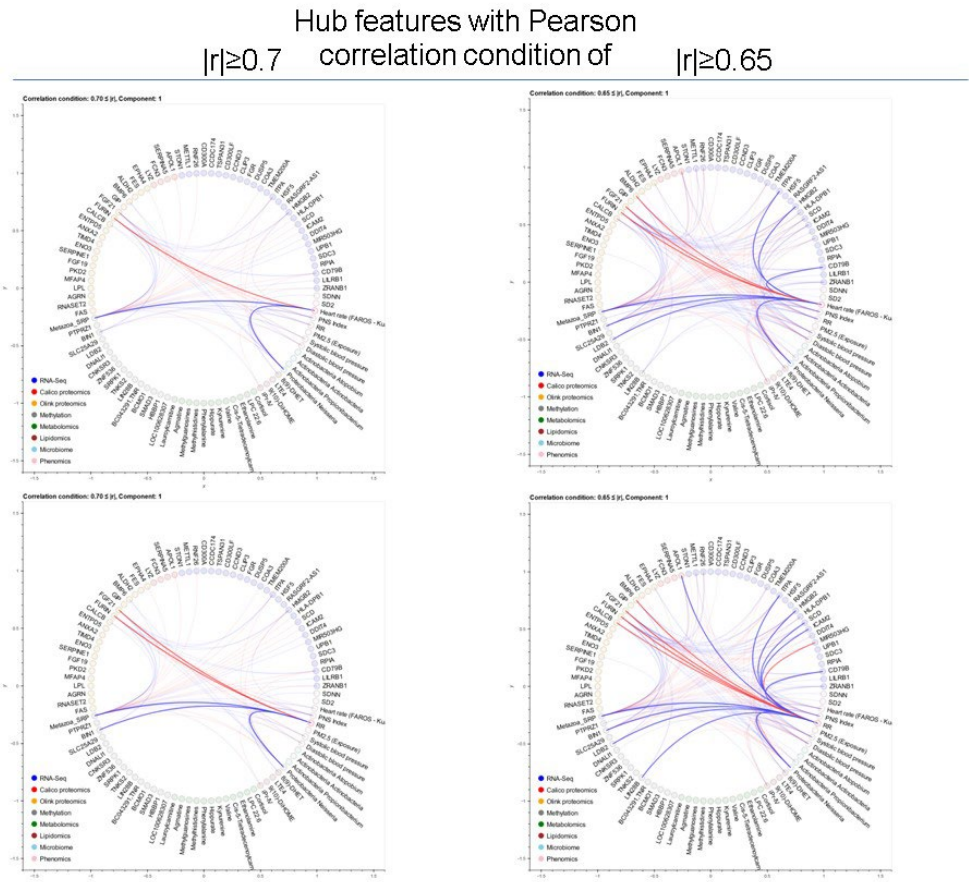

Hub features with Pearson correlation condition of |r|≥0.7 (left) to show transomic correlation linkage at a high stringency level, and of |r|≥0.65 to visualize the high number of transomic correlations.

##### *Supplementary Figure 13*

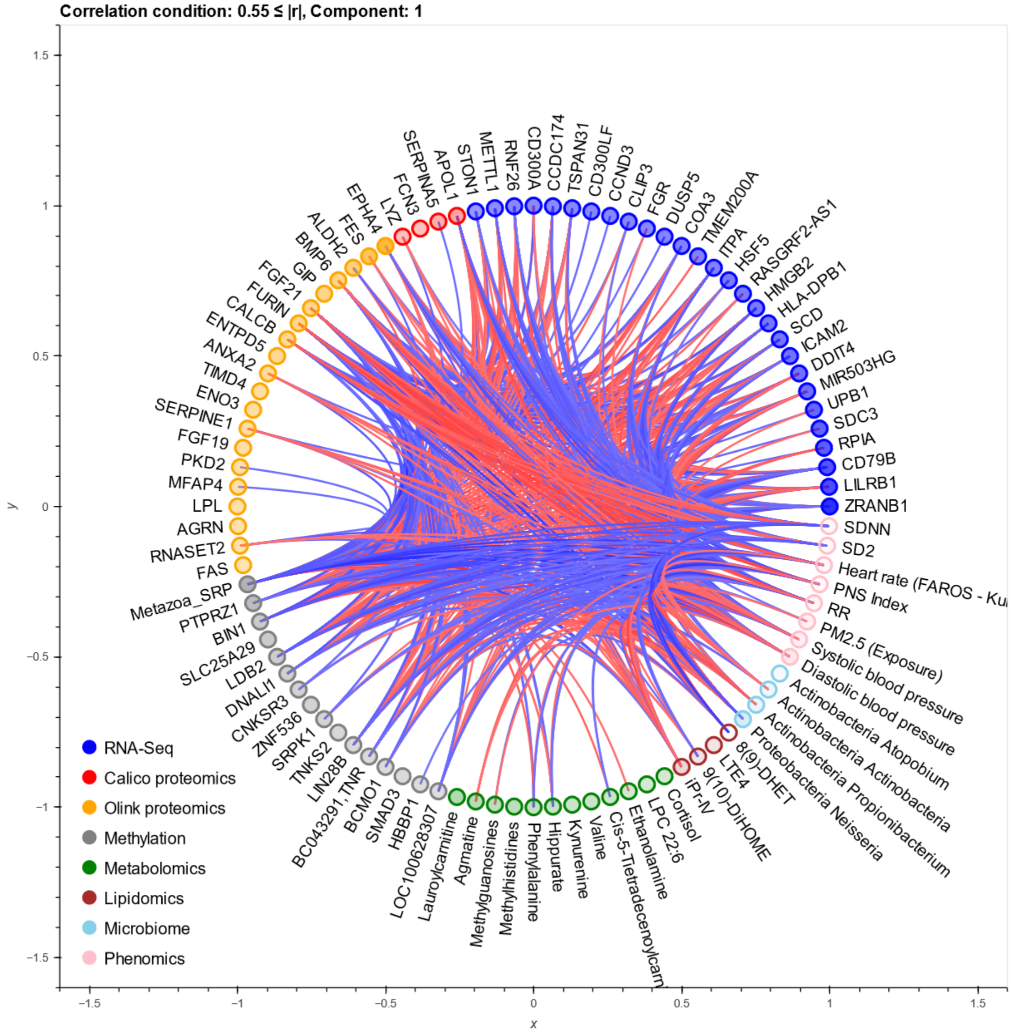

In contrast to oscillatory hub features, several features in each modality do not cross-correlate with other features even at the low Pearson correlation condition of |r|≥0.5.

***Supplementary Figure 14***

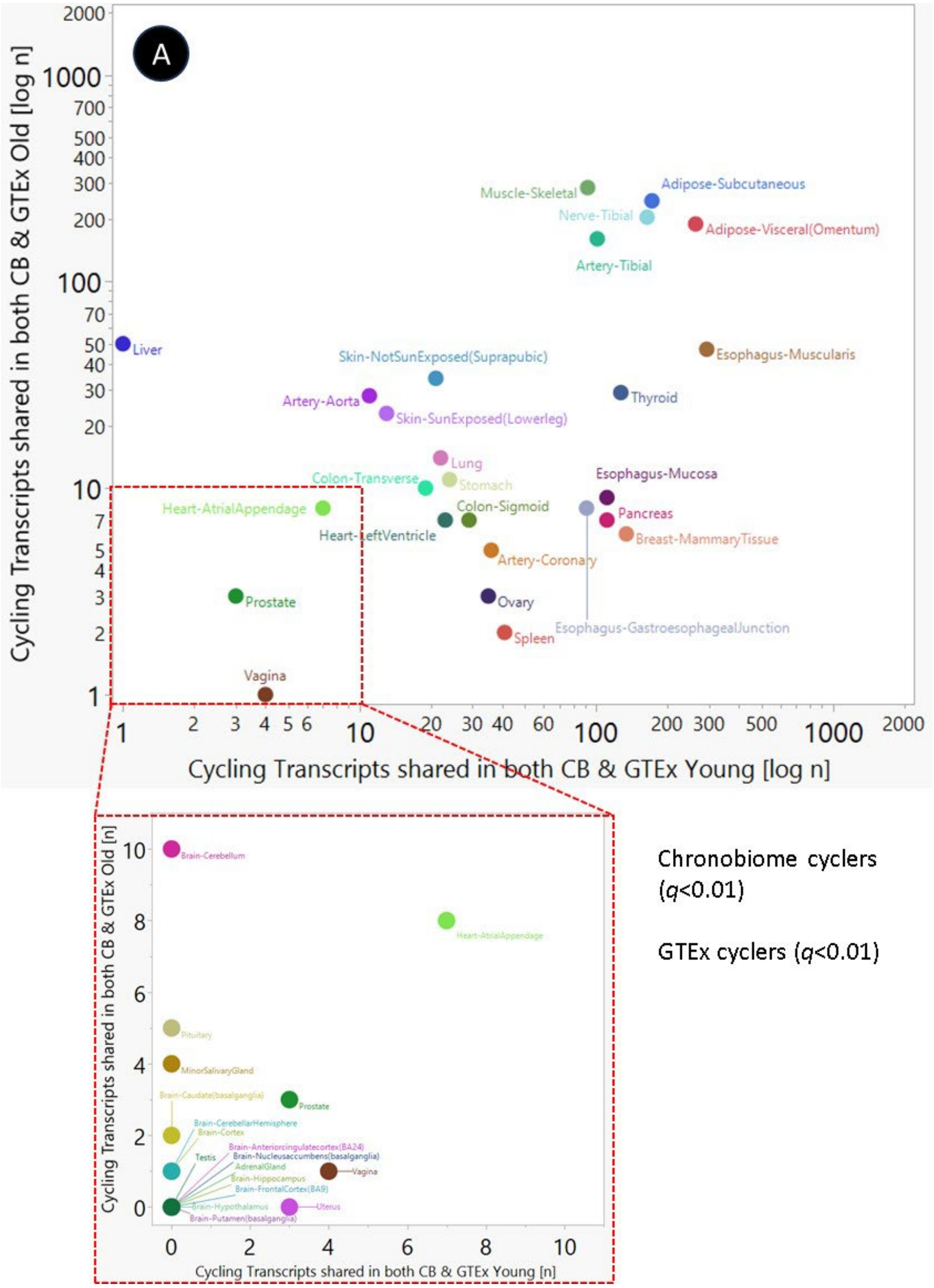

(A) Comprehensive gene expression studies in tissues donated by volunteers to the Adult Genotype Tissue Expression (GTEx) Project enabled to determine the overlap of rhythmic transcripts between these deceased GTEx donors and the alive aging chronobiome volunteers. Transcripts oscillating in the plasma chronobiome ($q$<0.01) aligned with the cycling tissue transcripts across 26 different deceased donor tissues ($q$<0.01).

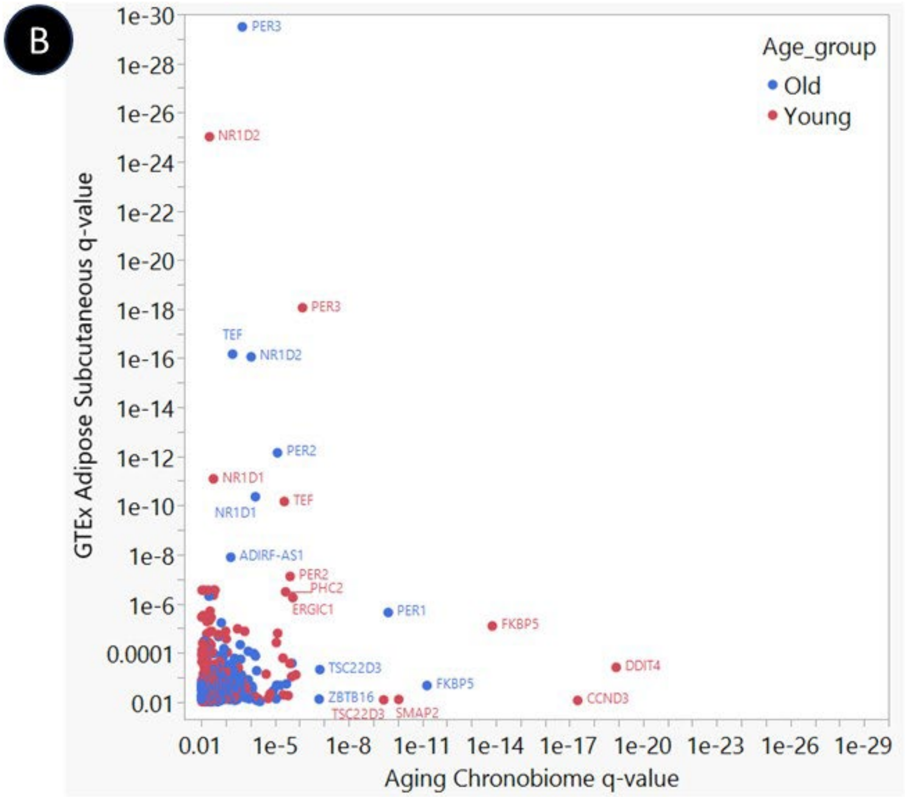
B
GTEx Adipose Subcutaneous q-value
Aging Chronobiome q-value
1e-30
1e-28
1e-26
1e-24
1e-22
1e-20
1e-18
1e-16
1e-14
1e-12
1e-10
1e-8
1e-6
0.0001
0.01
0.01 1e-5 1e-8 1e-11 1e-14 1e-17 1e-20 1e-23 1e-26 1e-29
Age_group
Old
Young
PER3
NR1D2
PER3
TEF
NR1D2
PER2
NR1D1
TEF
NR1D1
ADIRF-AS1
PER2
PHC2
ERGIC1
PER1
FKBP5
DDIT4
TSC22D3
FKBP5
ZBTB16
TSC22D3
SMAP2
CCND3

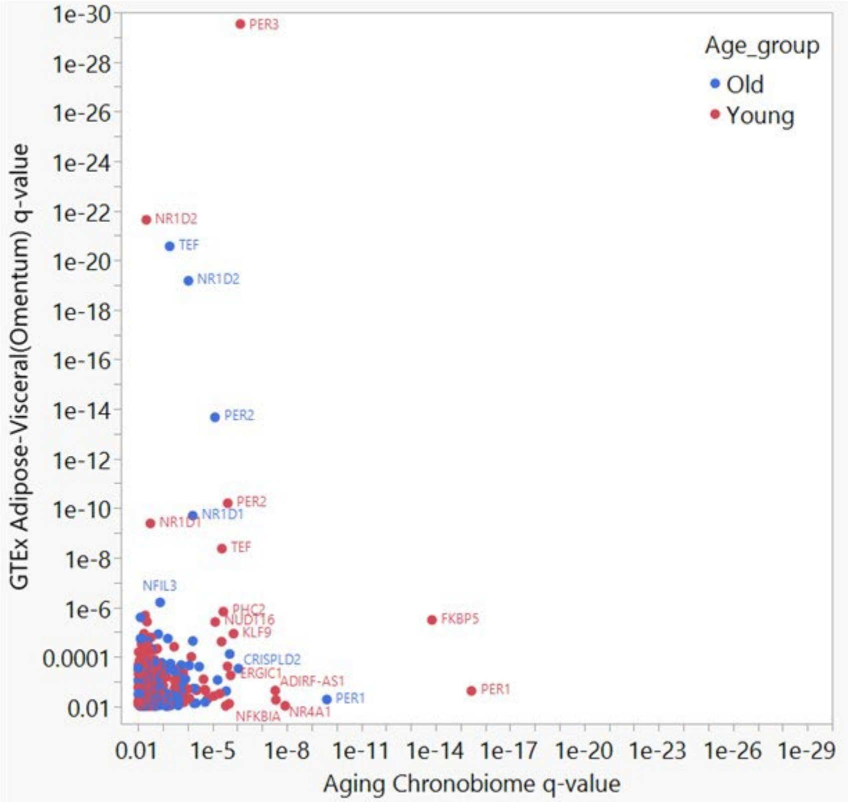
GTEx Adipose-Visceral(Omentum) q-value
Aging Chronobiome q-value
1e-30
1e-28
1e-26
1e-24
1e-22
1e-20
1e-18
1e-16
1e-14
1e-12
1e-10
1e-8
1e-6
0.0001
0.01
0.01 1e-5 1e-8 1e-11 1e-14 1e-17 1e-20 1e-23 1e-26 1e-29
Age_group
Old
Young
PER3
NR1D2
TEF
NR1D2
PER2
PER2
NR1D1
NR1D1
TEF
NFIL3
PHC2
NUDT16
KLF9
FKBP5
CRISPLD2
ERGIC1
ADIRF-AS1
PER1
PER1
NFKBIA
NR4A1

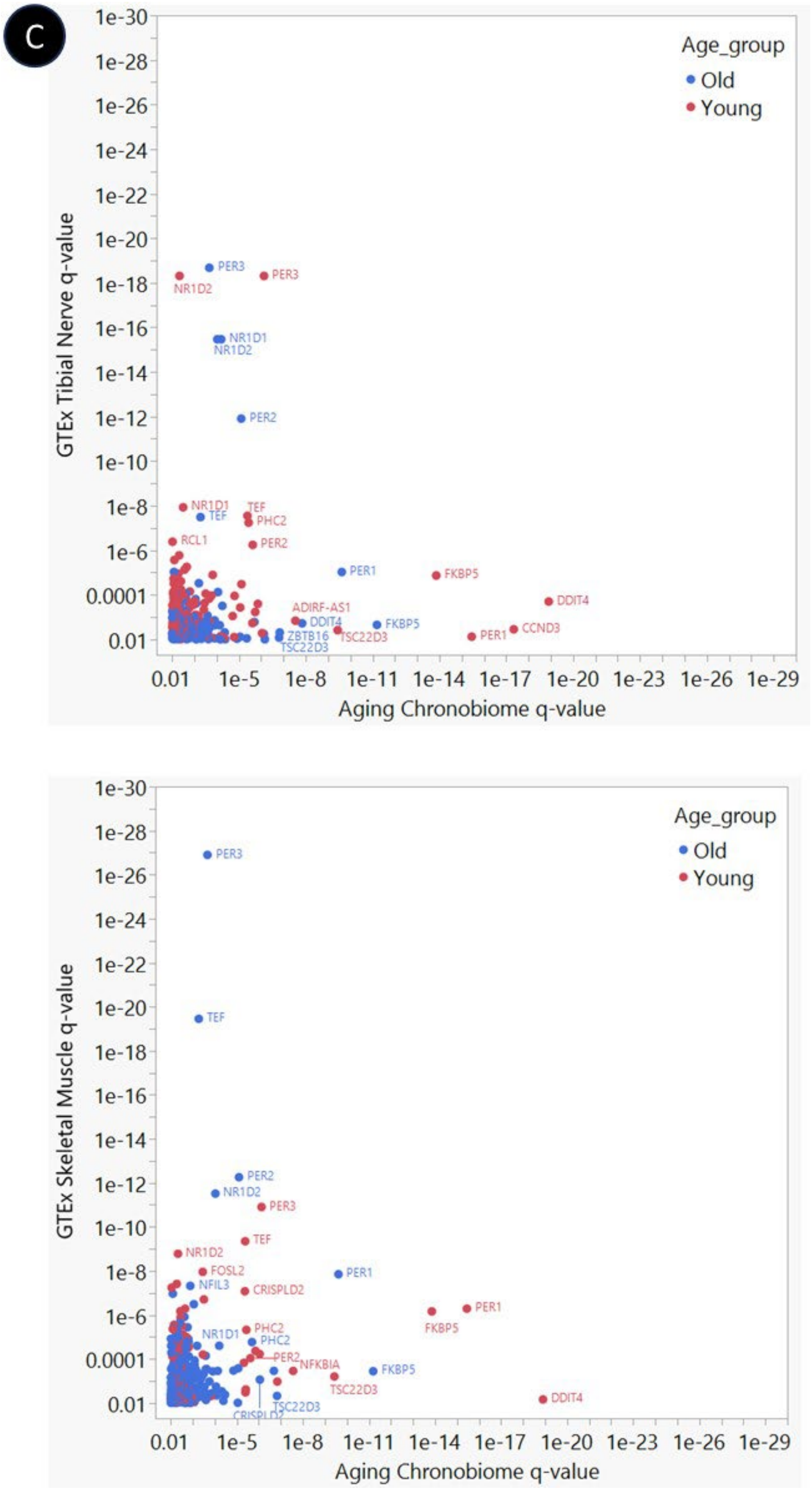
C
Age_group
Old
Young
GTEx Tibial Nerve q-value
Aging Chronobiome q-value
PER3
NR1D2
NR1D1
PER2
TEF
PHC2
RCL1
PER1
FKBP5
DDIT4
ADIRF-AS1
ZBTB16
TSC22D3
CCND3
GTEx Skeletal Muscle q-value
FOSL2
NFIL3
CRISPLD2
NFKBIA
1e-30
1e-28
1e-26
1e-24
1e-22
1e-20
1e-18
1e-16
1e-14
1e-12
1e-10
1e-8
1e-6
0.0001
0.01
1e-5
1e-11
1e-17
1e-23
1e-26
1e-29

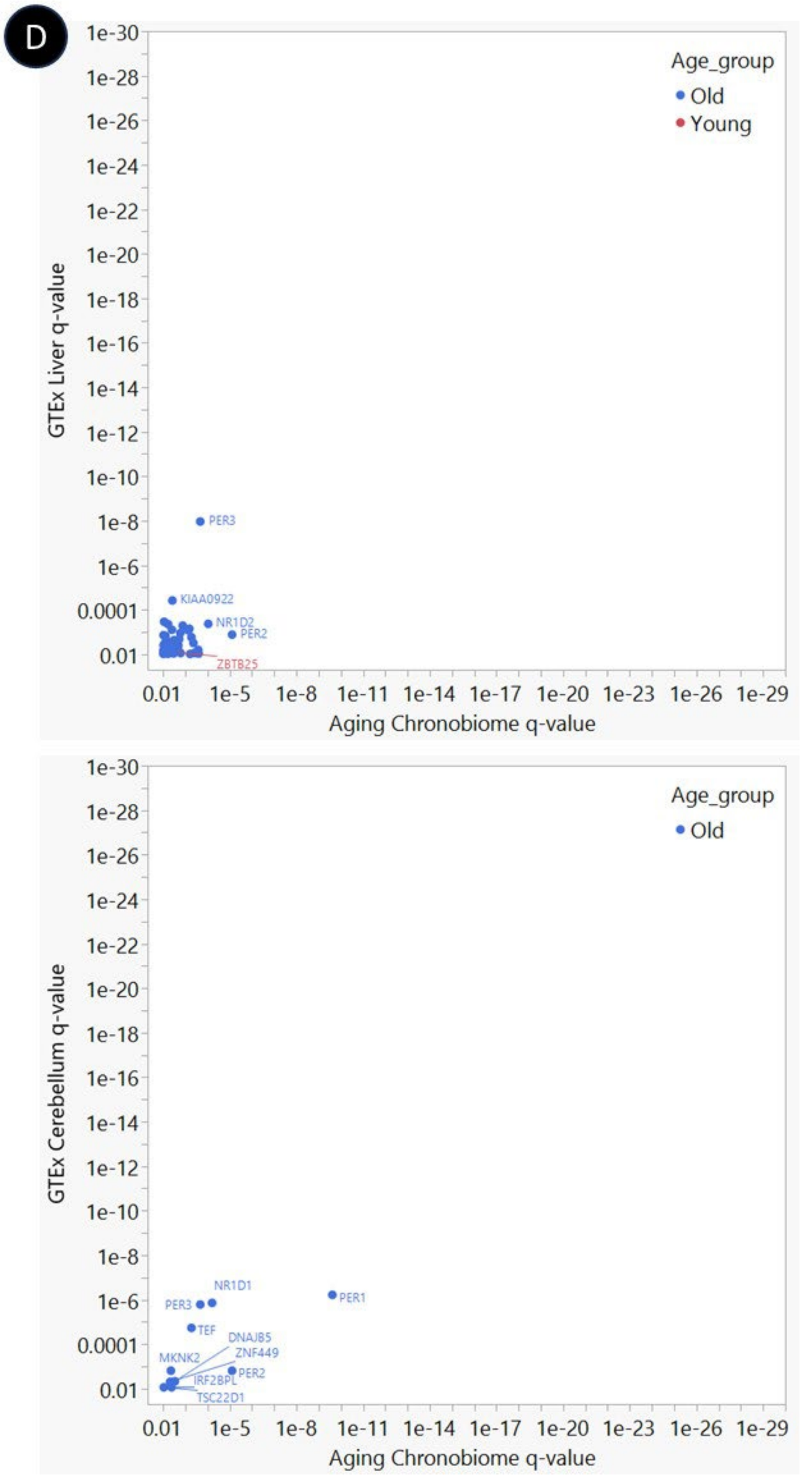

Overlapping cycling transcripts were prominent in the GTEx tissues from subcutaneous adipose (B top), skeletal muscle (B bottom), tibial nerve (C top) and visceral adipose from the omentum (C bottom). In contrast, liver (D top) and cerebellum (D bottom) showed limited overlapping cycling gene expression.

***Supplementary Figure 15***

### (A) Liver Disease

UKBB Incident Cases (n): 432
UKBB Controls (n): 47,104
UKBB Mean±SD Years to Incident Disease Diagnosis: 7±3.3

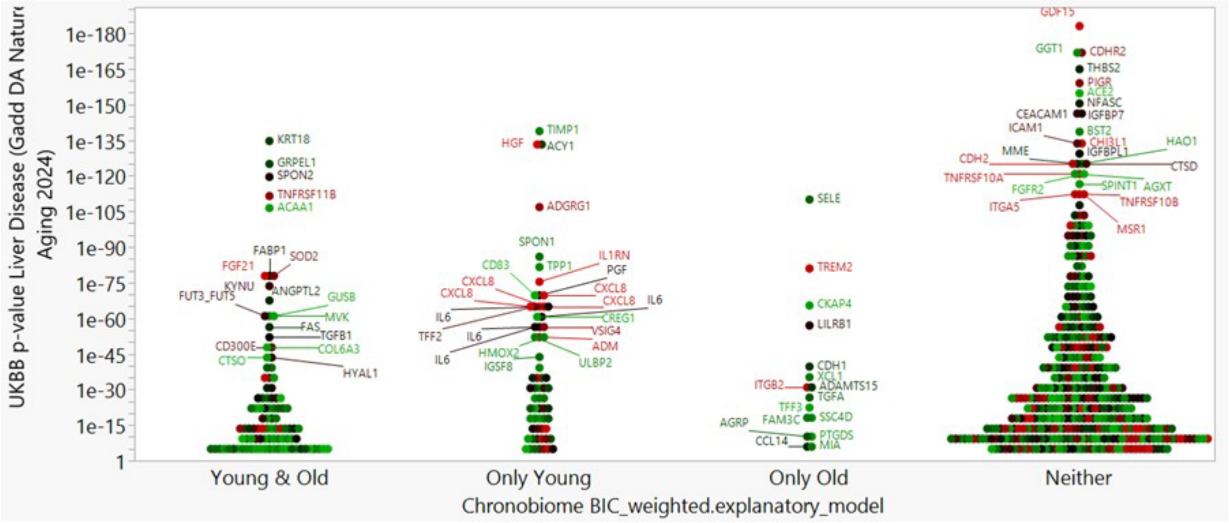

n=99 n=64 n=16 n=408

**UKBB p<3.1e-6 & Chronobiome BIC>0.75**

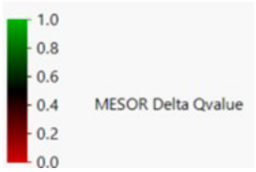

### (B) Parkinson's Disease

UKBB Incident Cases (n): 659
UKBB Controls (n): 46,802
UKBB Mean±SD Years to Incident Disease Diagnosis: 5.4±3.2

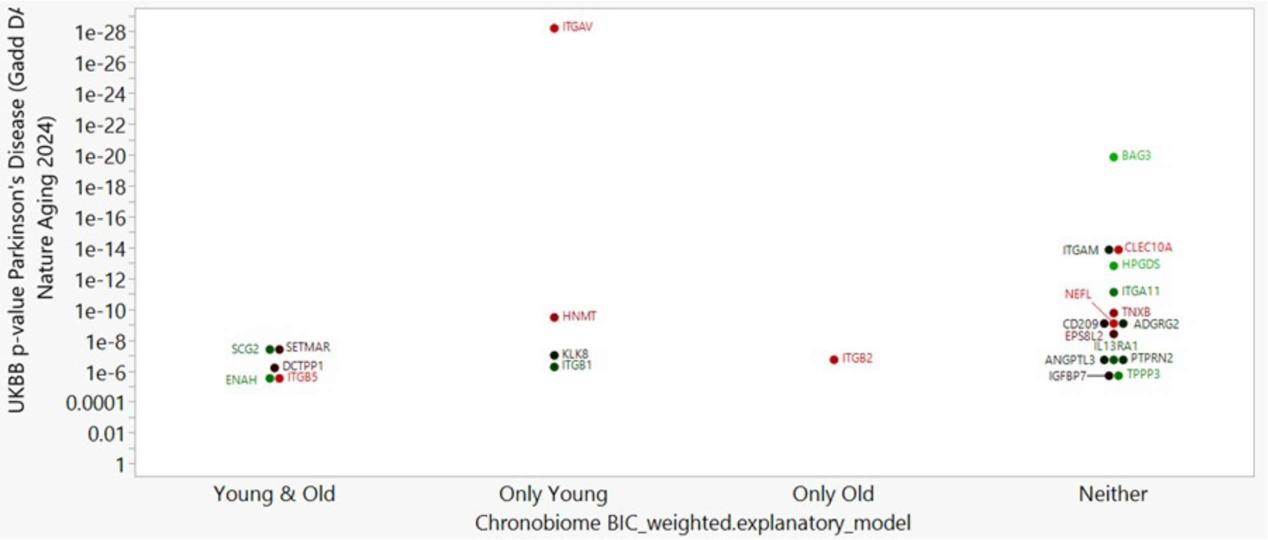

**n=5** **n=4** **n=1** **n=15**

**UKBB p<3.1e-6 & Chronobiome BIC>0.75**

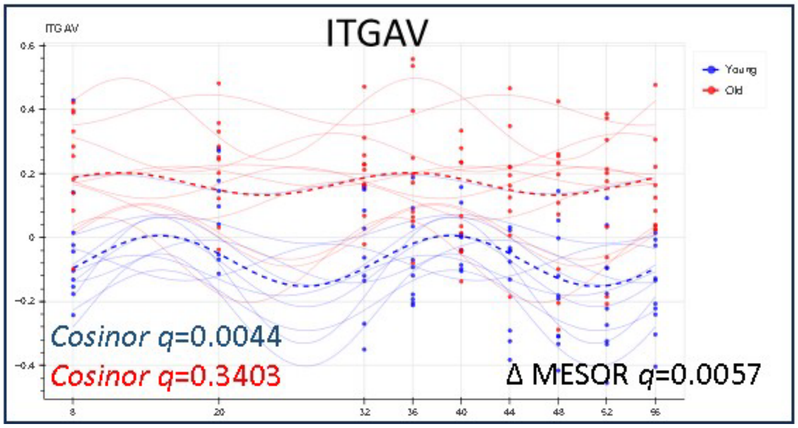

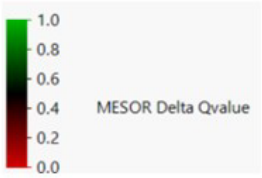

### (C) COPD

UKBB Incident Cases (n): 1,998
UKBB Controls (n): 44,948
UKBB Mean±SD Years to Incident Disease Diagnosis: 6.3±3.4

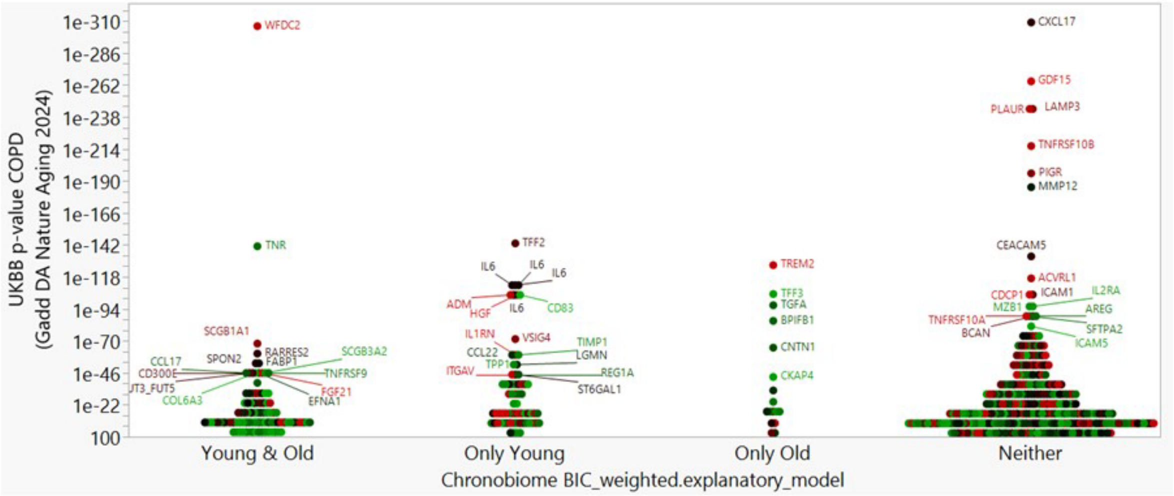

**n=95** **n=68** **n=17** **n=324**

**UKBB p<3.1e-6 & Chronobiome BIC>0.75**

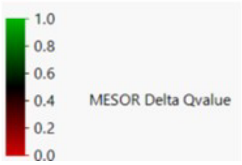

### (D) Ischemic Stroke

UKBB Incident Cases (n): 765
UKBB Controls (n): 46,657
UKBB Mean±SD Years to Incident Disease Diagnosis: 6.8±3.4

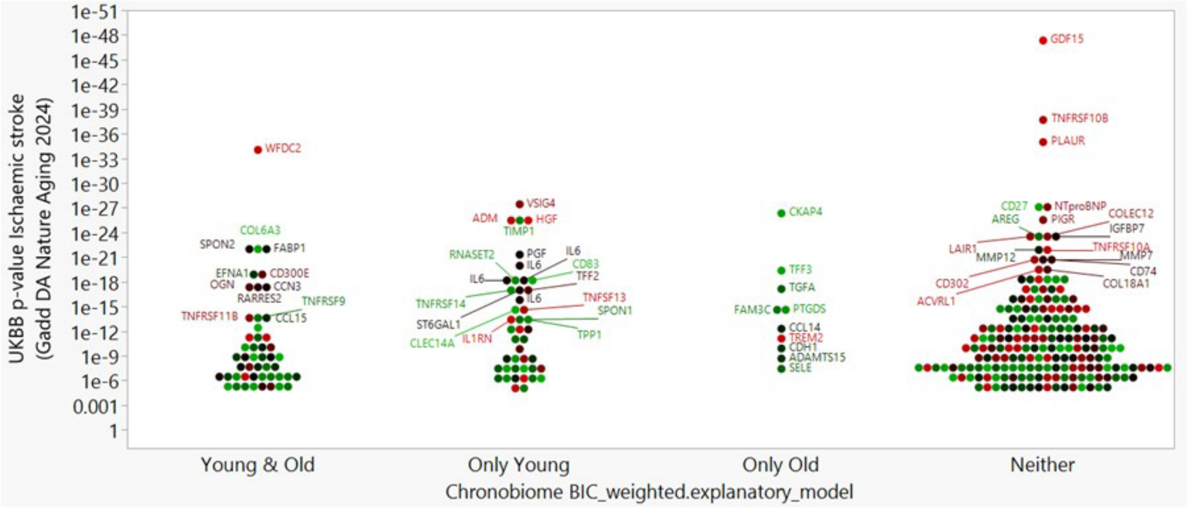

**n=49** **n=43** **n=10** **n=190**

**UKBB p<3.1e-6 & Chronobiome BIC>0.75**

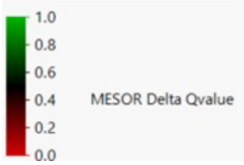

### (E) Rheumatoid Arthritis

UKBB Incident Cases (n): 593
UKBB Controls (n): 46,310
UKBB Mean±SD Years to Incident Disease Diagnosis: 6.8±3.2

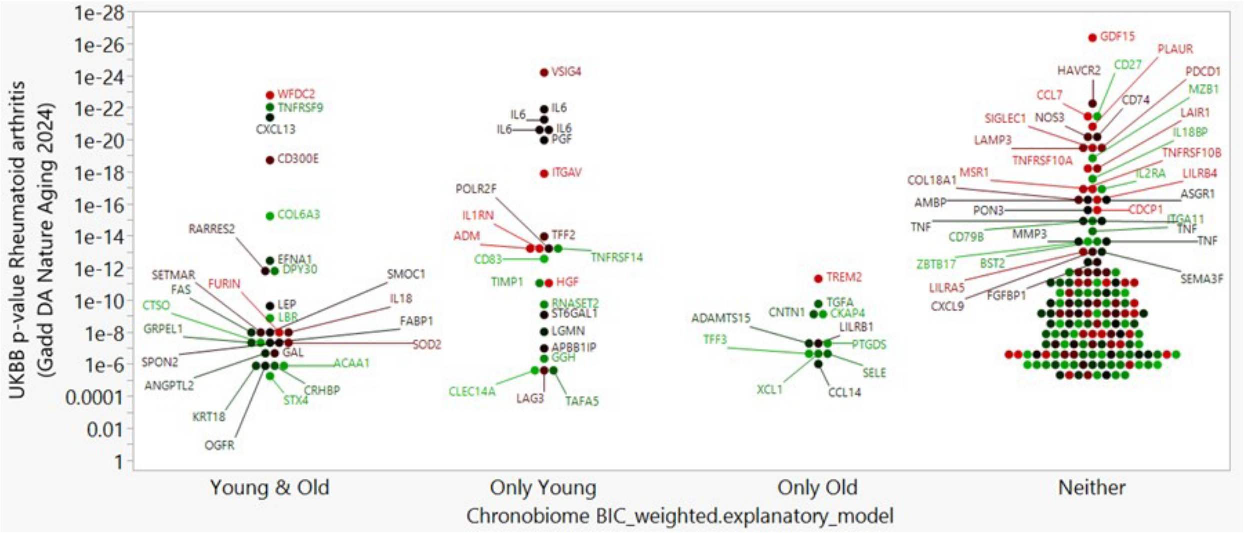

n=27 n=23 n=11 n=156

UKBB p<3.1e-6 & Chronobiome BIC>0.75

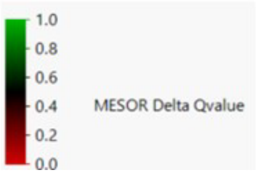

### (F) Lung Cancer

UKBB Incident Cases (n): 403
UKBB Controls (n): 47,158
UKBB Mean±SD Years to Incident Disease Diagnosis: 5.9±3.2

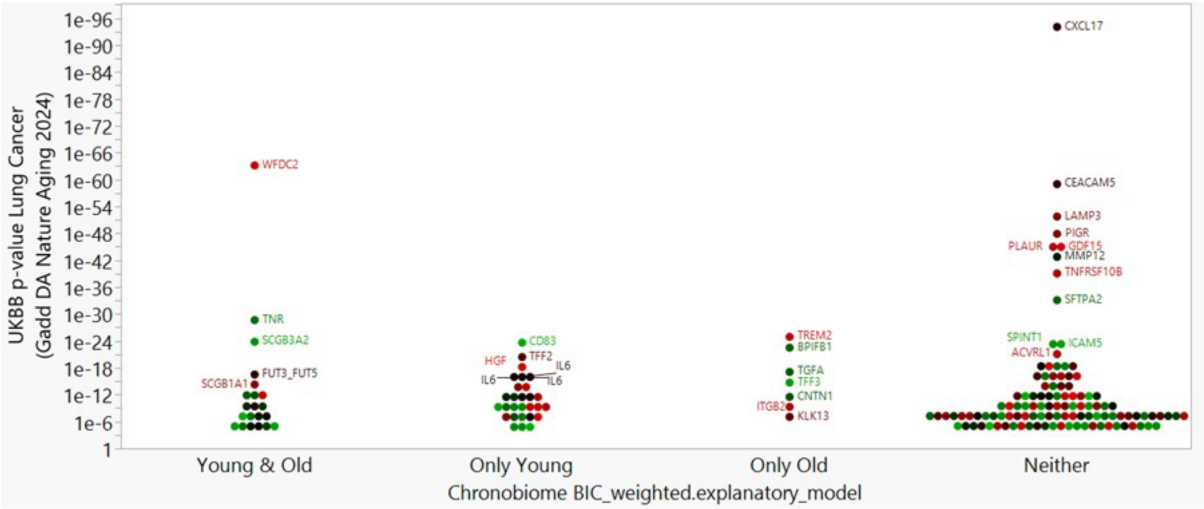

**n=21** **n=28** **n=7** **n=112**

**UKBB p<3.1e-6 & Chronobiome BIC>0.75**

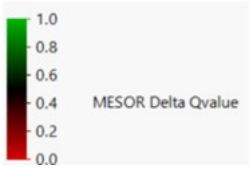

### (G) Systemic Lupus Erythematosus

UKBB Incident Cases (n): 134
UKBB Controls (n): 47,069
UKBB Mean±SD Years to Incident Disease Diagnosis: 5.1±2.6

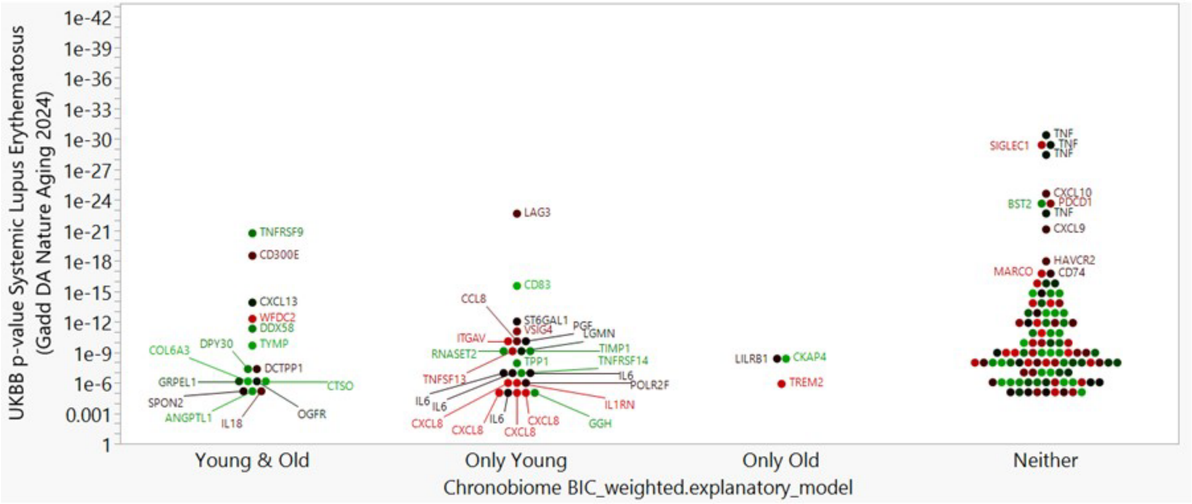

**n=15** **n=24** **n=3** **n=102**

**UKBB p<3.1e-6 & Chronobiome BIC>0.75**

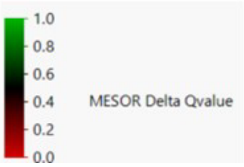

The overlap between the cycling AB-proteome (four categories on the abscissa: cycling in young and old, cycling in only the young, cycling in only the old, and cycling in neither young nor old) as determined by the chronobiome and the protein-disease association (ordinate) as determined by the UKBB [32] is shown for (A) liver disease, (B) Parkinson's disease, (C) COPD, (D) ischemic stroke, (E) rheumatoid arthritis, (F) lung cancer, and (G) systemic lupus erythematosus. The last category on the abscissa denotes the overlap between non-cycling and disease-related proteins. All proteins shown satisfy cut-off values for significance, chronobiome BIC>0.75, UKBB Bonferroni-adjusted p<3.1e-6. The sample sizes located under each graph indicate the number of cycling proteins for each of the three categories (cycling in young and old, cycling in only the young, and cycling in only the old), as well as the number of non-cycling proteins with disease prediction (neither cycling in young nor old). Data on UKBB Incident Cases (n), UKBB Controls (n), and UKBB Mean±SD Years to Incident Disease Diagnosis are taken from Table 1 as published in [32]. The color overlay indicates the chronobiome mesor delta q-value which highlights in red the proteins with a age-specific difference in abundance in the cycling and non-cycling proteome. Note that replicate feature measurements in each of the inflammation, neurology, oncology and cardiometabolic panels, as, for example, is the case for IL6, explain the multiple occurrences in the plots.

***Supplementary Figure 16***

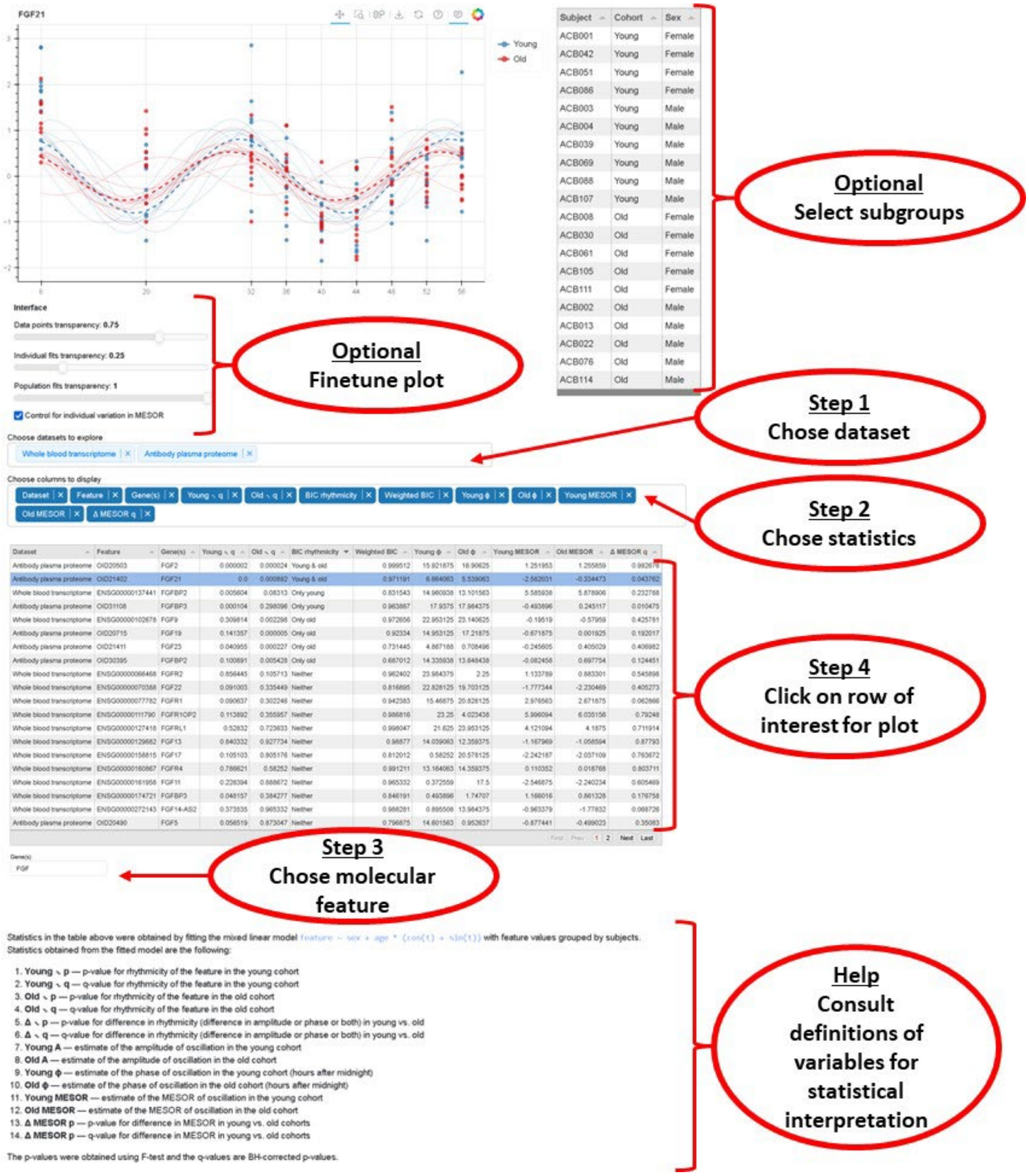

The chronobiome dashboard at https://chronobiome.org/ allows plotting of the data in four steps with options to change the display and help to interpret results. [*This resource becomes accessible once the peer review process concludes.*]
